# Retrospective evaluation of short-term forecast performance of ensemble sub-epidemic frameworks and other time-series models: The 2022-2023 mpox outbreak across multiple geographical scales, July 14^th^, 2022, through February 26th, 2023

**DOI:** 10.1101/2023.05.15.23289989

**Authors:** Amanda Bleichrodt, Ruiyan Luo, Alexander Kirpich, Gerardo Chowell

## Abstract

In May 2022, public health officials noted an unprecedented surge in mpox cases in non-endemic countries worldwide. As the epidemic accelerated, multi-model forecasts of the epidemic’s trajectory were critical in guiding the implementation of public health interventions and determining policy. As the case levels have significantly decreased as of early September 2022, evaluating model performance is essential to advance the growing field of epidemic forecasting.

Using laboratory-confirmed mpox case data from the Centers for Disease Control and Prevention (CDC) and Our World in Data (OWID) teams through the week of January 26th, 2023, we generated retrospective sequential weekly forecasts (e.g., 1-4-weeks) for Brazil, Canada, France, Germany, Spain, the United Kingdom, the USA, and at the global scale using models that require minimal input data including the auto-regressive integrated moving average (ARIMA), general additive model (GAM), simple linear regression (SLR), Facebook’s Prophet model, as well as the sub-epidemic wave (spatial-wave) and *n*-sub-epidemic modeling frameworks. We assess forecast performance using average mean squared error (MSE), mean absolute error (MAE), weighted interval score (WIS), 95% prediction interval coverage (95% PI coverage), and skill scores. Average Winkler scores were used to calculate skill scores for 95% PI coverage.

Overall, the *n*-sub-epidemic modeling framework outcompeted other models across most locations and forecasting horizons, with the unweighted ensemble model performing best across all forecasting horizons for most locations regarding average MSE, MAE, WIS, and 95% PI coverage. However, many locations had multiple models performing equally well for the average 95% PI coverage. The *n*-sub-epidemic and spatial-wave frameworks improved considerably in average MSE, MAE, and WIS, and Winkler scores (95% PI coverage) relative to the ARIMA model. Findings lend further support to sub-epidemic frameworks for short-term forecasting epidemics of emerging and re-emerging infectious diseases.

**Summary:** In the face of many unknowns (i.e., transmission, symptomology) posed by the unprecedented 2022-2023 mpox epidemic, near real-time short-term forecasts of the epidemic’s trajectory were essential in intervention implementation and guiding policy. As case levels continue to dissipate, evaluating the modeling strategies used in producing real-time forecasts is critical to refine and grow the field of epidemiological forecasting. Here, we systematically evaluate the performance of an ensemble *n*-sub-epidemic and related sub-epidemic wave (spatial-wave) modeling frameworks against ARIMA, GAM, Prophet, and SLR models in producing sequential retrospective weekly (1-4 week) forecasts of mpox cases for the highest burdened countries (i.e., Brazil, Canada, France, Germany, Spain, the United Kingdom, and the United States) and on a global scale. Overall, the *n*-sub-epidemic framework outperformed all other models most frequently, followed closely in success by the spatial-wave framework, GAM, and ARIMA models regarding average MSE, MAE, and WIS metrics. The *n*-sub-epidemic unweighted model and spatial-wave framework performed best overall based on average 95% PI coverage, and we noted widespread success for both frameworks in average Winkler scores. The considerable success seen with both frameworks highlights the continued utility of sub-epidemic methodologies in producing short-term forecasts and their potential application to other epidemiologically different diseases.

## Introduction

Over the past year, an unprecedented global surge in mpox cases has affected multiple countries previously free of the disease [1,2]. Early on, little was known regarding critical transmission and control factors – vaccination, the role of asymptomatic individuals, and transmission pathways – which shaped the overall epidemic trajectory [3,4]. For example, a majority of mpox cases in the current epidemic have occurred via close intimate and sexual contact, mainly in the gay, bisexual, and men who have sex with men (MSM) communities [5–10], whereas in past outbreaks and in endemic countries, this was not the case [7,11,12]. Therefore, as mpox cases increased in late July 2022, our team started to produce and make publicly available weekly short-term forecasts of new cases for the highest-affected countries (e.g., Brazil, Canada, France, Germany, Spain, the United Kingdom, and the United States) and at the global level employing semi-mechanistic growth models [13–16], which have shown past forecasting success in the context of emerging infectious disease [16–18].

For any emerging pathogen that rapidly transmits throughout a population, short-term forecasts of the epidemic’s trajectory at different spatial scales can help guide policy and intervention strategies [8,19–22]. However, there is little opportunity to assess forecasting performance and improve models amid an ongoing public health crisis. Fortunately, as of September 2022, cases have steadily declined worldwide [1,23–25], with non-endemic countries reporting 88,696 cases and 142 deaths overall as of October 11^th^, 2023 [26]. Therefore, as we continue into the declining phase of the epidemic, a retrospective evaluation of employed forecasting methodologies is vital to preparing for future public health events.

While over 108 countries experienced unprecedented levels of mpox cases, Brazil, Canada, France, Germany, Spain, the United Kingdom, and the United States saw the largest spike in confirmed cases during the initial wave of the epidemic [1,16]. Nevertheless, each country saw variable epidemic trajectories, with the United States facing the highest burden of cases, and Brazil experiencing the most prolonged peak (∼8-weeks)[1,26]. Canada, the United Kingdom, and France saw multiple subsequent smaller peaks during the declining phase of the outbreak, possibly due to a decrease in vaccine uptake [27,28], failure to receive the full vaccination regiment (i.e., two doses) [27,28], or relaxation of preventative behaviors [1,20,28] (Fig A in S1 Appendix). Given the heterogeneous impact of the epidemic at different spatial scales, a retrospective evaluation of multi-model forecasts is key for better understanding sub-epidemic model performance in different contexts, which can contribute to advancing the science of epidemic forecasting.

Several methodologies have been employed to forecast the case trajectory of the 2022-2023 mpox epidemic in various geographical regions, including, but not limited to, models focused on human judgment [29], deep learning, and artificial intelligence models (AI)[30–32], machine learning models [30–35], statistical models such as auto-regressive integrated moving average (ARIMA) models [31,33–37], compartmental models [38], and semi-mechanistic sub-epidemic models [16]. Performance metrics employed have mainly included variations of mean absolute error (MAE) [16,30,31,33–35,37], mean squared error (MSE) [16,30–37], and mean absolute percentage error (MAPE) [16,31,33–35], with only two studies also including probabilistic measures of performance such as the weighted interval score (WIS) [16,39]. Of the studies included here, four focused on the ascending phase of the epidemic (January 2022-Mid August 2022)[29,33,34,36], four evaluated forecasts of the ascending and peak of the epidemic (February 2022-September 2022) [30–32,39], and four focused on the ascending, peak and decline phases of the epidemic (January 2022-Present) [16,35,37,38].

In this paper, we carried out a comprehensive retrospective assessment of forecasting performance (1-4 weeks ahead) of sub-epidemic modeling frameworks compared against other commonly employed statistical models, including auto-regressive integrated moving average models (ARIMA), general additive models (GAM), simple linear regression (SLR), and Facebook’s Prophet model (Prophet) for the most impacted countries. We focus on producing and evaluating forecasts using the weeks of July 14th, 2022, through January 26^th^, 2023, for model calibration, which captures the full epidemic trajectory in the countries of interest. We considered a comprehensive set of performance metrics in the field of epidemic forecasting, namely the mean absolute error (MAE), mean squared error (MSE), 95% prediction interval coverage (95% PI), and weighted interval score (WIS).

## Methods

### Source Data

Our team utilized confirmed mpox cases from the highest-impacted countries (weeks of May 5th, 2022, through the week of January 26th, 2023) to produce and evaluate sequential retrospective weekly forecasts (weeks of July 14^th^, 2022, through the week of February 23rd, 2023) from the Centers for Disease Control (CDC) [40] and the Our World and Data (OWID) GitHub page [41]. We aggregated cases on the weekly level to ensure an adequate number of cases for the forecasting process, and to control for daily reporting differences between locations. Both data repositories employ different collection and reporting methodologies regarding their published data. For example, the CDC team obtains mpox data from the CDC call center or their emergency response common operating platform, DCIPHER, published weekly on Wednesdays but is now updated monthly [40]. The OWID team uses data assembled by the World Health Organization (WHO) from direct data reporting from WHO Member States and posts daily case updates [9,41].

Both the CDC and the OWID team define a *confirmed case* as a laboratory-confirmed case (i.e., infection detected by polymerase chain reaction testing) [42,43], but define the date associated with each case differently. The CDC defines the *onset date* as the earliest date available regarding the case (i.e., symptom onset, diagnosis date, positive laboratory test date, or date reported to the CDC)[40]. However, the OWID team defines *onset date* first as the date of symptom onset. If symptom onset is unknown, they then employ the date of lab or clinical diagnosis, followed by the date the case was reported [9].

### Data Collection and Preparation

We obtained daily cases series published weekly from the CDC [40] and daily from the OWID [41] team on August 9^th^, 2023, and August 15^th^, 2023, respectively. We then aggregated the data to the weekly level, with a week starting on Thursday and ending on Wednesday, to avoid calibrating the model with partial weeks of case data. Our team retrieved data for the United States from both the CDC and OWID sources to explore differences in model performance in the United States dependent on the data source used. Data on the global level and for the countries that experienced the most significant outbreaks during the initial epidemic wave (i.e., Brazil, Canada, France, Germany, Spain and the United Kingdom) were gathered only from the OWID team [1].

### Model calibration and forecasting strategy

Our analysis includes 12-models in total: the *n*-sub-epidemic framework (i.e., top-ranked, second-ranked, weighted ensemble, unweighted ensemble), the sub-epidemic wave (spatial wave) framework (i.e., top-ranked, second-ranked, weighted ensemble, unweighted ensemble), a generalized additive model (GAM), a simple linear regression model (SLR), an auto-regressive integrated moving average model (ARIMA), each with an assumption of normality, and Facebook’s Prophet model. Compared to a 9-week and 10-week calibration period, the 11-week calibration period produced the best forecast performance overall. Therefore, the remainder of this analysis employs an 11-week calibration period for all model fitting and forecasting. The full sensitivity analysis comparing calibration period forecasting performance can be found in supplementary file 2 (S2 Appendix).

For the week of July 14th, 2022, we could not produce forecasts for Canada for the *n*-sub-epidemic framework due to the low case counts during the early phase of the epidemic. We also could not produce forecasts for the week of July 14^th^, 2022, and July 21st, 2022, using the *n*-sub-epidemic and spatial-wave frameworks for Brazil due to low case counts. Similarly, some model calibration periods may be between 8 and 10 weeks long for the *n*-sub-epidemic and spatial-wave frameworks as initial zeros (i.e., any zeros that proceed the calibration period) are truncated until the first non-zero observation occurs.

Therefore, for each model and location, we conducted approximately 112 retrospective weekly sequential 1-to 4-week forecasts, with forecasts spanning from the weeks of July 21^st^, 2022, through the week of February 23^rd^, 2023. We primarily used 11-week calibration periods for each location, with the forecast period date representing the first day of the week in which the forecast was produced. Descriptions of the *n*-sub-epidemic and spatial-wave frameworks, ARIMA, GAM, and Prophet methodologies are below. The SLR model included *time* as the only predictor; therefore, its description is not included in further detail. Data posted in the week of August 3^rd^, 2023, was used to produce and evaluate all forecasts.

### The *n*-sub-epidemic modeling framework

The *n*-sub-epidemic framework employs multiple epidemic trajectories modeled as the aggregation of overlapping and asynchronous sub-epidemics [17]. A sub-epidemic follows the 3-parameter generalized-logistic growth model (GLM), which is given by the following:

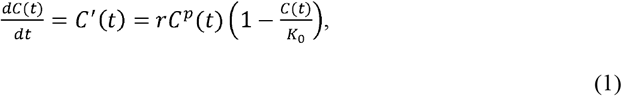

where *C* (*t*)denotes the cumulative mpox case count at time *t* and 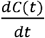 describes the incidence curve of mpox cases over time *t*. The parameter *r*, the growth rate per unit of time, remains positive, and parameter *k*_0_ represents the final outbreak size. The “scaling of growth” parameter *p* ∈ [0,1] allows the model to capture early sub-exponential and exponential growth patterns. If *p =*0, this equation describes a constant number of new cases over time, while *p =*1 indicates that the early growth phase is exponential. Intermediate values of *p* (0 *< p<*1) describe early sub-exponential (e.g., polynomial) growth dynamics. The GLM has displayed competitive performance in the context of varying infectious diseases, including Zika, Ebola, and COVID-19 [44–47].

An *n*-sub-epidemic trajectory is comprised of *n* overlapping sub-epidemics and is given by the following system of coupled differential equations:

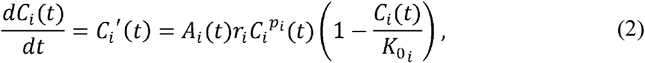

Where *C*_*i*_ (*t*) tracks the cumulative number of cases for sub-epidemic *i*, and the parameters that characterize the shape of the *i* _*th*_ sub-epidemic are given by 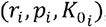, for *i =*1,…, *n*. Here, *n* represents the number of sub-epidemics considered in the epidemic’s trajectory. When *n =*1, the sub-epidemic model is equivalent to the GLM above (1). However, when *n>*1, we employ the indicator variable, *A*_*i*_ (*t*), to model the onset timing of the (*i* +1) _*th*_ sub-epidemic, where (*i* +1) ≤ *n*. Therefore, the (*i* +1) _*th*_ sub-epidemic is triggered when the cumulative curve of the *ith* sub-epidemic exceeds the case threshold value, *C*_*thr*_ (i.e., 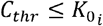). Thus, we have

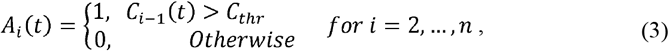

where *A*_*1*_(*t*) =1 for the first sub-epidemic. Therefore, 3*n*=1 parameters are needed to model an *n*-sub-epidemic trajectory for *n>*1. This analysis considers a maximum of two sub-epidemics in the *n*-sub-epidemic trajectory (*n ≤2*). The initial number of mpox cases is given by *C*_1_ (0) = *I*_0_, where *I*_0_ is the initial number of cases in the observed data. The cumulative curve of the *n*-sub-epidemic trajectory is given by:

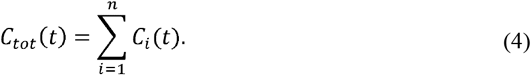

Overall, the modeling framework can be applied to diverse epidemic patterns including those characterized by multiple peaks and extended high-case level plateaus, similar to what has been observed throughout the mpox epidemic (Fig A in S1 Appendix).

### Parameter estimation for the *n*-sub-epidemic framework

Let the time series of new incident mpox cases used in model calibration be denoted as 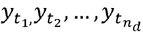 where *t*_*j*_,*j* = 1,2,…*n*_*d*_, are the time points for the time series data, and *n*_*d*_ is the number of observations. Using these case series, we estimated a total of 3 *n* +1 model parameters, namely 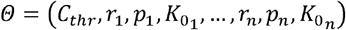 when *n*, the number of sub-epidemics, is greater than 1. Let *f* (*t, Θ*) denote the expected curve of new mpox cases of the epidemic’s trajectory. We employed the nonlinear least squares (LSQ) method to estimate the model parameters by fitting the model solution to the observed mpox data [48]. Specifically, we considered different values of *C*_*thr*_ by simply discretizing its range of plausible values. For each *C*_*thr*_, we searched for the set of parameters 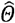 that minimized the sum of squared differences between the observed data 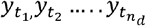 and the model mean *f* (*t, Θ*). We then selected the *C*_*thr*_ and other associated parameters based on which led to the smallest sum of squared errors. That is, 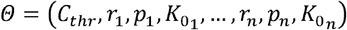 was estimated by 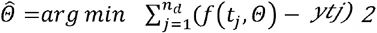 as. Finally, we selected the top-ranked sub-epidemic by the corrected Akaike Information Criterion (*AIC*_*c*_) values of the set of best-fit models based on one and two sub-epidemics discussed below.

### Parametric bootstrapping

We quantified parameter uncertainty using a bootstrapping approach described in ref [49] which allows the computation of standard errors and related statistics without closed-form solutions. To that end, we used the best-fit model, 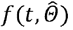, to generate *B*-times replicated simulated datasets of size *n*_*d*_, where the observation at time *t*_*j*_ is independently sampled from a normal distribution with mean 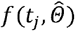 and variance 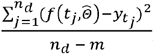 where *m* is the number of parameters with *m = 3* for 1 sub-epidemic (i.e., *n*=1) and *m = (3n)+1* for *n>*1. Then, we refit the model to each *B* simulated dataset to re-estimate each parameter. The new parameter estimates for each realization are denoted by 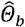 where *b* = 1,2,…,*B*. Using the sets of re-estimated parameters 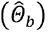 the empirical distribution of each estimate can be characterized, and the resulting uncertainty around the model fit was obtained from 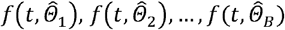. We also ran the calibrated model forward in time to generate short-term forecasts (i.e., 1 to 4 weeks ahead) with quantified uncertainty, employing the same parametric bootstrapping as above. We used 300 (*B =*300) bootstrap realizations for this analysis to characterize parameter and forecasting uncertainty.

### Selecting the top-ranked sub-epidemic models

We selected the top-ranked sub-epidemic models by the corrected Akaike Information Criterion (*AIC*_*c*_) values of the set of best-fit models based on one and two sub-epidemics. We ranked the models from best to worst according to their *AIC*_*c*_ values, which is given by [50,51]:

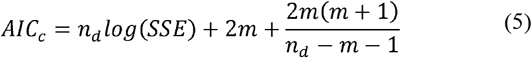

where 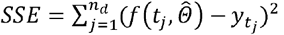,*m* is the number of model parameters, and *n*_*d*_ is the number of data points. Parameters from the above formula for *AIC*_*c*_ are estimated from the nonlinear least-squares fit, which implicitly assumes normal distribution for error. We selected the top two ranking sub-epidemic models, which were then employed to create an ensemble sub-epidemic model (i.e., Ensemble (2)) described in detail below.

### Constructing ensemble *n*-sub-epidemic models

We generated ensemble models from both the unweighted (equally weighted top-ranking models) and weighted combination of the two highest-ranking sub-epidemic models (i.e., top- and second-ranked) as deemed by the *AIC*_*ci*_ for the *i*-th ranked model where 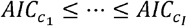 and *i* = 1, …, *I*. We compute the weight i for the *i* -th model, *i* = 1, …, *I*, as follows:

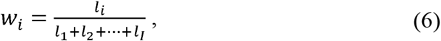

where *l*_*i*_ is the relative likelihood of model *i*, which is given by 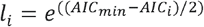 [52] and hence *w*_*I*_ ≤ … ≤ *w*_*1*_. For the unweighted model, we use the same weight *w*_*i*_ =1/*I* for all models, *i* = 1, …, *I*. The prediction intervals based on the ensemble model can be obtained using the bootstrap approach explained above.

### Sub-epidemic wave framework (Spatial wave framework)

The sub-epidemic wave, or spatial-wave modeling framework, aggregates linked, overlapping, synchronous sub-epidemics (“waves”) to capture complex disease trajectories of differing shapes and sizes [18,53]. Thus, the modeling framework is referred to as the “spatial wave” framework.

Similar to the *n*-sub-epidemic framework, this framework has shown past success in capturing diverse epidemic waves (i.e., high-level, prolonged epidemic plateaus and multi-peak trajectories), such as those seen throughout the mpox epidemic (Fig A in S1 Appendix) [18,53].

The mathematical building block for the spatial-wave model is the same 3-parameter GLM discussed above (1). However, a sub-epidemic wave consists of *n-*overlapping sub-epidemics employing the following system of coupled differential equations:

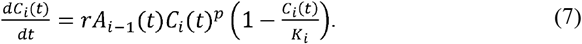

The incidence curve of mpox cases is given by 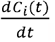, where *C*_*i*_ (*t*)is the cumulative number of cases for sub-epidemic *i* at time *t*, and *K*_*i*_ is the size of sub-epidemic *i*, where *i=* 1,…,*n*. The indicator parameter *A*_*i*_ (*t*)is the same as given in (3). Starting from an initial sub-epidemic size *K*_*0*,_ the size of subsequent sub-epidemics *K*_*i*_, decline at rate *q* following either an exponential or power-law function as described below. The growth rate per unit of time is represented by *r*, and the “scaling of growth” parameter by *p*. Unlike the equation given in (2), parameters *p* and *r* do not vary across sub-epidemics, and the value of *K*_*i*_ depends on the decline rate *q*. Hence, a total of 5 parameters (*r,p,C*_*thr*_,*q, K*_*0*_) for *i* =1,…,*n* are needed to characterize a sub-epidemic wave composed of two or more sub-epidemics (*n*>1), and four are needed when *n* = 1. This analysis considers a maximum of five sub-epidemics (*n* ≤5) in the epidemic wave trajectory.

### Sub-epidemic decline

In this framework, the size of the subsequent *i*_*th*_ sub-epidemic (*K*_*i*_) remains steady or declines in response to external factors (i.e., behavior changes or interventions). We considered both exponential and inverse (power-law) decline functions to model the size of consecutive sub-epidemics as described below, where the decline function that yields the best model fit is employed for each forecast period and location.

### Exponential decline of sub-epidemic sizes

When consecutive sub-epidemics follow exponential decline, *K*_*i*_ is given by:

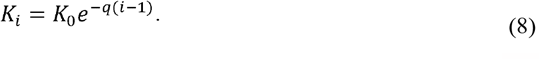

The parameter *K*_*0*_ is the size of the initial sub-epidemic, and *K*_*1*_= *K*_*0*_. If *q* = 0, then the model predicts an epidemic wave composed of sub-epidemics of the same size. When *q* > 0, the epidemic wave is composed of a finite number of sub-epidemics given by *n*_*tot*_ which is a function of *C*_*thr*_, *q*, and *K*_*0*_:

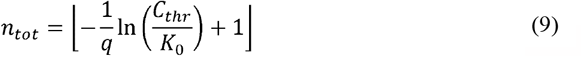

where the brackets ⌊ *⌋ denote the largest integer that is smaller than or equal to *. The total size of the epidemic wave composed of *n*_*tot*_ overlapping sub-epidemics has the following closed-form solution:

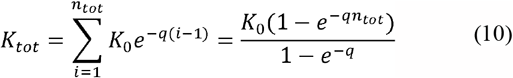

### Inverse (Power-law) decline of sub-epidemic sizes

When consecutive sub-epidemics decline according to the inverse function, *K*_*i*_ is given by:

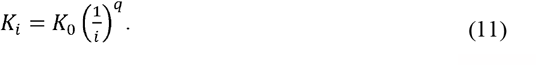

When q > 0, then the total number of sub-epidemics (*n*_*tot*_) comprising the epidemic wave is finite and given by:

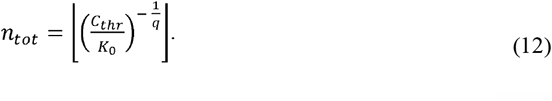

The total size of an epidemic wave is given by the aggregation of *n*_*tot*_ overlapping sub -epidemics:

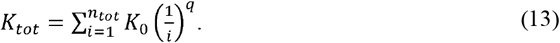

### Selecting the top-ranked sub-epidemic models

We examined the *AIC*_*c*_ values of the set of best-fit sub-epidemic wave models with different values of the threshold parameter, *C* _*thr*_, to select the top two ranking sub-epidemic models. The *AIC*_*c*_ is given by (5), where *m*=5 when the number of sub-epidemics is greater than one (n>1) and *m*=4 when working with a single sub-epidemic (*n*=1) The *AIC*_*c*_ given in (5) for the parameter estimation assumes nonlinear least-squares fit, which implicitly assumes normal distribution for error.

Parameter estimation and bootstrapping, construction of the ensemble models, and forecasting methodologies follow the same procedures explained for the *n*-sub-epidemic model.

### Auto-Regressive Integrated Moving Average Models (ARIMA)

The auto regressive integrate moving average (ARIMA) model is a tool for understanding and predicting future values in a time series. It is commonly employed in forecasting financial [54– 56] and weather [57–59] trends, and have become a standard benchmark in disease forecasting [17,31,33–37,60,61]. ARIMA models consist of three parts: the auto-regression (AR) part involving regressing on the most recent values of the series, the moving average (MA) of error terms occurring contemporaneously and at previous times, and the integration (I) or differencing to account for the overall trend in the data and to make the time series stable. Without the differencing part, the ARMA (p, q) model

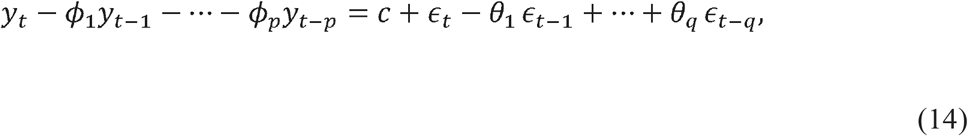

consists of the AR part of regressing *yt* on the lagged values *y*_*t*−*1*,_ … *y*_*t* −*p*_, and the MA part of the linear combination of errors *ϵ* _*t*_, *ϵ* _*t* −*1*_, *… ϵ* _*t* −*q*_, where {*ϵ* _*t*_} is a white noise process with mean 0 and variance *σ*^2^, and c is a constant. The ARMA model (14) can be used to describe stationary stochastic process whose joint probability distribution does not change with time. When data show evidence of non-stationary and the mean changes over time (i.e., trend), a differencing step (corresponding to the “integrated” [62] part of the model) can be applied one or more times to eliminate the trend. To difference the data, the difference between consecutive observations is computed, such as *y*_*t*_ − *y*_*t* −*1*_ and then the ARMA model can be applied to the data after differencing. This leads to an ARIMA model. Mathematically, an ARIMA (p, d, q) process is given by:

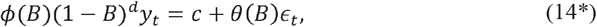

where B denotes the backshift operator implying *By*_*t*_ = *y*_*t* −*1*_ and (*By*_*t*_)= *B* (*By*_*t*_)= *B*^*2*^*y*_*t*_ =*y*_*t* −*2*_, etc, *ϕ* (B) =1− *ϕ* _1_ B − … − *ϕ* _*p*_ *B*^*p*^ and p is the order of the AR model, d is the degree of differencing *θ* (B) =1− *θ* _1_ *B* − … − *θ* _*q*_ *B*^*p*^ and the q is the order of the MA model. With this notation, *ϕ* (B) *y*_*t*_ = *y*_*t*_ − *ϕ* _1_ *y*_*t* −*1*_ − … − *ϕ*_*p*_ *y*_*t* −*p*_, *θ* (*B*) *ϵ* _*t*_ =*ϵ* _*t*_ − *θ* _1_*ϵ* _*t* − *1*_ +… +*θ* _*q*_*ϵ* _*t* −*q*_ and (1−*B)* ^*d*^ means conducting the differencing d times. The *auto*.*arima* function in the R package “forecast” was used to select orders and build the model [63], and the *forecast* function in the R package “forecast” was used for forecasting [64]. Any negative predicted values were truncated at zero.

### Generalized Additive Models (GAM)

Generalized Additive Models (GAM) are an extension of Generalized Linear Models (GLM) models by including a sum of unknown smooth functions of some covariates [65]. They can capture non-linear trends, while maintaining similar levels of model explainability and simplicity seen with GLMs [66]. Specific to our study with time as the only covariate, assuming that normal distribution, our GAM is given as [66]:

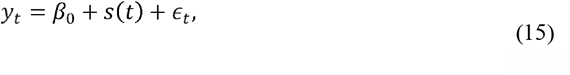

where *s*(·) is an unknown smooth function of time, and ϵ_*t*_ ∼ *N* (0, *σ*^2^). We used the *gam* function in the R package “mgcv” to fit this model [67], where the smooth function *s*(·) using basis functions, which are building blocks for creating more complex functions via linear combination. The default setting in the “mgcv” uses basis splines, which are piecewise polynomial functions. Specifically,

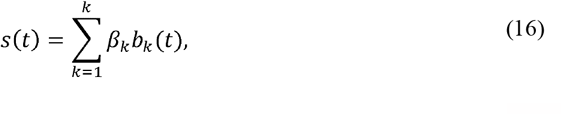

where{*b*_*k*_ (·)} represent the basis functions, {β_*k*_} are the expansion coefficients to be estimated, and *k* is the number of basis functions [66]. Here, the number of basis functions varied depending on the length of calibration data available for a given forecasting period. A discrete penalty was imposed on the basis coefficients to control the degree of smoothness, and the model was fitted by solving a penalized least square problem. The generalized cross-validation (GCV) criterion selected the smoothness tuning parameter. A more detailed description of the model fitting methodology can be found in [67], and the associated *predict* function was used for forecasting [68]. Any negative predicted values were truncated at zero.

### Facebook’s Prophet Model

Facebook’s Prophet model [69], initially designed for business-related forecasting, has recently been applied more frequently across multiple fields, including infectious disease modeling. Specifically, the model has produced both COVID-19 [60,70,71] and mpox forecasts [31]. The model’s primary assumption is that it is “decomposable”, therefore, *y*(*t*), the main trend, can be decomposed into three pieces plus an error term. Thus, the model has the following form:

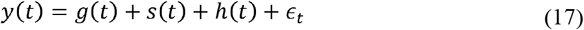

where *g*(*t*) is a non-periodic component used for modeling of the overall trend, *s*(*t*) is a periodic component used for modeling of periodic (e.g., monthly, weekly or other seasonal) changes over time and *h*(*t*) is an irregular events component used for modeling of irregular changes (e.g. holidays or similar events). The component □_*t*_ is an error of the model at time *t*.

We used the default setting of the R *prophet* function from the “prophet” package where the model has been implemented as given in [72]. Therefore, we fit the default fits of the model and assess only the model’s forecasting performance. The *predict* function was used for forecasting from the model fit [68]. Any negative predicted values were truncated at zero. A more detailed description of the model fitting methodology can be found in [72].

### Model evaluation

Forecast performance was evaluated for all the retrospectively generated 1-4 weeks ahead forecasts utilizing data downloaded the week of August 3^rd^, 2023. Descriptions of the mean absolute error (MAE), mean squared error (MSE), 95% prediction interval coverage (95% PI) and weighted interval score (WIS) metrics, along with Winkler and skill score calculations used in the evaluation, can be found below.

### Performance Metrics

To assess performance, we used the mean absolute error (MAE), the mean squared error (MSE), the coverage of the 95% prediction interval (95% PI coverage), and the weighted interval score (WIS). A more detailed description of the performance metrics can be found in [17].

MAE and MSE were used in assessing deviations of the mean model fit to the observed case data [73,74]:

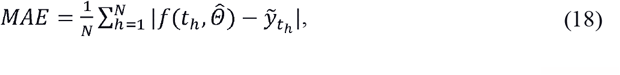

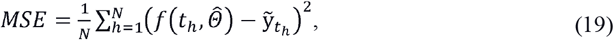

where 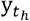 represents the originally observed case counts across each study location of the *h*-time units ahead forecasts, and *t*_*h*_ denotes the time points for the sample data [73,74].

Model uncertainty was assessed via the 95% prediction interval coverage and WIS. Here, the 95% PI coverage represents the proportion of observed cases that fall within the 95% PI. The WIS proves quantiles of the predictive forecast distribution by combining a set of interval scores (IS) from probabilistic forecasts. Only a central (1−_α_)×100% PI is needed to calculate an IS [73]. See below:

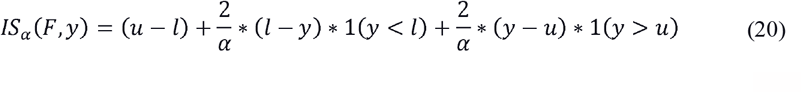

The indicator function, represented above by **1**, refers to the fact that **1**(*y* < *l*) = 1 if *y* < *l* and 0 otherwise. The 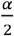 and 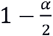 quantiles of the forecast *F* are represented by *l* and *u*. The IS consists of three distinct quantities:

1. The sharpness of *F*, given by the width *u* − *l* of the central (1 − α) 100% PI.
2. A penalty term 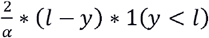 for the observations that fall below the lower end point *l* of the (1 − α) 100% PI. This penalty term is directly proportional to the distance between *y* and the lower end *l* of the PI. The strength of the penalty depends on the level.
3. An analogous penalty term 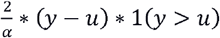 for the observations falling above the upper limit *u* of the PI.

We report several central PIs at different levels (1 − α_1_)< (1 − α_2_)< … <(1 − α_*K*_) along with the predictive median, *m*, which can be seen as a central prediction interval at level 1 − α_0_ ⟶ 0. This is referred to as the WIS, and it can be evaluated as follows:

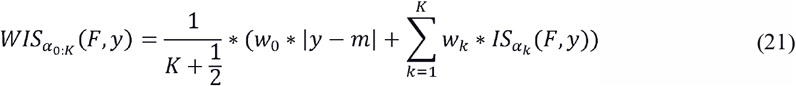

where, 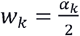 for *k* = 1,2, …. *K* and 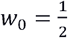. Hence, WIS can be interpreted as a measure of how close the entire distribution is to the observation in units on the scale of the observed data [75,76].

### Skill Scores and Winkler Score

We calculated skill scores to compare the proportion of improvement over the established ARIMA model (baseline model) for average MSE, MAE, and WIS, using the *n*-sub-epidemic and spatial-wave frameworks as comparison models. Here, *average* refers to the average metric over all forecasting periods for each model, location, and forecasting horizon. The formula is as follows [77]:

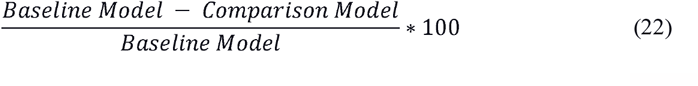

For 95% PI coverage, we first calculated Winkler scores to allow for the quantification of the proportion improvement over the ARIMA model, using the *n*-sub-epidemic and spatial-wave frameworks as comparison frameworks. Winkler scores were calculated as follows [77]:

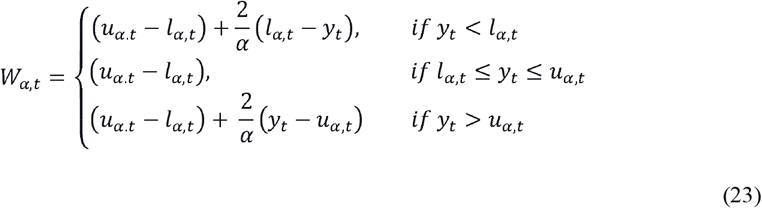

where *u* _*α,t*_ is the upper bound of the 95% PI interval at time *t, l*_*α,t*_ is the lower bound at time *t, y*_*t*_ is the observed mpox case incidence at *t*, and α =0.05 as we are working with 95% PI intervals. We then calculated the skill scores (22) for average Winkler scores, where scores are averaged across all forecasting periods, for each model, location, and forecasting horizon.

We selected the ARIMA model as baseline, as it has been frequently evaluated against other forecasting methodologies in the context of mpox [31,33–37]. Therefore, its inclusion in skill score calculations provides a more in-depth quantitative evaluation of the forecasting abilities of the *n*-sub-epidemic and spatial-wave frameworks against a well-vetted methodology.

R-studio (Version: *4*.*3*.*1 (Beagle Scouts)*) and MATLAB (Version: *R2022a*) were used to produce and evaluate all forecasts. Forecast period specific performance metrics, associated forecasts, Winkler scores, and all software scripts used to produce and evaluate forecasts will be available in a public repository upon journal acceptance.

## Results

### Overall and time-period specific forecast performance

The *n*-sub-epidemic framework outperformed the established statistical (i.e., ARIMA, GAM, SLR, Prophet) and spatial-wave framework models across most locations. Specifically, the *n*-sub-epidemic unweighted ensemble was the most successful in Brazil, the United Kingdom, the US (OWID), and the World. The model was followed in success by the top-ranked (Brazil) and weighted ensemble (Canada and Spain) models. Nevertheless, the spatial-wave top-ranked model performed best overall in Germany, the ARIMA model in France, and the GAM in the US(CDC). More detailed descriptions of location- and metric-specific performance can be found below.

When looking at model performance across countries during specific epidemic phases (i.e., ascending, peak, descending, tail-end), the GAM model performed best overall during the ascending phase of the epidemic regarding average MSE, MAE, and WIS. However, the *n*-sub-epidemic unweighted ensemble and second-ranked models performed best most frequently regarding average 95% PI coverage. As the epidemic peaked and then declined in countries, the *n*-sub-epidemic framework performed best across most forecasting metrics. However, the spatial-wave top- and second-ranked models produced the lowest average WIS during the declining phase most frequently. Finally, as the epidemic settled at low case levels, the spatial-wave top-ranked model did best, producing the lowest average MSE, MAE, and WIS most often. The ARIMA model produced the best average 95% PI coverage during the same time frame. Overall, model performance improved as case levels declined. Supplementary figures B-E include visualizations of average forecast performance trends across epidemic phases for each country and performance metric (S1 Appendix).

### Country-level performance

#### Brazil

The *n*-sub-epidemic framework, specifically the top-ranked and unweighted ensemble models, performed best overall in Brazil, outperforming other models 44% of the time across performance metrics and horizons (Figs 1-4). The models were followed closely in success by the *n*-sub-epidemic weighted ensemble model (38%) (Figs 1-4). The *n*-sub-epidemic top-ranked and weighted ensemble models produced the lowest average MSE (Range: 16,302.0-128,915.5) and MAE (Range: 86.4-135.0) across most forecasting horizons (Figs 1-2). Both models improved between 8.5 and 26.8% over the ARIMA model regarding average MSE and 21.0-33.2% for average MAE. The *n*-sub-epidemic top-ranked model most frequently produced the lowest average WIS (Range: 57.4-97.5), improving 23.0-30.2% over the ARIMA model (Fig 3). The *n*-sub-epidemic unweighted model produced the lowest average WIS (56.4), MSE (14,240.9), and MAE (85.2) during the 1-week forecasting horizon and performed best across all forecasting horizons regarding 95% PI coverage (Range: 93.8-96.3%) (Figs 1-4). When comparing average Winkler scores, the *n*-sub-epidemic unweighted ensemble improved 6.3-44.6% over the ARIMA model. Tables A1 through A5 in S3 Appendix contain the tabulation of skill scores and performance metrics across epidemic phases, models, and forecasting horizons for Brazil.

**Fig 1.**
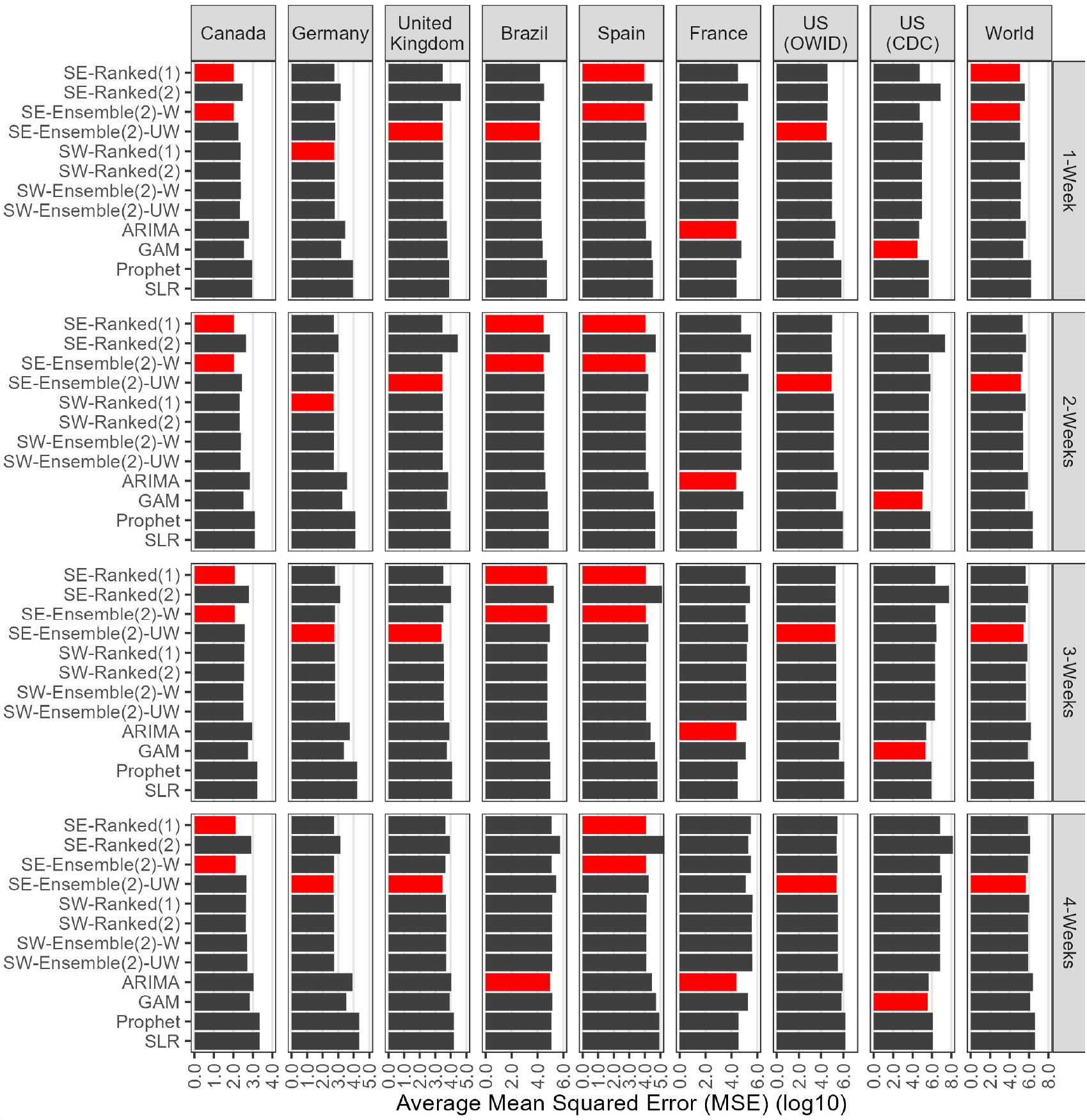
Average Mean Squared Errors (MSE) for each location, model, and forecasting horizon (weeks of July 21st, 2022-Febraruy 23rd, 2023). A total of 1,008 forecasts were produced for each model. The graph is shown on the log10 scale. The red bars indicate the best-performing model at a given location and horizon, and multiple red bars indicate ties between models. The *n*-sub-epidemic framework performed best most frequently across forecasting horizons and locations regarding average MSE.

#### Canada

The *n*-sub-epidemic weighted ensemble and top-ranked models performed best overall in Canada, outperforming other models across forecasting horizons and metrics 69% and 56% of the time, respectively (Figs 1-4). Across all forecasting horizons for average MSE (Range: 103.2-128.9) and MAE (Range: 6.3-6.6), the *n*-sub-epidemic top-ranked and weighted ensemble models produced the lowest values (Figs 1-2). Both models improved considerably over the ARIMA model regarding average MSE (Range: 83.6-88.2%) and MAE (Range: 58.8-67.6%). The success of the *n*-sub-epidemic weighted ensemble model continued for average WIS, as the model produced the lowest WIS across most forecasting horizons (Range: 4.3-4.5) and improved between 54.3 and 63.0% over the ARIMA model (Fig 3). However, the ARIMA model performed best most frequently regarding average 95% PI coverage (Range: 94.25-96.55%). Tables B1 through B5 in S3 Appendix contain the tabulation of skill scores and performance metrics across epidemic phases, models, and forecasting horizons for Canada.

#### France

Unlike the other countries, the ARIMA model outperformed all other models 94% of the time across forecasting metrics and horizons, followed by the *n*-sub-epidemic top-ranked (6%) and weighted ensemble (6%) models (Figs 1-4). The ARIMA model produced the lowest average MSE (Range: 22,986.7-24,413.1), WIS (Range: 61.1-62.3), and closest to 95% PI coverage on average (Range: 89.7-93.1%) consistently across forecasting horizons (Figs 1-2, 4). Regarding average MAE, the ARIMA model performed best across most forecasting horizons (Range: 90.5-93.8), albeit the 1-week horizon where the *n*-sub-epidemic top-ranked (80.9) and weighted ensemble performed best (80.9)(Fig 2). However, the *n*-sub-epidemic top-ranked and weighted ensemble models saw minimal improvement over the ARIMA regarding average MAE (<10%). Tables C1 through C5 in S3 Appendix contain the tabulation of skill scores and performance metrics across epidemic phases, models, and forecasting horizons for France.

**Fig 2.**
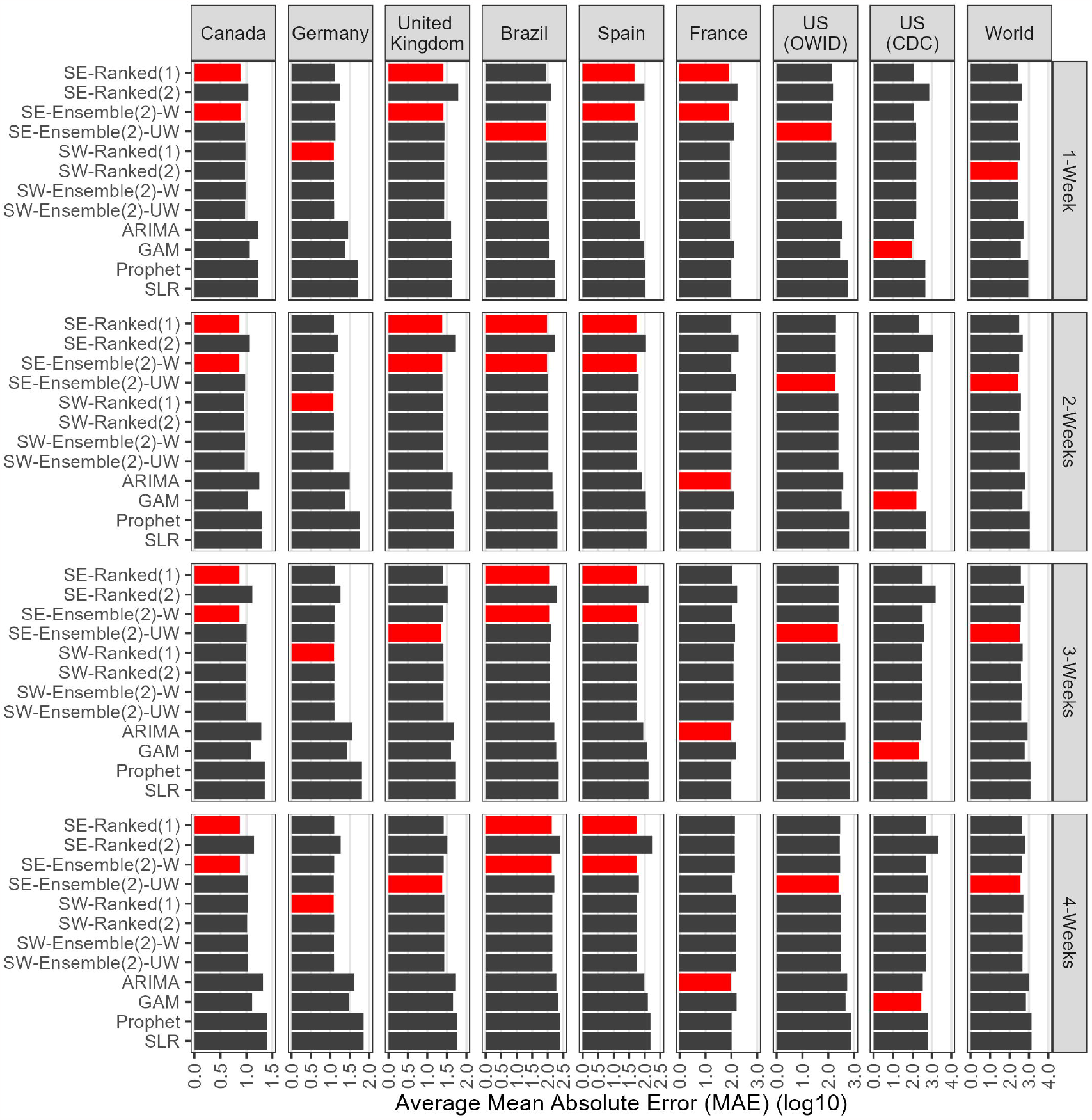
Average Mean Absolute Errors (MAE) for each location, model, and forecasting horizon (weeks of July 21st, 2022-Febraruy 23rd, 2023). A total of 1,008 forecasts were produced for each model. The graph is shown on the log10 scale. The red bars indicate the best-performing model at a given location and horizon, and multiple red bars indicate ties between models. The *n*-sub-epidemic framework performed best most frequently across forecasting horizons and locations regarding average MAE.

#### Germany

The spatial wave top-ranked model outperformed the other models 69% of the time across forecasting horizons and performance metrics, followed by the *n*-sub-epidemic (38%) unweighted ensemble model (Figs 1-4). The spatial-wave top-ranked model produced the lowest average MSE in the one (559.9) and two-week (514.2) forecasting horizons and across all forecasting horizons regarding average MAE (Range: 11.0-11.4) and WIS (Range: 7.9-8.3) (Figs 1-3). We also noted considerable improvement in average MSE (Range: 80.0-86.3%), MAE (Range: 59.1-72.7%), and WIS (Range: 55.1-70.8%) over the ARIMA model. The *n*-sub-epidemic unweighted ensemble produced the lowest average MSE during the three (563.06) and four-week (504.64) forecasting horizons, improving 90.1-94.0% over the ARIMA model (Fig 1). Similarly, the *n*-sub-epidemic unweighted ensemble performed best most frequently regarding average 95% PI coverage (89.7%), improving 42.8-74.4% over the ARIMA model in average Winkler scores (Fig 4). Tables D1 through D5 in S3 Appendix contain the tabulation of skill scores and performance metrics across epidemic phases, models, and forecasting horizons for Germany.

**Fig 3.**
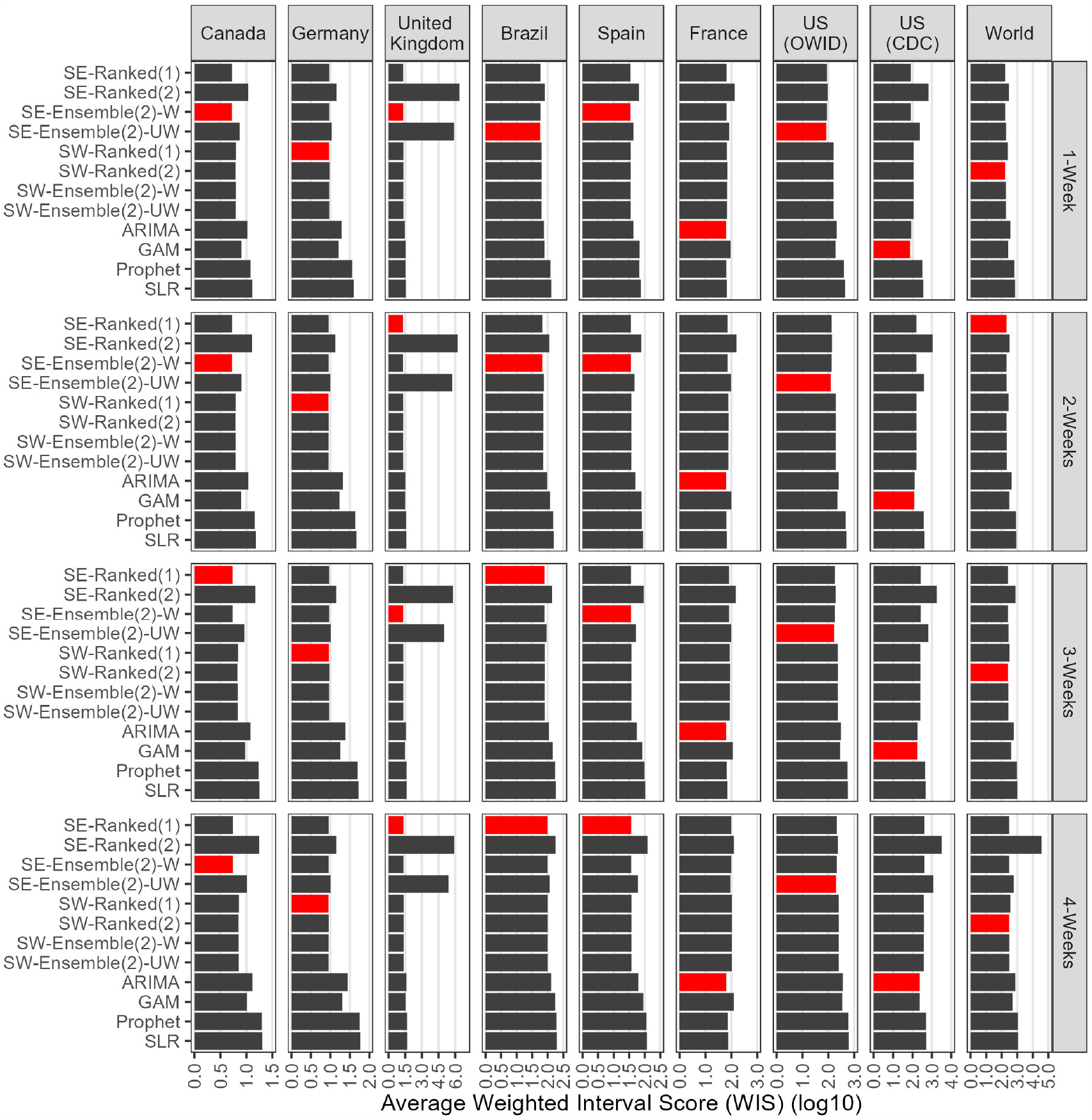
Weighted Interval Score (WIS) for each location, model, and forecasting horizon (weeks of July 21st, 2022-Febraruy 23rd, 2023). A total of 1,008 forecasts were produced for each model. The graph is shown on the log10 scale. The red bars indicate the best-performing model at a given location and horizon, and multiple red bars indicate ties between models. The *n*-sub-epidemic framework performed best most frequently across forecasting horizons and locations regarding average WIS.

**Fig 4.**
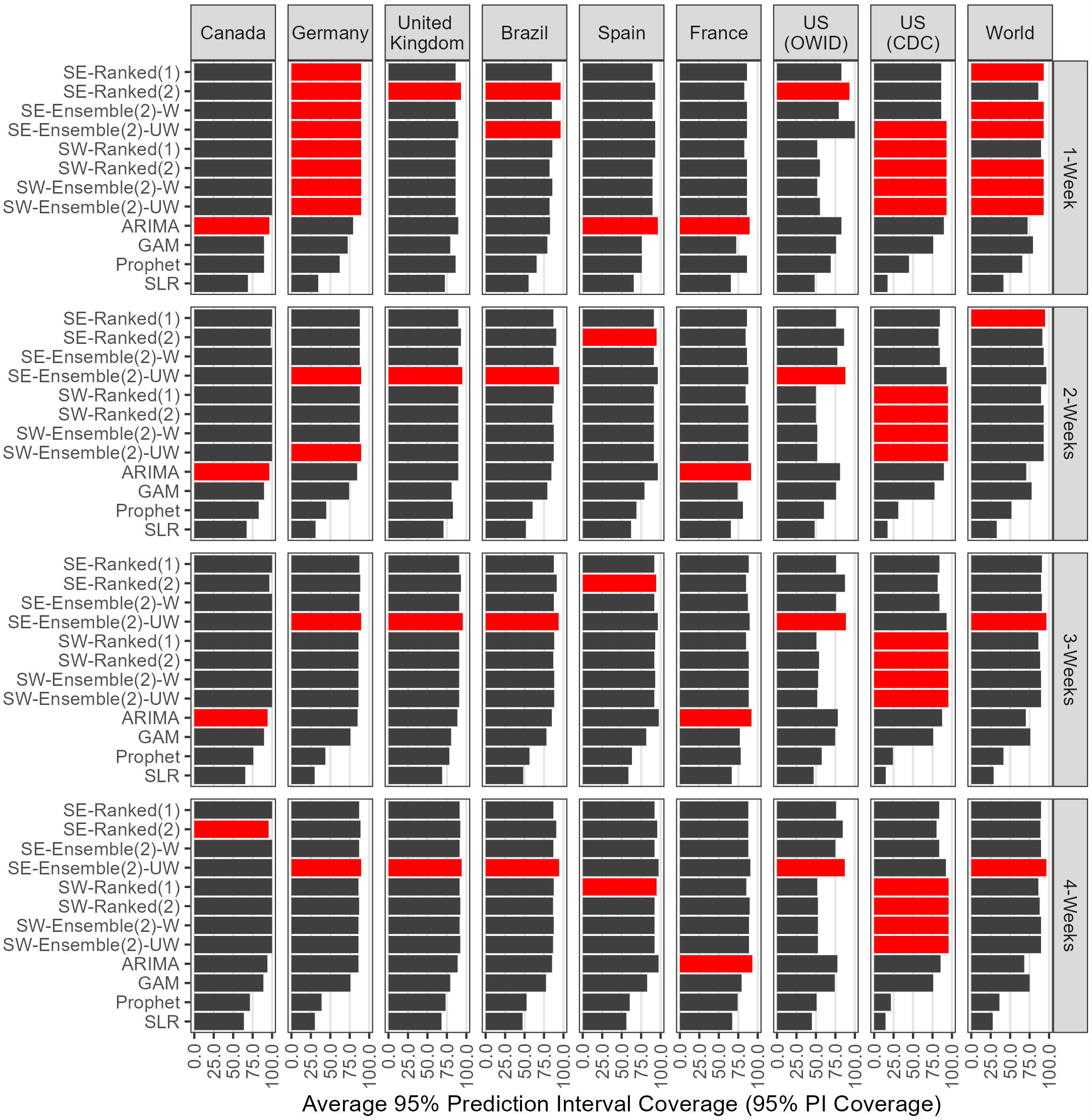
Average 95% Prediction Interval Coverage (95% PI coverage) for each location, model, and forecasting horizon (weeks of July 21st, 2022-Febraruy 23rd, 2023). A total of 1,008 forecasts were produced for each model. The red bars indicate the best-performing model at a given location and horizon, and multiple red bars indicate ties between models. The *n*-sub-epidemic unweighted ensemble performed best most frequently across forecasting horizons and locations regarding average 95% PI coverage.

#### Spain

The *n*-sub-epidemic framework performed superior in Spain, with the most success noted for the weighted ensemble model (69%) across forecasting periods and metrics, followed by the top-ranked model (63%) (Figs 1-4). Regarding average MSE (Range: 9020.3-12,286.1) and MAE (Range: 46.3-52.4), the *n*-sub-epidemic top-ranked and weighted ensemble models produced the lowest values across all forecasting horizons (Figs 1-2). Similarly, both models improved considerably over the ARIMA model regarding average MSE (Range: 25.7-57.4%) and MAE (Range: 33.2-45.9%). The *n*-sub-epidemic weighted ensemble produced the lowest average WIS (Range: 33.7-35.4) most frequently across forecasting horizons, improving 19.9-41.6% over the ARIMA model (Fig 3). The *n*-sub-epidemic second-ranked model performed well regarding average 95% coverage, producing the closest to 95% coverage across most forecasting horizons (Range: 94.3-94.8%) (Fig 4). However, the ARIMA model outperformed the *n*-sub-epidemic second-ranked model across all forecasting horizons regarding Winkler scores. Tables E1 through E5 in S3 Appendix contain the tabulation of skill scores and performance metrics across epidemic phases, models, and forecasting horizons for Spain.

#### United Kingdom

Overall, the *n*-sub-epidemic unweighted ensemble model outperformed other models 56% of the time across forecasting horizons and metrics, followed in success by the top-ranked (25%) and weighted (25%) ensemble models (Figs 1-4). The *n*-sub-epidemic unweighted ensemble model produced the lowest average MSE (Range: 2454.9-3012.8) across all forecasting horizons and for the three-(21.5) and four-week (22.8) horizons regarding average MAE (Figs 1-2). However, the model saw considerable success over the ARIMA model across all horizons in average MSE (Range: 45.9-72.1%) and average MAE (Range: 32.1-56.6%). The *n*-sub-epidemic top-ranked and weighted ensemble produced the lowest average MAE in the one (24.7) and two-week (22.9) horizons and saw split success across forecasting horizons regarding average WIS (Figs 2-3).

The *n*-sub-epidemic unweighted ensemble performed superior across most forecasting horizons for average 95% coverage (Fig 4). Tables F1 through F5 in S3 Appendix contain the tabulation of skill scores and performance metrics across epidemic phases, models, and forecasting horizons for the United Kingdom.

#### United States (CDC)

Unlike the other countries, the GAM model performed best in the US (CDC), outperforming the other models 69% of the time, followed by all models of the spatial-wave framework (25%) across forecasting horizons and metrics (Figs 1-4). The GAM produced the lowest average MSE (Range: 32,070.5-352,246.3) and MAE (Range: 95.0-291.3) across all forecasting horizons and the one-week through three-week forecasting horizons for average WIS (Range: 74.9-179.0) (Figs 1-3). However, all spatial-wave framework models performed superior across all forecasting horizons for average 95% PI coverage (Range: 93.1-95.7%), albeit the one-week horizon where the *n*-sub-epidemic unweighted ensemble model performed equally well (93.1%) (Fig 4). However, the ARIMA model outperformed the spatial-wave framework and the *n*-sub-epidemic unweighted ensemble model across all forecasting horizons regarding average Winkler scores. Tables G1 through G5 in S3 Appendix contain the tabulation of skill scores and performance metrics across epidemic phases, models, and forecasting horizons for the US (CDC).

#### United States (OWID)

The *n*-sub-epidemic unweighted ensemble model performed best across all forecasting horizons regarding average MSE, MAE, and WIS and across most horizons regarding average 95% PI coverage, outperforming other models 94% of the time (Figs 1-4). Regarding average MSE, the model produced values ranging from 31,944.2 to 246,467.4, a 66.0-82.9% improvement over the ARIMA model (Fig 1). We also observed considerable improvements over the ARIMA model in average MAE (Range: 50.6-60.9%) and WIS (Range: 45.4-60.8%). The *n*-sub-epidemic unweighted ensemble model produced superior average 95% PI coverage across most forecasting horizons (Range: 87.1-88.5%), improving 27.9-70.5% over the ARIMA model in average Winkler scores (Fig 4). Tables H1 through H5 in S3 Appendix contain the tabulation of skill scores and performance metrics across epidemic phases, models, and forecasting horizons for the US (OWID).

#### World

The *n*-sub-epidemic unweighted ensemble model, followed by the spatial-wave second-ranked model, outperformed other models across forecasting horizons and metrics 56% and 31% of the time, respectively (Figs 1-4). The *n*-sub-epidemic unweighted ensemble model produced the lowest average MSE (Range: 147,046.3-435,707.3) and MAE (Range: 286.3-362.3) across the two through four-week forecasting horizons, improving considerably over the ARIMA model (Figs 1-2). Similarly, the model performed superior regarding average 95% PI coverage most frequently across forecasting horizons (Range: 93.1-96.6%), albeit the two-week forecasting horizon where the *n*-sub-epidemic top-ranked model performed best (94.8%) (Fig 4). The *n*-sub-epidemic unweighted ensemble model improved 37.8-49.3% over the ARIMA model regarding average Winkler scores. However, the spatial-wave second-ranked model performed best across most forecasting horizons regarding average WIS (Range: 166.5-297.6), improving 53.5-59.7% over the ARIMA model (Fig 3). Tables I1 through I5 in S3 Appendix contain the tabulation of skill scores and performance metrics across epidemic phases, models, and forecasting horizons for the World.

## Discussion

In the context of the global emergency posed by the 2022-2023 mpox epidemic, we have systematically investigated short-term forecasting performance of multiple models that have shown competitive performance during epidemics and pandemics and rely on minimal data of the epidemic’s trajectory [16–18]. Overall, the *n*-sub-epidemic framework, specifically the unweighted ensemble model, followed by the spatial-wave framework, ARIMA model, and the GAM, performed best most frequently across locations and forecasting horizons compared to the other established modeling techniques (e.g., SLR, Prophet).

The *n*-sub-epidemic framework outperformed other models in Brazil, Canada, Spain, the United Kingdom, US (OWID) and the World, whereas the spatial-wave framework did best in Germany. However, France and the US (CDC) behaved uniquely compared to other countries with the ARIMA model and GAM, respectively, performing best across forecasting horizons. Except in France and the US (CDC), the spatial-wave and *n*-sub-epidemic models frequently improved considerably in each metric, including in average Winkler scores, over the ARIMA model.

Overall, our findings further support the sub-epidemics and ensemble frameworks for forecasting emerging infectious diseases, especially in the face of limited epidemiological data [17,18].

Although each included location experienced unique epidemic trajectories throughout the mpox epidemic, the consistent success of the *n*-sub-epidemic and spatial-wave frameworks highlights the utility of the aggregated sub-epidemic approach in capturing and producing short-term forecasts for a wide variety of epidemic trends. For example, the epidemic trajectories of the mpox epidemic in Canada, the United Kingdom, and the US (OWID) differ considerably.(Fig A in S1 Appendix). Canada sees multiple peaks in cases throughout the ascending and declining phases of the epidemic. In contrast, the United Kingdom sees a relatively smooth ascending phase with a prominent peak during the descending phase, and the US (OWID) sees a mostly smooth epidemic trajectory without drastic peaks during the ascending or descending phases.

Although different, the *n*-sub-epidemic unweighted ensemble model performed best across forecasting horizons for all three locations. Similar observations hold for Germany, Brazil, Spain, and the World.

France and the US (CDC) behaved differently than the other locations, with the ARIMA performing best in France and the GAM in the US (CDC). The mpox epidemic in France did not follow a uni-modal epidemic disease trajectory as seen in other study areas; instead, they experienced two prominent case peaks with slow ascending and descending phases (Fig A in S1 Appendix). However, throughout the outbreak, large stretches of the epidemic trajectory appear to form linear trends (i.e., ascending, between peaks, tail-end). As the ARIMA model inherently assumes linearity, the linear-like trajectory of mpox in France may explain the success of the ARIMA model in capturing and forecasting the disease’s trajectory [78]. Unlike France, the US (CDC) had little-to-no week-to-week fluctuation, forming a smooth, uni-modal epidemic trajectory (Fig A in S1 Appendix). The GAM, which performed best in the US (CDC), captures common non-linear trends, such as in the US (CDC) epidemic trajectory [66]. Therefore, similar to France, epidemic data following distinctive trajectories may explain the success seen by the ARIMA and GAM models in France and the US (CDC).

Model performance also differed when looking at specific epidemic phases (i.e., ascending, peak, descending, and tail-end of the epidemic). For example, across locations, the established statistical models outperformed other models most frequently during the ascending phase of the epidemic. Similar to France, the ascending phase for most locations was linear. Therefore, as the included established statistical techniques are inherently linear, they may best capture the linear-like trends. However, there were multiple epidemic phases with few forecasting periods available, limiting the ability to examine country-specific epidemic phase performance in greater detail.

Finally, the significant success of the *n*-sub-epidemic unweighted and weighted ensemble models highlights the utility of ensemble modeling in short-term forecasting [79–87]. Ensemble modeling has also shown considerable success against individual top-ranking sub-epidemic and other statistical models in past mpox short-term forecasting efforts [14,16], along with that of COVID-19 [17,76,84], seasonal influenza [82,87], and Zika [88]. Each ensemble model (e.g., weighted, and unweighted) included within the analysis performed competitively at various forecasting horizons and for different areas. However, the unweighted ensemble *n*-sub-epidemic model outperformed all other included models for most locations, demonstrating the continued competitiveness of ensemble modeling in short-term mpox forecasting.

Our study is not exempt from limitations. We used two sources for weekly time-series laboratory-confirmed mpox cases, each using varying approaches to compile and disseminate case information. Similarly, the reporting patterns for countries varied as well. Therefore, the trends noted above may be a function of reporting delays rather than true case fluctuation.

However, to account for the effect of reporting differences on the observed epidemic trajectories, we aggregate cases weekly to adjust for within-week reporting delay. Due to the unprecedented nature of the epidemic and the limited epidemic data available, we examined forecasts derived from the second-ranked spatial wave and *n*-sub-epidemic models. Frequently, there was little statistical support for the second-ranked models relative to the top-ranked model for both frameworks. The spatial-wave and ensemble *n*-sub-epidemic frameworks are semi-mechanistic and provide insight into the natural processes that formed the observed epidemic trends from the aggregated sub-epidemics. Nevertheless, both frameworks do not account for explicit mechanisms of reactive behavioral modification and interventions, which likely played a significant role in controlling the epidemic at local and global levels [6,8]. Finally, current models are not sensitive to seasonal or event-specific risk behaviors, which limits their application to short-term forecasts.

In conclusion, our findings further support the competitive performance of the aggregated sub-epidemic methodologies in producing short-term forecasts within the context of mpox, along with the utility of ensemble modeling therein producing short-term forecasts. However, the frameworks could be expanded to other disease types as they have shown utility in short-term forecasting for epidemiologically different diseases [16–18]. Finally, short-term forecasting tools are essential in the face of unprecedented pandemics and epidemics. Therefore, further evaluation and refinements of the spatial-wave and ensemble *n*-sub-epidemic frameworks could be achieved via comparison to other models since all the forecasting results are publicly available to the community for transparency and further investigation.

## Acknowledgments

We would like to acknowledge the GSU Advanced Research Computing Technology & Innovation Core (ARCTIC) team for their help with the development of the mpox forecast website.

## Contributions

A.B., G.C., R.L., and A.K., developed different portions of the code used in analysis. A.B. and G.C. retrieved and managed data; A.B., and G.C. analyzed the data. A.B., R.L., A.K., and G.C. wrote the first draft of the manuscript. All authors contributed to writing and revising subsequent versions of the manuscript. All authors read and approved the final manuscript.

## Data availability statement

All original data sources are stated within the paper. Individual forecasts, performance metrics, and Winkler scores will be made publicly available upon journal acceptance and can be made available during the peer-review process upon request. Source code for producing and evaluating the GAM, SLR, ARIMA, and Prophet forecasts will be made publicly available upon journal acceptance and can be made available during the peer-review process upon request. Source code for producing and evaluating the ensemble *n*-sub-epidemic forecasts is publicly available at: https://github.com/gchowell/ensemble_n-subepidemic_framework. Source code for producing and evaluating spatial-wave forecasts is publicly available at: https://github.com/gchowell/spatial_wave_subepidemic_framework.

## Supporting Information

**S1 Appendix. Visualizations of epidemic trajectories and epidemic phase-specific forecast performance**. This file contains multiple figures visualizing the mpox epidemic trajectory in each study location, and average forecast performance for each model across epidemic phases for each study location and performance metric.

**S2 Appendix. Sensitivity analysis comparing forecasting performance across different calibration periods**. This file contains materials related to the sensitivity analysis conducted to determine the optimal calibration period for the primary analysis.

**S3 Appendix. Tabulation of forecasting performance metrics for each model, location, forecasting horizon**. This file contains multiple tables that contain average forecast performance metrics for each location, model, forecasting horizon, epidemic phase and the overall average.

