## Supplementary material for "Retrospective evaluation of short-term forecast performance of ensemble sub-epidemic frameworks and other time-series models: The 2022-2023 mpox outbreak across multiple geographical scales, July 14^th^, 2022, through February 26th, 2023": S1 Appendix

**S1 Appendix: Visualizations of epidemic trajectories and epidemic phase-specific forecast performance.** This file contains multiple figures visualizing the mpox epidemic trajectory in each study location, and average forecast performance for each model across epidemic phases for each study location and performance metric.

Amanda Bleichrodt*[1], Ruiyan Luo[1], Alexander Kirpich[1], Gerardo Chowell

[1] Department of Population Health Sciences, School of Public Health, Georgia State University, Atlanta, GA, USA.

| Figure | Description | Pages |
|---|---|---|
| A | Epidemic trajectories for each study location. | 3 |
| B | Average MSE across epidemic phases. | 4 |
| C | Average MAE across epidemic phases. | 5 |
| D | Average 95% PI coverage across epidemic phases. | 6 |
| E | Average WIS across epidemic phases. | 7 |

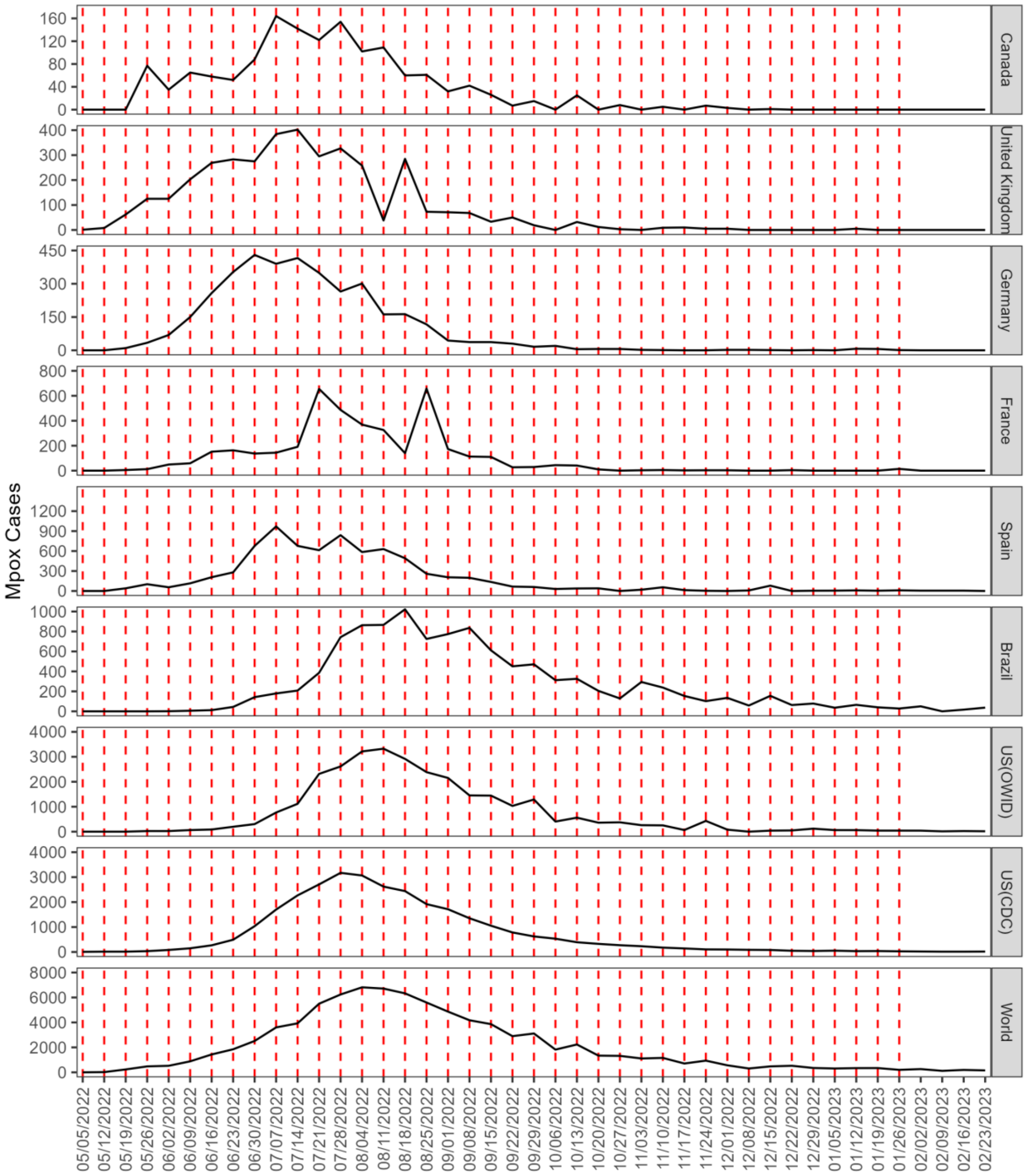


**Fig A.** The epidemic trajectories for each study location of interest. The black solid line is the reported cases as of the week of August 3rd, 2023. The red-dashed lines are each of the forecasting periods included within the analysis. Forecasts (1-4 weeks) were produced through February 23rd, 2023, with the last forecast period being the week of January 26th, 2023. Epidemic trajectories for Canada, the United Kingdom, Germany, France, Spain, Brazil, the US (OWID), and the World are from the Our World in Data (OWID) team [1] and from the Centers for Disease Control and Prevention (CDC) for US (CDC) [2].

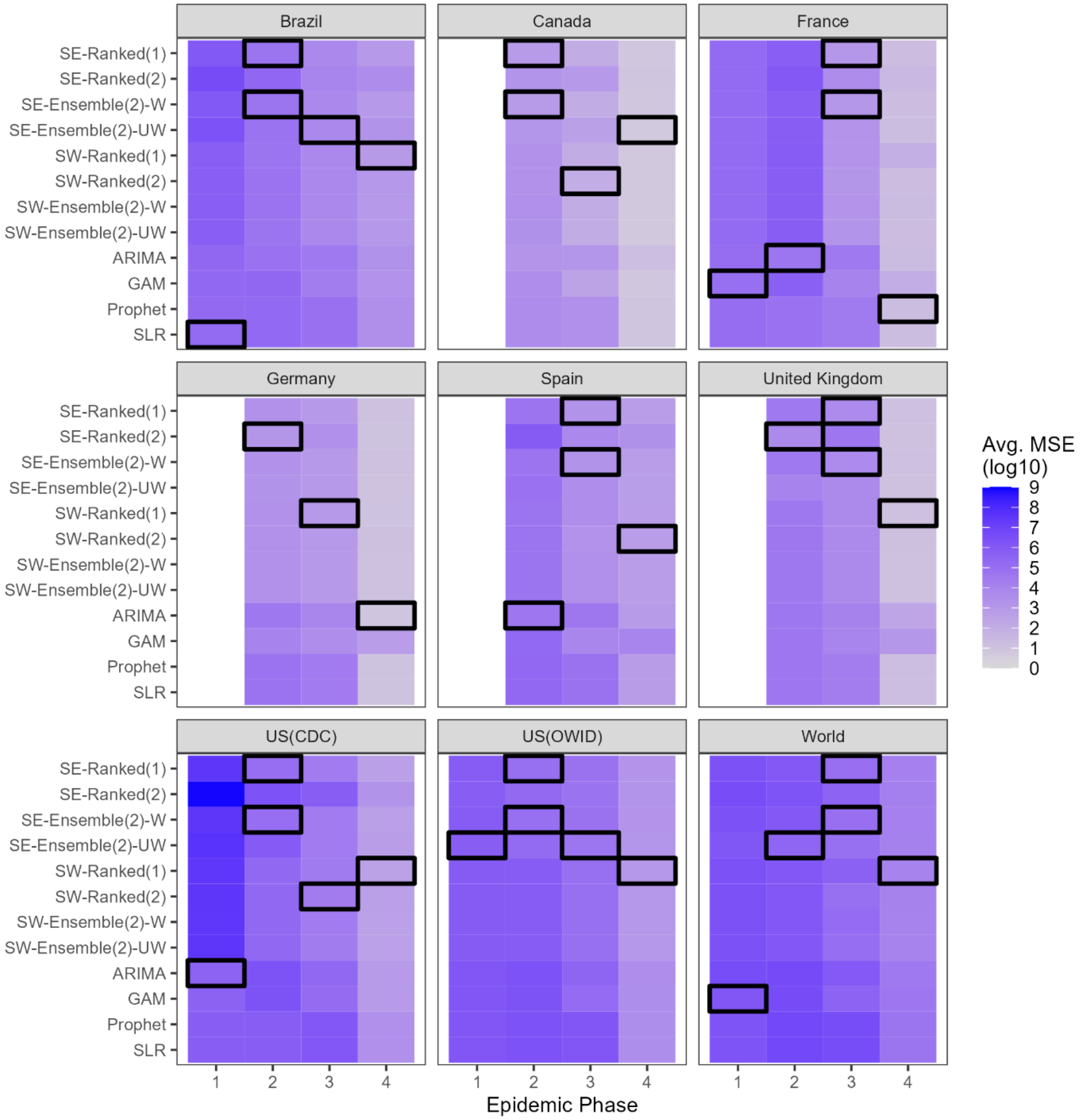


**Fig B.** The average mean squared error (MSE) averaged across forecasting horizons for each study location, model, and epidemic phase. The epidemic phase refers to the period in the disease trajectory (i.e., *1* = Ascending; *2* = Peak; *3* = Descending; *4* = Tail End of the epidemic). The black squares indicate the best-performing model during each epidemic phase. If more than one black box is present in a given epidemic phase, there was more than one best-performing model (i.e., the top-performing models produced the same average MSE value). The white columns seen in the ascending phase for Spain, Germany, Canada, and the United Kingdom indicate that forecast metrics were unavailable for the ascending phase of their respective epidemic. Regarding the included models, *SE* indicates the *n*-sub-epidemic framework, SW refers to the spatial-wave framework, ARIMA is the autoregressive-integrated moving-average model, GAM is a general additive model, and SLR is the simple linear regression model. The average MSE is shown in the *log10* scale.

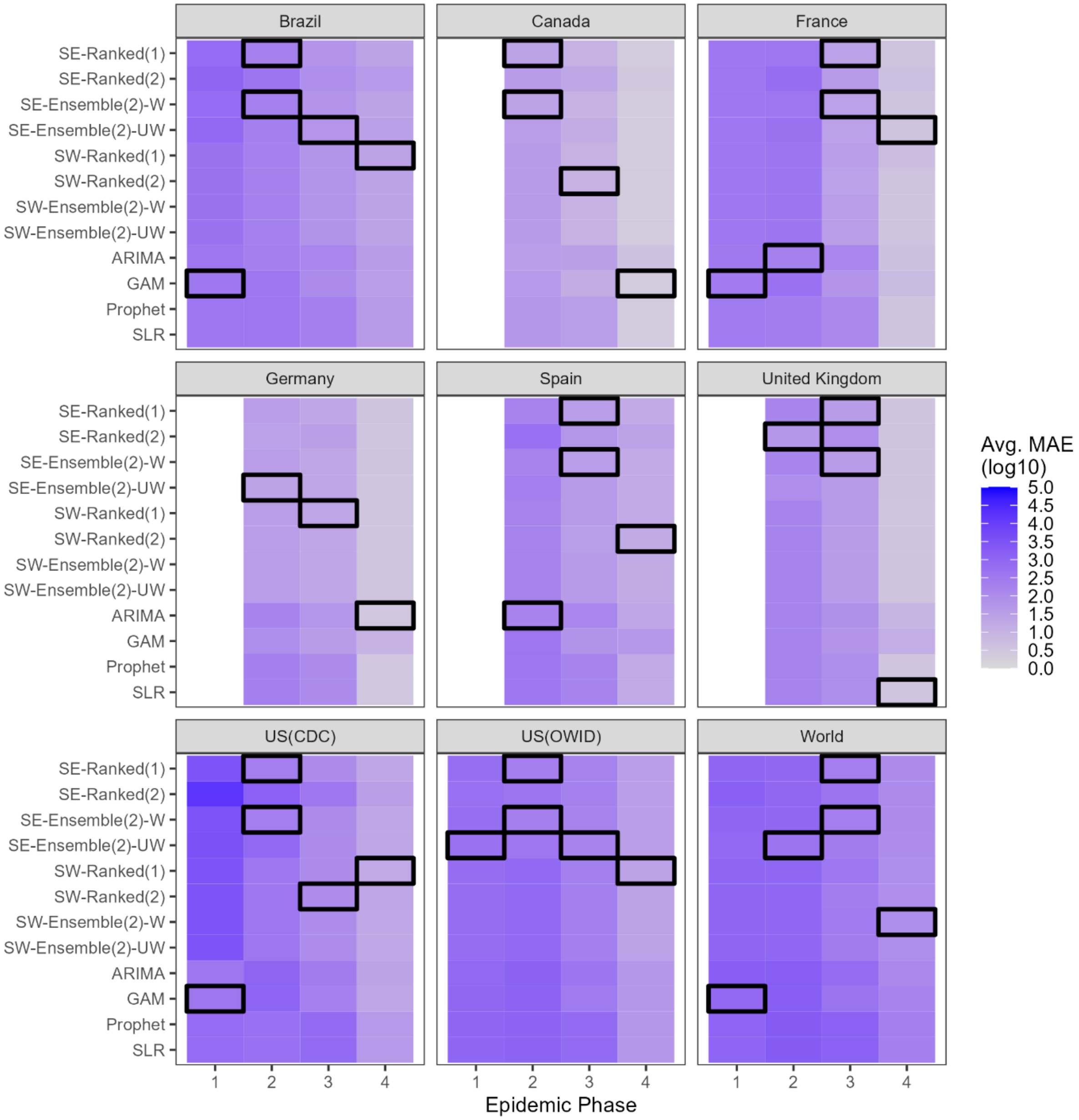


**Fig C.** The average mean absolute error (MAE) averaged across forecasting horizons for each study location, model, and epidemic phase. The epidemic phase refers to the period in the disease trajectory (i.e., *1* = Ascending; *2* = Peak; *3* = Descending; *4* = Tail End of the epidemic). The black squares indicate the best-performing model during each epidemic phase. If more than one black box is present in a given epidemic phase, there was more than one best-performing model (i.e., the top-performing models produced the same average MAE value). The white columns seen in the ascending phase for Spain, Germany, Canada, and the United Kingdom indicate that forecast metrics were unavailable for the ascending phase of their respective epidemic. Regarding the included models, *SE* indicates the *n*-sub-epidemic framework, SW refers to the spatial-wave framework, ARIMA is the autoregressive-integrated moving-average model, GAM is a general additive model, and SLR is the simple linear regression model. The average MAE is shown in the *log10* scale.

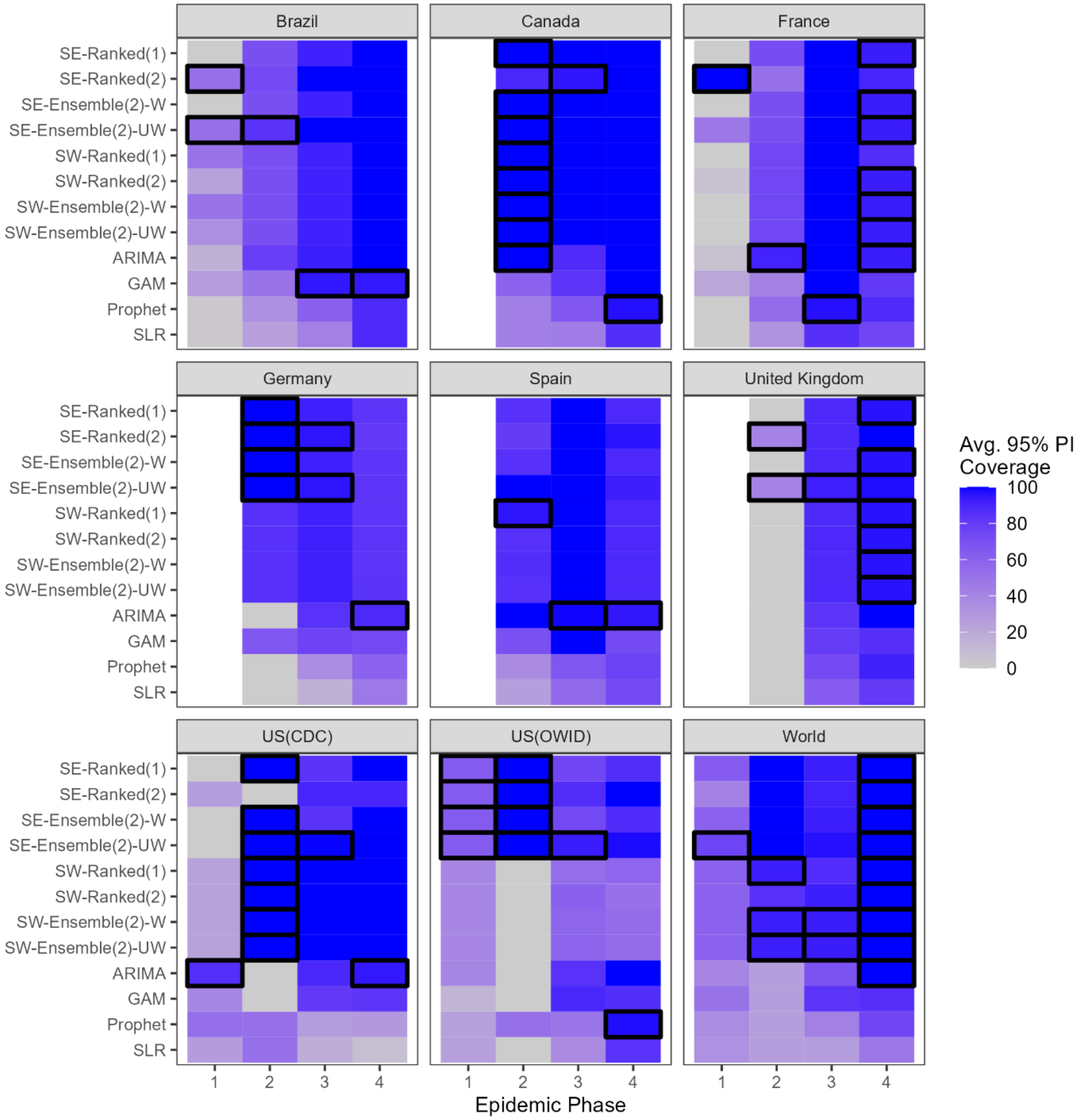


**Fig D.** The average 95% prediction interval coverage (95% PI) averaged across forecasting horizons for each study location, model, and epidemic phase. The epidemic phase refers to the period in the disease trajectory (i.e., *1* = Ascending; *2* = Peak; *3* = Descending; *4* = Tail End of the epidemic). The black squares indicate the best-performing model during each epidemic phase. Here, *best-performing* refers to the model with an average 95% PI coverage closest to 95%. If more than one black box is present in a given epidemic phase, there was more than one best-performing model (i.e., the top-performing models produced the same average 95% coverage value). The white columns seen in the ascending phase for Spain, Germany, Canada, and the United Kingdom indicate that forecast metrics were unavailable for the ascending phase of their respective epidemic. Regarding the included models, *SE* indicates the *n*-sub-epidemic framework, SW refers to the spatial-wave framework, ARIMA is the autoregressive-integrated moving-average model, GAM is a general additive model, and SLR is the simple linear regression model.

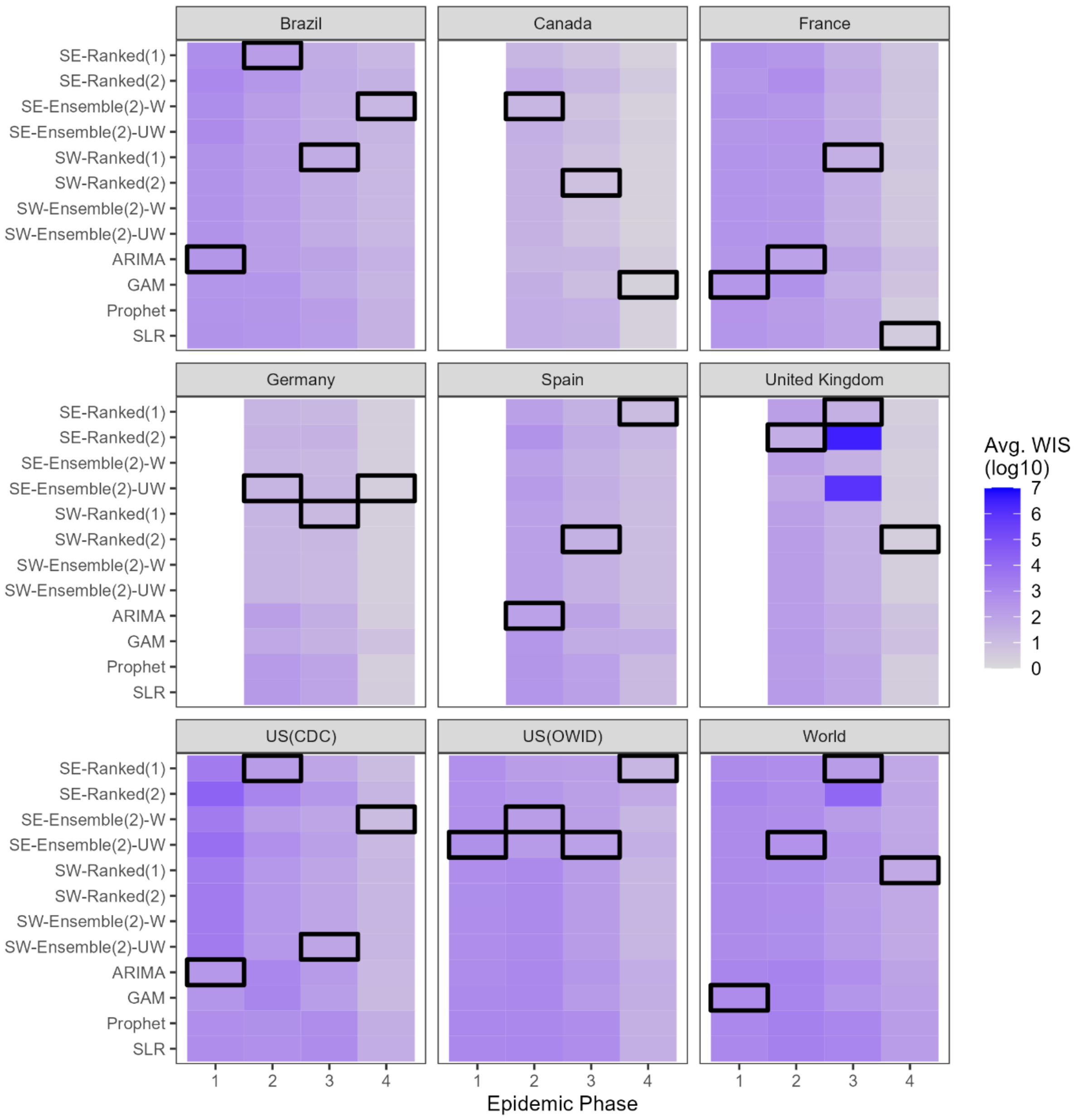


**Fig E.** The average weighted interval score (WIS) averaged across forecasting horizons for each study location, model, and epidemic phase. The epidemic phase refers to the period in the disease trajectory (i.e., *1* = Ascending; *2* = Peak; *3* = Descending; *4* = Tail End of the epidemic). The black squares indicate the best-performing model during each epidemic phase. If more than one black box is present in a given epidemic phase, there was more than one best-performing model (i.e., the top-performing models produced the same average WIS value). The white columns seen in the ascending phase for Spain, Germany, Canada, and the United Kingdom indicate that forecast metrics were unavailable for the ascending phase of their respective epidemic. Regarding the included models, *SE* indicates the *n*-sub-epidemic framework, SW refers to the spatial-wave framework, ARIMA is the autoregressive-integrated moving-average model, GAM is a general additive model, and SLR is the simple linear regression model. The average WIS is shown in the *log10* scale.
