## Supplementary material for "Retrospective evaluation of short-term forecast performance of ensemble sub-epidemic frameworks and other time-series models: The 2022-2023 mpox outbreak across multiple geographical scales, July 14^th^, 2022, through February 26th, 2023": S2 Appendix

**S2 Appendix. Sensitivity analysis comparing forecasting performance across different calibration periods:** A sensitivity analysis evaluating multi-model and ensemble forecasts in the context of the 2022-2023 mpox outbreak in multiple countries for 9-, 10- and 11-week calibration periods.

Amanda Bleichrodt*[1], Ruiyan Luo[1], Alexander Kirpich[1], Gerardo Chowell[1]

[1] Department of Population Health Sciences, School of Public Health, Georgia State University, Atlanta, GA, USA.

.

### 1. Purpose

This sensitivity analysis aims to determine the optimal calibration period length for producing short-term mpox forecasts using the methodology employed in the primary analysis.

### 2. Methodology

To determine the optimal calibration period length for producing short-term mpox forecasts using the methodology explained in the main manuscript, our team conducted 1-4 week out forecasts employing 9-, 10-, and 11-week calibration periods for the weeks of June 30th, 2022, through January 26th, 2023 (forecast periods). Therefore, we produced and evaluated forecasts from the week of July 7th, 2022, through the week of February 23rd, 2023. We produced the forecasts using the same data, models, and locations discussed in the primary analysis.

We evaluated each forecast employing both measures of accuracy (i.e., mean squared error and mean absolute error) and probabilistic measures of performance (i.e., weighted interval score and 95% prediction interval coverage). A full description of the four-performance metrics and evaluation methodology can be found in the main text.

After producing performance metrics for each forecast, we calculated each metric's average across locations, models, forecasting horizons, and calibration periods. Therefore, we have the average performance metrics for each unique combination of those factors. Next, we determined the number of times each calibration performance was the best performing (i.e., the metric "won") for a given location, forecasting horizon, model, and performance metric. A "win" is granted if the calibration period produces the lowest score for the mean squared error (MSE), mean absolute error (MAE), or weighted interval score (WIS) metrics. For 95% PI coverage, a win is granted to the model with coverage closest to 95%. For all performance metrics, there may be multiple models which perform equally well.

Finally, to determine the optimal calibration period for producing short-term mpox forecasts, we calculated the total number of "wins" for each calibration period, aggregated only by location.

### 3. Results and Conclusion

*Mean Squared Error (MSE)*

The 11-week calibration period performed best overall most frequently, followed by the 10-week and 9-week calibrations periods regarding average MSE. The 11-week calibration period produced the lowest average MSE for approximately 41.7- 66.7% of the time across models and forecasting horizons in Brazil, Germany, Spain, the United Kingdom, US (OWID), and the World. However, the 10-week calibration performed best in Canada (62.5% of the time), and the US (CDC) (43.8% of the time), and the 9-week calibration period saw success only in France (54.2% of the time). Figure 1s below includes the overall number of times each calibration period performs best regarding average MSE for each study location.

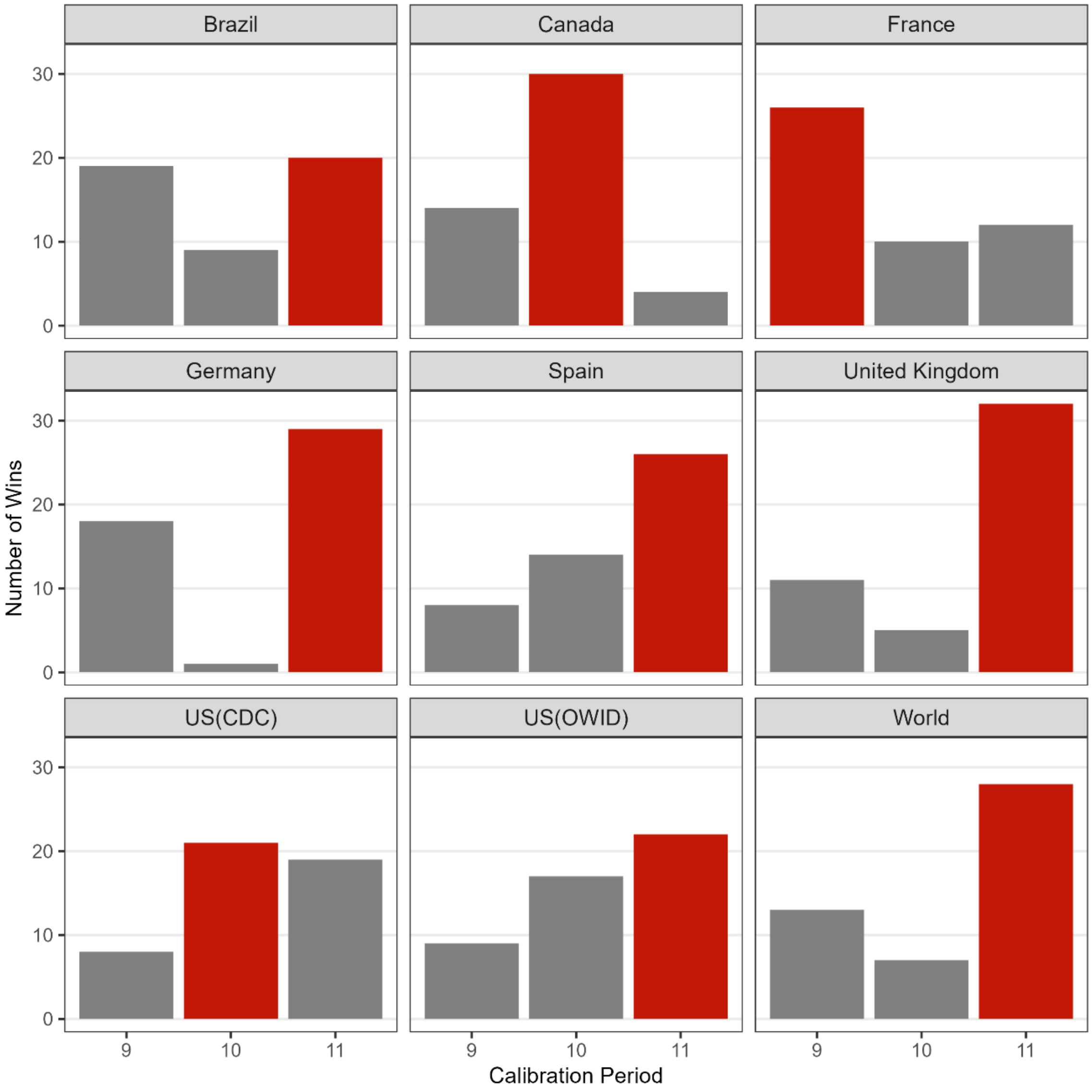


**Figure 1s. Overall performance of each calibration period by location regarding average MSE.** The figure above illustrates the number of forecasts where each calibration period produced the lowest average MSE compared to the other two calibration periods. The red bar indicates the best performing calibration period overall for each study location. The 11-week calibration period produced the lowest average MSE most frequently across study locations.

*Mean Absolute Error (MAE)*

The 11-week calibration period also performed best overall regarding average MAE, followed by the 10-week and 9-week calibration periods. The 11-week calibration period produced the lowest average MAE for approximately 45.8- 66.7% of the time in Germany, Spain, the United Kingdom, the US(CDC), and the World. The 10-week calibration performed best in Canada (66.7% of the time), France (39.6% of the time), and the US (OWID) (43.8% of the time), and the 9-week calibration period saw success only in Brazil (41.7% of the time). Figure 2s below includes the overall number of times each calibration period performs best regarding average MAE for each study location.

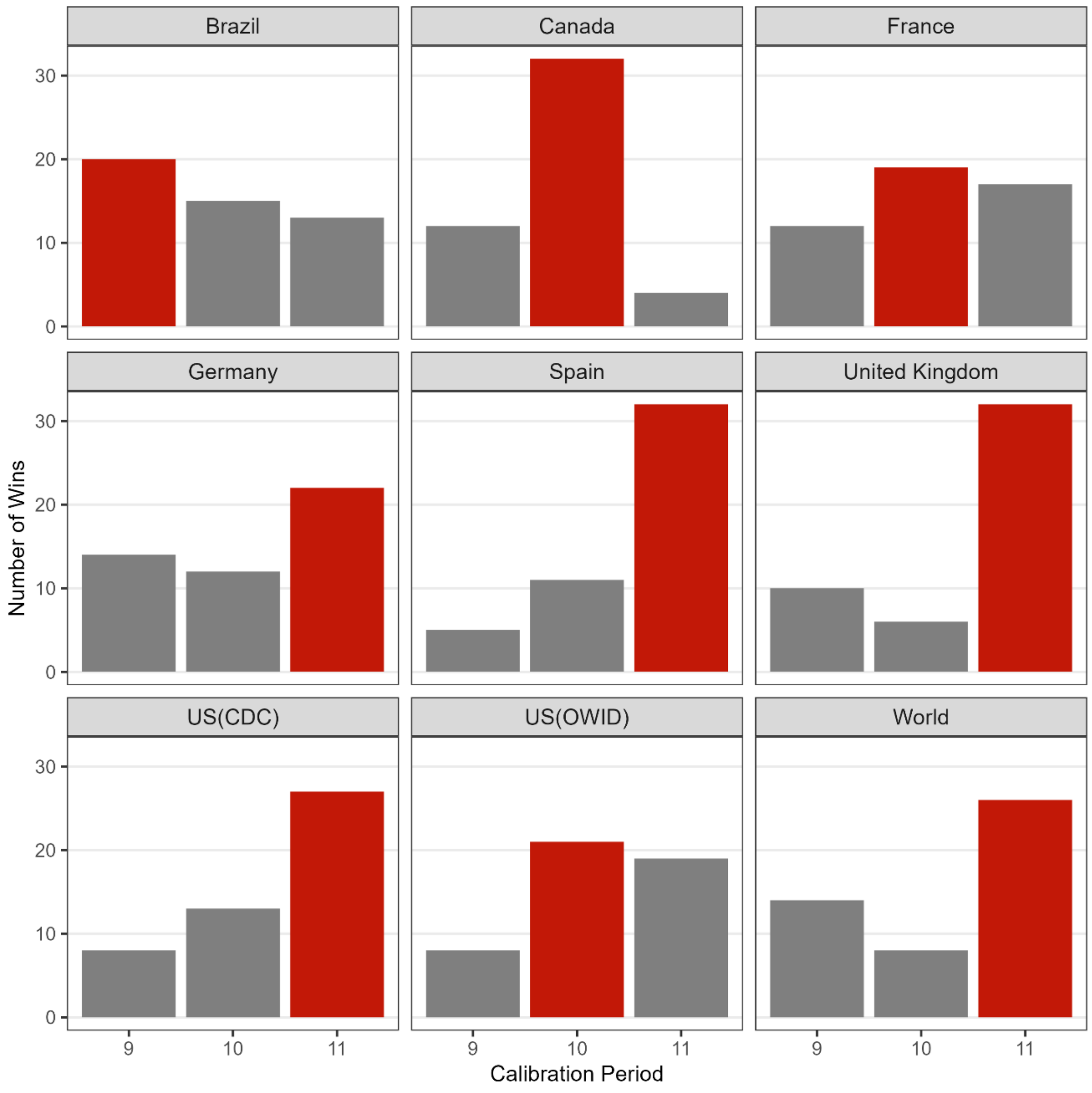


**Figure 2s. Overall performance of each calibration period by location regarding average MAE.** The figure above illustrates the number of forecasts where each calibration period produced the lowest average MAE compared to the other two calibration periods. The red bar indicates the best performing calibration period overall for each study location. The 11-week calibration period produced the lowest average MAE most frequently across study locations.

*Weighted Interval Score (WIS)*

As with the average MAE and MSE metrics, the 11-week calibration produced the lowest average WIS most frequently across study locations, followed in success by the 10-week calibration period. The 9-week calibration was not the best performing calibration period for any study location included. The 11-week calibration period saw the most success in Brazil, Germany, Spain, the United Kingdom, the US(CDC), and the World, performing best approximately 43.8-64.6% of the time. The 10-week calibration period performed best overall for Canada (83.3% of the time), France (52.1% of the time), and the US (OWID) (43.8% of the time). Figure 3s below includes the overall number of times each calibration period performs best regarding average WIS for each study location.

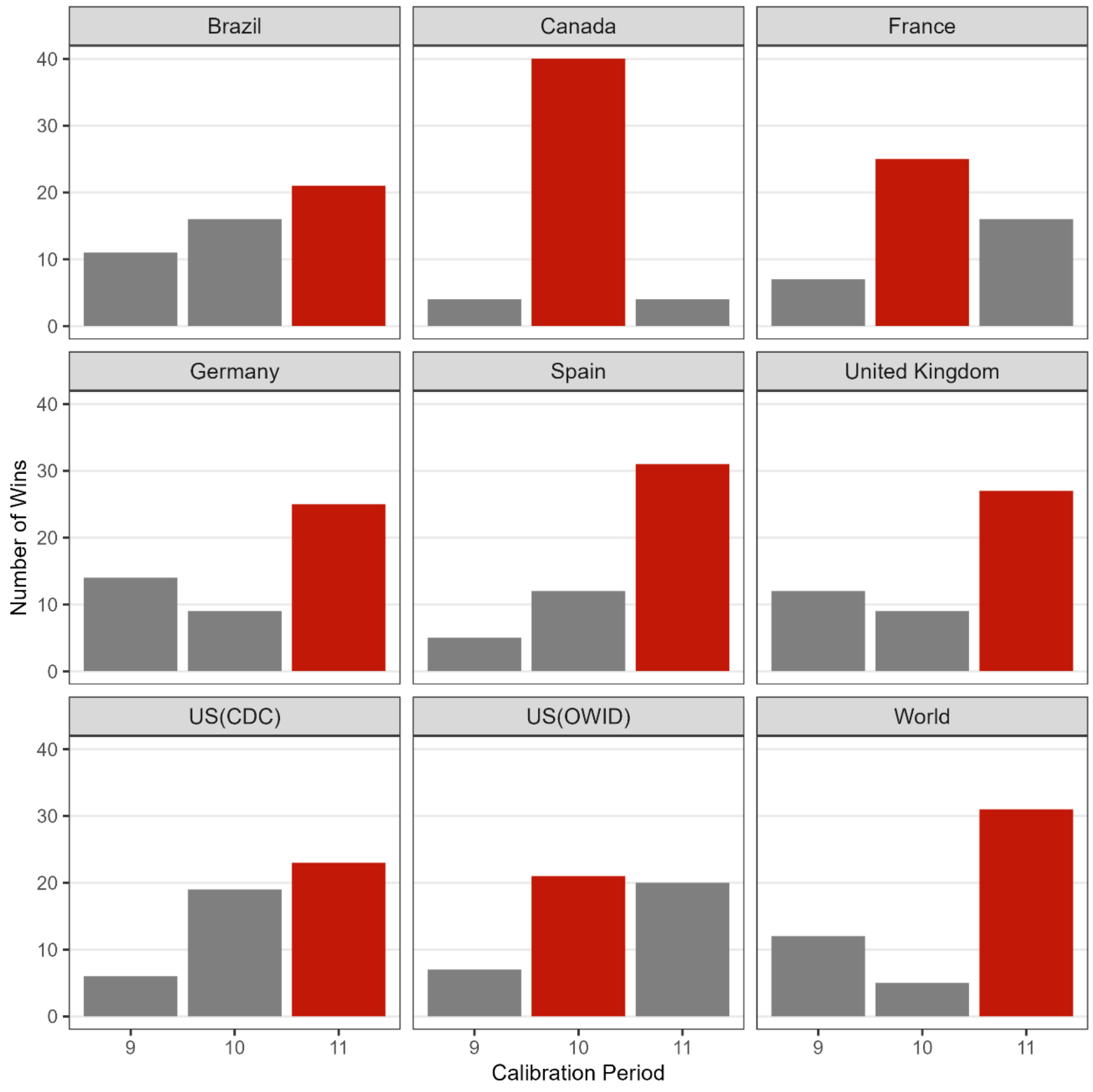


**Figure 3s. Overall performance of each calibration period by location regarding average WIS.** The figure above illustrates the number of forecasts where each calibration period produced the lowest average WIS compared to the other two calibration periods. The red bar indicates the best performing calibration period overall for each study location. The 11-week calibration period produced the lowest average WIS most frequently across study locations.

*95% Prediction Interval Coverage*

Finally, the 11-week calibration produced the closest to 95% PI coverage most frequently across study locations, followed in success by the 9-week and 10-week calibration periods. The 11-week calibration period performed best in France (50.0% of the time), Spain (60.4% of the time), the United Kingdom (56.3% of the time), and the World (54.2% of the time) across forecasting periods and horizons. Brazil (58.3%), Canada (75.0%), and the US(OWID) (41.7%) saw the most success with the 9-week calibration period, and the 10-week calibration period performed best in Germany (50.0%) and US(CDC) (47.9%) compared to the other calibration periods when looking across forecasting periods and horizons. Figure 4s below includes the overall number of times each calibration period performs best regarding average 95% PI coverage for each study location.

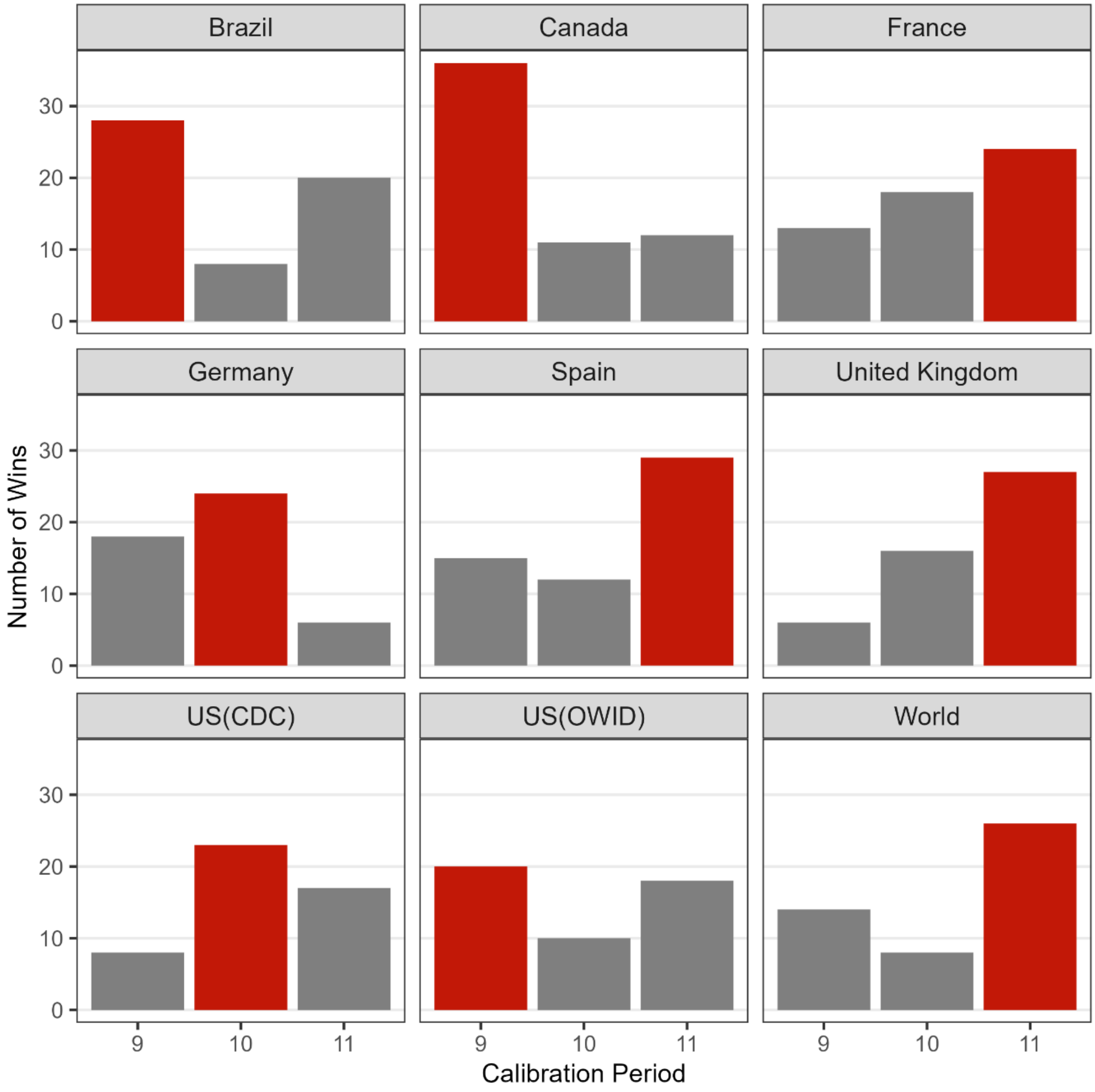


**Figure 4s. Overall performance of each calibration period by location regarding average 95% PI coverage.** The figure above illustrates the number of forecasts where each calibration period produced the closest to average 95% PI compared to the other two calibration periods. The red bar indicates the best performing calibration period overall for each study location. The 11-week calibration period produced the best average 95% PI coverage most frequently across study locations.

*Conclusion*

Overall, we found that the 11-week calibration period is best suited to our desired analysis. The 11-week calibration period performed best most frequently across models, forecasting horizons, and performance metrics compared to the 9- and 10-week calibration periods (Table 1s).

**Table 1s.** Frequency[1] and percentage [2] of “wins” for each calibration period over all locations.

| | **9-Week[3]** | **10-Week[3]** | **11-Week[3]** |
|---|---|---|---|
| Mean Squared Error (MSE) | 1 (11.1%) | 2 (22.2%) | 6 (66.7%) |
| Mean Absolute Error (MAE) | 1 (11.1%) | 3 (33.3%) | 5 (55.6%) |
| 95% PI Coverage (95% PI) | 3 (33.3%) | 2 (22.2%) | 4 (44.4%) |
| Weighted Interval Score (WIS) | 0 (0.0%) | 3 (33.3%) | 6 (66.7%) |

[1] The number of times a given calibration period produced the best average forecasting performance metric over all models, forecasting periods (i.e., 1-4 weeks), and locations.

[2] The percentage of times, compared to the other calibration period lengths, a given calibration period produces the best average forecasting performance metric over all models, forecasting periods (i.e., 1-4 weeks), and locations.

[3] Calibration period length.
