## Supplementary material for "Retrospective evaluation of short-term forecast performance of ensemble sub-epidemic frameworks and other time-series models: The 2022-2023 mpox outbreak across multiple geographical scales, July 14^th^, 2022, through February 26th, 2023": S3 Appendix

**S3 Appendix: Tabulation of forecasting performance metrics for each model, location, forecasting horizon.**

The average forecasting performance metrics for each model, location, epidemic phase, and overall, and tables with included the skill scores for each metric and location.

Amanda Bleichrodt*[1], Ruiyan Luo[1], Alexander Kirpich[1], Gerardo Chowell

[1] Department of Population Health Sciences, School of Public Health, Georgia State University, Atlanta, GA, USA.

| Country | | Description | Pages |
|---|---|---|---|
| A. Brazil | | | 3-7 |
| | **Table A1:** | Average Mean Squared Error | 3 |
| | **Table A2:** | Average Mean Absolute Error | 4 |
| | **Table A3:** | Average 95% Prediction Interval Coverage | 5 |
| | **Table A4:** | Average Weighted Interval Score | 6 |
| | **Table A5:** | Skill Scores | 7 |
| B. Canada | | | 8-12 |
| | **Table B1:** | Average Mean Squared Error | 8 |
| | **Table B2:** | Average Mean Absolute Error | 9 |
| | **Table B3:** | Average 95% Prediction Interval Coverage | 10 |
| | **Table B4:** | Average Weighted Interval Score | 11 |
| | **Table B5:** | Skill Scores | 12 |
| C. France | | | 13-17 |
| | **Table C1:** | Average Mean Squared Error | 13 |
| | **Table C2:** | Average Mean Absolute Error | 14 |
| | **Table C3:** | Average 95% Prediction Interval Coverage | 15 |
| | **Table C4:** | Average Weighted Interval Score | 16 |
| | **Table C5:** | Skill Scores | 17 |
| D. Germany | | | 18-22 |
| | **Table D1:** | Average Mean Squared Error | 18 |
| | **Table D2:** | Average Mean Absolute Error | 19 |
| | **Table D3:** | Average 95% Prediction Interval Coverage | 20 |
| | **Table D4:** | Average Weighted Interval Score | 21 |
| | **Table D5:** | Skill Scores | 22 |
| E. Spain | | | 23-27 |
| | **Table E1:** | Average Mean Squared Error | 23 |
| | **Table E2:** | Average Mean Absolute Error | 24 |
| | **Table E3:** | Average 95% Prediction Interval Coverage | 25 |
| | **Table E4:** | Average Weighted Interval Score | 26 |
| | **Table E5:** | Skill Scores | 27 |
| F. United Kingdom | | | 28-32 |
| | **Table F1:** | Average Mean Squared Error | 28 |
| | **Table F2:** | Average Mean Absolute Error | 29 |
| | **Table F3:** | Average 95% Prediction Interval Coverage | 30 |
| | **Table F4:** | Average Weighted Interval Score | 31 |
| | **Table F5:** | Skill Scores | 32 |
| G. United States (CDC) | | | 33-37 |
| | **Table G1:** | Average Mean Squared Error | 33 |
| | **Table G2:** | Average Mean Absolute Error | 34 |
| | **Table G3:** | Average 95% Prediction Interval Coverage | 35 |
| | **Table G4:** | Average Weighted Interval Score | 36 |
| | **Table G5:** | Skill Scores | 37 |
| H. United States (OWID) | | | 38-42 |
| | **Table H1:** | Average Mean Squared Error | 38 |
| | **Table H2:** | Average Mean Absolute Error | 39 |
| | **Table H3:** | Average 95% Prediction Interval Coverage | 40 |
| | **Table H4:** | Average Weighted Interval Score | 41 |
| | **Table H5:** | Skill Scores | 42 |
| I. World | | | 43-47 |
| | **Table I1:** | Average Mean Squared Error | 43 |
| | **Table I2:** | Average Mean Absolute Error | 44 |
| | **Table I3:** | Average 95% Prediction Interval Coverage | 45 |
| | **Table I4:** | Average Weighted Interval Score | 46 |
| | **Table I5:** | Skill Scores | 47 |

**Table A1.** Average Mean Squared Error (MSE) of the forecasts generated for Brazil [1] (weeks of July 14th, 2022, through February 23rd, 2023) for each forecasting horizon (1-4 weeks), time-period [2] of the epidemic, and averaged across all forecast periods (*Avg.*) for each model [3] of interest.

| Time Period | SW 1st Ranked | SW 2nd Ranked | SW Weighted Ensemble (2) | SW Unweighted Ensemble (2) | SE 1st Ranked | SE 2nd Ranked | SE Weighted Ensemble (2) | SE Unweighted Ensemble (2) | Simple Linear Regression | Prophet | General Additive Model | ARIMA[3] |
|---|---|---|---|---|---|---|---|---|---|---|---|---|
| **1-Week Forecasting Horizon** | | | | | | | | | | | | |
| 1 | 75158.57 | 68956.83 | 78078.84 | 68389.98 | 68759.56 | **10071.12** | 68759.56 | 20611.94 | 127075.87 | 127221.16 | 36379.83 | 51439.84 |
| 2 | 48622.95 | 50442.14 | 49201.67 | 49515.21 | 49853.81 | 122967.73 | 49853.81 | 47675.49 | 95613.97 | 95623.28 | 84323.46 | **39045.20** |
| 3 | 6632.39 | 7315.69 | 7080.54 | 7181.08 | 6160.31 | 8653.31 | 6160.31 | **5264.56** | 59085.71 | 58957.96 | 11195.84 | 22823.21 |
| 4 | 1086.98 | 1294.06 | 1239.44 | 1275.08 | **1066.88** | 4880.71 | **1066.88** | 2516.82 | 2826.46 | 2810.93 | 2242.39 | 2864.90 |
| Avg. | 18544.59 | 18809.43 | 19091.70 | 18515.46 | 16302.01 | 32711.77 | 16302.01 | **14239.95** | 54277.01 | 54244.55 | 25843.54 | 22257.65 |
| **2-Week Forecasting Horizon** | | | | | | | | | | | | |
| 1 | 258461.89 | 244622.18 | 249655.59 | 248993.88 | 399571.11 | 566242.46 | 399571.11 | 490639.95 | 177661.87 | 177831.99 | **138335.26** | 159756.76 |
| 2 | 54466.87 | 55683.79 | 56426.69 | 55003.94 | **52356.30** | 298529.36 | **52356.30** | 67188.49 | 155917.08 | 156007.33 | 195954.25 | 60843.72 |
| 3 | 6452.81 | 6566.70 | 6543.97 | 6697.45 | 6505.60 | 9588.82 | 6505.60 | **5590.86** | 70479.26 | 70380.72 | 16827.10 | 32934.50 |
| 4 | **987.41** | 1226.93 | 1093.35 | 1152.78 | 1018.10 | 4161.66 | 1018.10 | 1984.06 | 2881.08 | 2864.46 | 2039.88 | 2011.23 |
| Avg. | 32790.26 | 32188.69 | 32651.59 | 32375.49 | **29220.22** | 92404.57 | **29220.22** | 35908.15 | 75934.19 | 75930.75 | 61358.66 | 41165.17 |
| **3-Week Forecasting Horizon** | | | | | | | | | | | | |
| 1 | 613211.64 | 588330.25 | 603672.05 | 579876.78 | 1033326.66 | 2921491.53 | 1033326.66 | 1975078.47 | **211835.76** | 212019.83 | 244241.75 | 260565.06 |
| 2 | 64464.16 | 65010.64 | 65073.65 | 63772.89 | **61581.28** | 317730.56 | **61581.28** | 71754.16 | 223633.64 | 223750.05 | 270277.53 | 88506.42 |
| 3 | **6097.90** | 6175.38 | 6222.46 | 6270.46 | 6329.82 | 10742.69 | 6329.82 | 6152.49 | 85576.02 | 85481.68 | 26648.49 | 39819.35 |
| 4 | **1019.70** | 1204.81 | 1113.79 | 1147.54 | 1092.88 | 4086.09 | 1092.88 | 1996.62 | 3287.46 | 3278.21 | 2233.78 | 2215.62 |
| Avg. | 60156.58 | 58590.22 | 59683.88 | 57734.68 | **54705.23** | 184302.32 | **54705.23** | 92114.61 | 98825.65 | 98833.06 | 91145.28 | 59761.50 |
| **4-Week Forecasting Horizon** | | | | | | | | | | | | |
| 1 | 1679908.50 | 1622969.86 | 1662213.19 | 1659397.16 | 2963085.72 | 12973285.64 | 2963085.72 | 6726129.30 | **225495.91** | 225699.21 | 455421.11 | 479517.46 |
| 2 | 81957.00 | 82275.69 | 82592.04 | 81068.78 | **73956.00** | 287762.68 | **73956.00** | 87344.66 | 318631.41 | 318712.56 | 400193.19 | 131994.01 |
| 3 | **6115.38** | 6160.96 | 6196.22 | 6198.81 | 6463.60 | 11981.04 | 6463.60 | 6907.78 | 98689.88 | 98595.72 | 33748.71 | 47177.09 |
| 4 | **860.73** | 1006.36 | 944.67 | 943.14 | 926.11 | 3536.36 | 926.11 | 1643.33 | 2988.63 | 2980.55 | 2140.46 | 2195.86 |
| Avg. | 140047.15 | 136116.68 | 138978.13 | 138450.95 | 128915.51 | 550186.51 | 128915.51 | 271692.90 | 124312.46 | 124315.03 | 142286.70 | **93939.52** |

[1] Data was obtained from the Our World in Data Team (OWID) [1].
[2] The time-period of the epidemic, where *1* refers to ascending phase of the epidemic (July 2022), *2* the peak (August-Early Sept. 2022), *3* for the immediate declining phase (Mid-September 2022, Mid-November 2022), and *4* for the tail-end of the epidemic (Late November 2022 - Current).
[3] The models of interest include models of the spatial-wave framework (SW), and *n*-sub-epidemic frameworks (SE), a simple linear regression model, Facebook's Prophet model (Prophet), a general additive model, and an auto-regressive integrated moving average model (ARIMA). All models were calibrated with 11-weeks' worth of data at a time.
* The bolded values indicate the best performing model(s) at each time-period for each forecasting horizon. Here, *best performing* refers to the model with the lowest metric value.

**Table A2.** Average Mean Absolute Error (MAE) of the forecasts generated for Brazil [1] (weeks of July 14th, 2022, through February 23rd, 2023) for each forecasting horizon (1-4 weeks), and time-period [2] of the epidemic, and averaged across all forecast periods (*Avg.*) for each model [3] of interest.

| Time Period | SW 1st Ranked | SW 2nd Ranked | SW Weighted Ensemble (2) | SW Unweighted Ensemble (2) | SE 1st Ranked | SE 2nd Ranked | SE Weighted Ensemble (2) | SE Unweighted Ensemble (2) | Simple Linear Regression | Prophet | General Additive Model | ARIMA[3] |
|---|---|---|---|---|---|---|---|---|---|---|---|---|
| **1-Week Forecasting Horizon** | | | | | | | | | | | | |
| 1 | 272.31 | 262.25 | 277.44 | 260.97 | 262.22 | **100.35** | 262.22 | 143.57 | 341.44 | 341.65 | 169.74 | 214.78 |
| 2 | 190.43 | 193.28 | 192.18 | 191.23 | 198.71 | 336.59 | 198.71 | 210.75 | 283.42 | 283.64 | 246.24 | **153.52** |
| 3 | 65.69 | 68.97 | 68.29 | 68.44 | 60.89 | 86.36 | 60.89 | **55.46** | 189.31 | 189.12 | 86.77 | 117.10 |
| 4 | 26.97 | 28.86 | 27.19 | 28.01 | **26.90** | 50.06 | **26.90** | 33.69 | 38.74 | 38.61 | 33.84 | 43.46 |
| Avg. | 93.35 | 95.09 | 95.10 | 94.07 | 86.38 | 129.04 | 86.38 | **85.17** | 172.60 | 172.55 | 110.09 | 109.35 |
| **2-Week Forecasting Horizon** | | | | | | | | | | | | |
| 1 | 415.85 | 408.75 | 412.68 | 410.41 | 558.43 | 580.10 | 558.43 | 560.25 | 402.87 | 403.07 | **297.40** | 332.01 |
| 2 | 191.76 | 194.32 | 195.44 | 192.65 | **183.94** | 443.50 | **183.94** | 221.93 | 357.09 | 357.23 | 369.09 | 202.19 |
| 3 | 63.89 | 64.15 | 63.98 | 64.81 | 62.83 | 82.63 | 62.83 | **54.47** | 213.65 | 213.52 | 100.52 | 151.69 |
| 4 | **24.04** | 26.49 | 24.31 | 25.26 | 24.64 | 45.84 | 24.64 | 30.00 | 40.48 | 40.33 | 34.01 | 35.08 |
| Avg. | 102.20 | 103.21 | 102.89 | 102.76 | **93.95** | 167.62 | **93.95** | 101.36 | 203.19 | 203.14 | 153.52 | 140.58 |
| **3-Week Forecasting Horizon** | | | | | | | | | | | | |
| 1 | 550.47 | 547.07 | 548.83 | 539.84 | 877.92 | 1307.80 | 877.92 | 1116.22 | 441.96 | 442.16 | 410.35 | **404.90** |
| 2 | 211.52 | 213.96 | 213.58 | 210.97 | **207.00** | 465.22 | **207.00** | 231.11 | 419.73 | 419.78 | 428.85 | 253.81 |
| 3 | 63.66 | 63.64 | 64.16 | 64.31 | 63.72 | 84.68 | 63.72 | **58.26** | 230.96 | 230.87 | 125.85 | 163.11 |
| 4 | **24.32** | 26.76 | 25.00 | 25.66 | 24.96 | 46.26 | 24.96 | 30.38 | 42.70 | 42.60 | 35.23 | 36.25 |
| Avg. | 116.07 | 117.21 | 116.81 | 115.90 | **111.36** | 200.32 | **111.36** | 125.53 | 226.92 | 226.89 | 186.72 | 163.14 |
| **4-Week Forecasting Horizon** | | | | | | | | | | | | |
| 1 | 810.22 | 795.28 | 803.80 | 805.62 | 1398.08 | 2622.80 | 1398.08 | 1983.65 | **440.73** | 440.92 | 534.86 | 502.13 |
| 2 | 236.29 | 238.13 | 238.22 | 235.73 | **225.41** | 449.97 | **225.41** | 249.63 | 492.00 | 492.00 | 506.33 | 309.67 |
| 3 | 64.90 | **64.49** | 64.99 | 64.93 | 66.13 | 87.86 | 66.13 | 64.62 | 241.41 | 241.33 | 140.09 | 173.35 |
| 4 | **22.46** | 24.26 | 23.03 | 23.02 | 23.41 | 44.28 | 23.41 | 28.21 | 42.01 | 41.93 | 36.43 | 36.90 |
| Avg. | 139.71 | 139.53 | 139.90 | 139.47 | **135.03** | 246.08 | **135.03** | 163.32 | 245.12 | 245.08 | 220.96 | 188.51 |

[1] Data was obtained from the Our World in Data Team (OWID) [1].
[2] The time-period of the epidemic, where *1* refers to ascending phase of the epidemic (July 2022), *2* the peak (August-Early Sept. 2022), *3* for the immediate declining phase (Mid-September 2022, Mid-November 2022), and *4* for the tail-end of the epidemic (Late November 2022 - Current).
[3] The models of interest include models of the spatial-wave framework (SW), and *n*-sub-epidemic frameworks (SE), a simple linear regression model, Facebook's Prophet model (Prophet), a general additive model, and an auto-regressive integrated moving average model (ARIMA). All models were calibrated with 11-weeks' worth of data at a time.
* The bolded values indicate the best performing model(s) at each time-period for each forecasting horizon. Here, *best performing* refers to the model with the lowest metric value.

**Table A3.** Average 95% Prediction Interval Coverage (95% PI Coverage) of the forecasts generated for Brazil [1] (weeks of July 14th, 2022, through February 23rd, 2023) for each forecasting horizon (1-4 weeks), and time-period [2] of the epidemic, and averaged across all forecast periods (*Avg.*) for each model [3] of interest.

| Time Period | SW 1st Ranked | SW 2nd Ranked | SW Weighted Ensemble (2) | SW Unweighted Ensemble (2) | SE 1st Ranked | SE 2nd Ranked | SE Weighted Ensemble (2) | SE Unweighted Ensemble (2) | Simple Linear Regression | Prophet | General Additive Model | ARIMA[3] |
|---|---|---|---|---|---|---|---|---|---|---|---|---|
| **1-Week Forecasting Horizon** | | | | | | | | | | | | |
| 1 | 50.00 | 0.00 | 50.00 | 0.00 | 0.00 | **100.00** | 0.00 | **100.00** | 0.00 | 0.00 | 33.33 | 0.00 |
| 2 | 66.67 | 66.67 | 66.67 | 66.67 | 66.67 | **83.33** | 66.67 | **83.33** | 33.33 | 50.00 | 50.00 | **83.33** |
| 3 | **90.00** | **90.00** | **90.00** | **90.00** | **90.00** | **100.00** | **90.00** | **100.00** | 50.00 | 70.00 | **100.00** | **90.00** |
| 4 | **100.00** | **100.00** | **100.00** | **100.00** | **100.00** | **100.00** | **100.00** | **100.00** | **90.00** | **90.00** | **90.00** | **100.00** |
| Avg. | 85.71 | 82.14 | 85.71 | 82.14 | 85.19 | **96.30** | 85.19 | **96.30** | 55.17 | 65.52 | 79.31 | 82.76 |
| **2-Week Forecasting Horizon** | | | | | | | | | | | | |
| 1 | **50.00** | 25.00 | **50.00** | **50.00** | 0.00 | **50.00** | 0.00 | **50.00** | 0.00 | 0.00 | 33.33 | 16.67 |
| 2 | 75.00 | 75.00 | 75.00 | 75.00 | 75.00 | 66.67 | 75.00 | **83.33** | 25.00 | 33.33 | 50.00 | 75.00 |
| 3 | 90.00 | 90.00 | 90.00 | 90.00 | 90.00 | 100.00 | 90.00 | 100.00 | 45.00 | 65.00 | **95.00** | **95.00** |
| 4 | 100.00 | 100.00 | 100.00 | 100.00 | 100.00 | 100.00 | 100.00 | 100.00 | 90.00 | 90.00 | **95.00** | 100.00 |
| Avg. | 87.50 | 85.71 | 87.50 | 87.50 | 87.04 | 90.74 | 87.04 | **94.44** | 51.72 | 60.34 | 79.31 | 84.48 |
| **3-Week Forecasting Horizon** | | | | | | | | | | | | |
| 1 | **50.00** | 33.33 | **50.00** | **50.00** | 0.00 | 33.33 | 0.00 | 33.33 | 0.00 | 0.00 | 22.22 | 22.22 |
| 2 | 72.22 | 72.22 | 72.22 | 72.22 | 72.22 | 72.22 | 72.22 | **83.33** | 22.22 | 27.78 | 50.00 | 77.78 |
| 3 | **93.33** | **93.33** | **93.33** | **93.33** | **93.33** | 100.00 | **93.33** | 100.00 | 40.00 | 60.00 | **93.33** | **93.33** |
| 4 | 100.00 | 100.00 | 100.00 | 100.00 | 100.00 | 100.00 | 100.00 | 100.00 | 86.67 | 86.67 | **96.67** | 100.00 |
| Avg. | 88.10 | 86.90 | 88.10 | 88.10 | 87.65 | 91.36 | 87.65 | **93.83** | 48.28 | 56.32 | 78.16 | 85.06 |
| **4-Week Forecasting Horizon** | | | | | | | | | | | | |
| 1 | **50.00** | 37.50 | **50.00** | 37.50 | 0.00 | 25.00 | 0.00 | 25.00 | 8.33 | 8.33 | 16.67 | 25.00 |
| 2 | 66.67 | 66.67 | 66.67 | 66.67 | 66.67 | 70.83 | 66.67 | **87.50** | 20.83 | 25.00 | 50.00 | 79.17 |
| 3 | **95.00** | **95.00** | **95.00** | **95.00** | **95.00** | 100.00 | **95.00** | 100.00 | 35.00 | 47.50 | 92.50 | 92.50 |
| 4 | 100.00 | 100.00 | 100.00 | 100.00 | 100.00 | 100.00 | 100.00 | 100.00 | 87.50 | 87.50 | **97.50** | 100.00 |
| Avg. | 87.50 | 86.61 | 87.50 | 86.61 | 87.04 | 90.74 | 87.04 | **94.44** | 47.41 | 52.59 | 77.59 | 85.34 |

[1] Data was obtained from the Our World in Data Team (OWID) [1].
[2] The time-period of the epidemic, where *1* refers to ascending phase of the epidemic (July 2022), *2* the peak (August-Early Sept. 2022), *3* for the immediate declining phase (Mid-September 2022, Mid-November 2022), and *4* for the tail-end of the epidemic (Late November 2022 - Current).
[3] The models of interest include models of the spatial-wave framework (SW), and *n*-sub-epidemic frameworks (SE), a simple linear regression model, Facebook's Prophet model (Prophet), a general additive model, and an auto-regressive integrated moving average model (ARIMA). All models were calibrated with 11-weeks' worth of data at a time.
* The bolded values indicate the best performing model(s) at each time-period for each forecasting horizon. Here, *best performing* corresponds to the model which produces coverage closest to 95%.
** Each value is shown as a percentage (%).

**Table A4.** Average Weighted Interval Scores (WIS) of the forecasts generated for Brazil [1] (weeks of July 14th, 2022, through February 23rd, 2023) for each forecasting horizon (1-4 weeks), and time-period [2] of the epidemic, and averaged across all forecast periods (*Avg.*) for each model [3] of interest.

| Time Period | SW 1st Ranked | SW 2nd Ranked | SW Weighted Ensemble (2) | SW Unweighted Ensemble (2) | SE 1st Ranked | SE 2nd Ranked | SE Weighted Ensemble (2) | SE Unweighted Ensemble (2) | Simple Linear Regression | Prophet | General Additive Model | ARIMA[3] |
|---|---|---|---|---|---|---|---|---|---|---|---|---|
| **1-Week Forecasting Horizon** | | | | | | | | | | | | |
| 1 | 182.50 | 184.19 | 183.07 | 182.96 | 175.24 | **60.17** | 176.30 | 99.38 | 301.67 | 286.55 | 143.41 | 175.54 |
| 2 | 128.91 | 127.65 | 127.79 | 127.33 | 129.66 | 217.48 | 129.91 | 129.96 | 213.33 | 193.99 | 187.45 | **105.13** |
| 3 | 43.55 | 44.78 | 44.16 | 43.90 | 42.44 | 49.34 | 42.51 | **42.24** | 134.24 | 122.93 | 50.03 | 74.08 |
| 4 | 17.61 | 17.78 | 17.58 | 17.55 | **17.17** | 30.77 | 17.27 | 22.08 | 25.17 | 23.87 | 22.69 | 26.27 |
| Avg. | 62.50 | 62.85 | 62.51 | 62.30 | 57.38 | 80.23 | 57.54 | **56.38** | 130.32 | 120.40 | 78.69 | 74.51 |
| **2-Week Forecasting Horizon** | | | | | | | | | | | | |
| 1 | 287.23 | 292.39 | 287.55 | 285.98 | 423.81 | 508.78 | 423.51 | 442.52 | 360.46 | 348.22 | **261.61** | 269.45 |
| 2 | 134.23 | 133.08 | 133.31 | 133.25 | 129.63 | 291.18 | **129.19** | 152.84 | 280.13 | 261.53 | 296.06 | 134.78 |
| 3 | **42.18** | 42.77 | 42.48 | 42.28 | 43.03 | 49.99 | 42.84 | 42.96 | 152.43 | 141.04 | 62.71 | 89.59 |
| 4 | 16.62 | 17.15 | 16.90 | 16.86 | 16.72 | 27.95 | **16.60** | 20.43 | 24.83 | 23.37 | 21.89 | 25.56 |
| Avg. | 70.28 | 70.80 | 70.31 | 70.10 | 66.63 | 112.41 | **66.41** | 73.83 | 156.37 | 146.82 | 117.49 | 95.47 |
| **3-Week Forecasting Horizon** | | | | | | | | | | | | |
| 1 | 397.39 | 400.33 | 394.13 | 396.24 | 675.20 | 1130.26 | 674.84 | 822.48 | 396.88 | 387.49 | 361.54 | **322.83** |
| 2 | 144.66 | **143.24** | 143.70 | 143.78 | 144.26 | 297.20 | 144.65 | 161.81 | 338.15 | 323.87 | 344.67 | 161.79 |
| 3 | 41.45 | 41.56 | **41.13** | 41.45 | 42.11 | 51.67 | 41.99 | 43.30 | 167.73 | 158.20 | 76.59 | 98.23 |
| 4 | **16.47** | 16.81 | 16.58 | 16.66 | 16.61 | 27.87 | 16.66 | 20.53 | 27.10 | 25.73 | 22.15 | 27.30 |
| Avg. | 80.07 | 80.13 | 79.56 | 79.87 | **78.81** | 137.36 | 78.86 | 90.06 | 178.20 | 170.51 | 142.76 | 110.16 |
| **4-Week Forecasting Horizon** | | | | | | | | | | | | |
| 1 | 602.11 | 603.93 | 598.13 | 605.01 | 1096.32 | 2215.23 | 1095.91 | 1460.47 | 399.22 | **391.68** | 473.58 | 394.81 |
| 2 | 163.88 | 163.02 | 163.42 | 163.38 | 160.33 | 288.14 | **160.33** | 168.83 | 405.46 | 395.66 | 409.53 | 196.20 |
| 3 | 41.10 | 40.98 | **40.79** | 41.04 | 41.70 | 53.26 | 41.94 | 43.93 | 178.70 | 171.65 | 86.40 | 107.58 |
| 4 | **15.60** | 15.80 | 15.76 | 15.66 | 15.80 | 26.30 | 15.72 | 19.34 | 26.54 | 25.57 | 22.25 | 28.49 |
| Avg. | 98.38 | 98.35 | 97.94 | 98.48 | **97.53** | 175.54 | 97.58 | 115.04 | 195.96 | 190.39 | 171.19 | 128.36 |

[1] Data was obtained from the Our World in Data Team (OWID) [1].
[2] The time-period of the epidemic, where *1* refers to ascending phase of the epidemic (July 2022), *2* the peak (August-Early Sept. 2022), *3* for the immediate declining phase (Mid-September 2022, Mid-November 2022), and *4* for the tail-end of the epidemic (Late November 2022 - Current).
[3] The models of interest include models of the spatial-wave framework (SW), and *n*-sub-epidemic frameworks (SE), a simple linear regression model, Facebook's Prophet model (Prophet), a general additive model, and an auto-regressive integrated moving average model (ARIMA). All models were calibrated with 11-weeks' worth of data at a time.
* The bolded values indicate the best performing model(s) at each time-period for each forecasting horizon. Here, *best performing* refers to the model with the lowest metric value.

**Table A5.** Skills scores[1] for overall average forecast performance metrics (weeks of July 14th, 2022, through February 23rd, 2023) comparing the ARIMA[2] model to the *n*-sub-epidemic (SE) and spatial-wave (SW) frameworks across forecasting horizons (1-4 weeks) for Brazil.[3]

| **Baseline[4]** | ARIMA | | | | | | | |
|---|---|---|---|---|---|---|---|---|
| **Comparison[5]** | SW 1st Ranked | SW 2nd Ranked | SW Weighted Ensemble (2) | SW Unweighted Ensemble (2) | SE 1st Ranked | SE 2nd Ranked | SE Weighted Ensemble (2) | SE Unweighted Ensemble (2) |
| **Mean Squared Error (MSE)** | | | | | | | | |
| 1-Week | 16.68 | 15.49 | 14.22 | 16.81 | 26.76 | -46.97 | 26.76 | 36.02 |
| 2-Week | 20.34 | 21.81 | 20.68 | 21.35 | 29.02 | -124.47 | 29.02 | 12.77 |
| 3-Week | -0.66 | 1.96 | 0.13 | 3.39 | 8.46 | -208.40 | 8.46 | -54.14 |
| 4-Week | -49.08 | -44.90 | -47.94 | -47.38 | -37.23 | -485.68 | -37.23 | -189.22 |
| **Mean Absolute Error (MAE)** | | | | | | | | |
| 1-Week | 14.63 | 13.04 | 13.03 | 13.97 | 21.00 | -18.01 | 21.00 | 22.11 |
| 2-Week | 27.30 | 26.59 | 26.81 | 26.90 | 33.17 | -19.23 | 33.17 | 27.90 |
| 3-Week | 28.86 | 28.16 | 28.40 | 28.96 | 31.74 | -22.78 | 31.74 | 23.06 |
| 4-Week | 25.89 | 25.99 | 25.79 | 26.02 | 28.37 | -30.53 | 28.37 | 13.36 |
| **95% Prediction Interval Coverage (95% PI)** | | | | | | | | |
| 1-Week | 26.13 | 26.24 | 28.66 | 26.96 | 38.25 | 23.97 | 37.03 | 44.61 |
| 2-Week | 33.35 | 30.85 | 32.43 | 32.27 | 31.04 | -21.37 | 31.96 | 28.26 |
| 3-Week | 29.56 | 26.60 | 27.45 | 28.52 | 21.61 | -48.03 | 20.99 | 21.44 |
| 4-Week | 15.26 | 14.37 | 16.32 | 14.15 | 6.80 | -84.05 | 6.39 | 6.31 |
| **Weighted Interval Score (WIS)** | | | | | | | | |
| 1-Week | 16.12 | 15.65 | 16.11 | 16.39 | 22.99 | -7.67 | 22.78 | 24.33 |
| 2-Week | 26.38 | 25.84 | 26.35 | 26.57 | 30.20 | -17.75 | 30.44 | 22.66 |
| 3-Week | 27.31 | 27.26 | 27.78 | 27.50 | 28.46 | -24.70 | 28.41 | 18.24 |
| 4-Week | 23.36 | 23.38 | 23.70 | 23.28 | 24.02 | -36.76 | 23.98 | 10.38 |

[1] Skill scores are calculated by subtracting the mean metric score for the comparison model from the benchmark model, dividing by the mean metric score for the benchmark model and multiplying by 100. They are shown here as a percentage.
[2] Auto-regressive integrated moving average
[3] Data was obtained from the Our World in Data Team (OWID) [1].
[4] The ARIMA model used an 11-week calibration period for all forecasts.
[5] Comparison models used only a 11-week calibration period and were comprised of the 1st ranked, 2nd ranked, weighted (W) ensemble, and unweighted (UW) ensemble models from both the *n*-sub-epidemic (SE) and spatial-wave (SW) frameworks.
* Skill scores for 95% PI coverage are based on average Winkler scores.

**Table B1.** Average Mean Squared Error (MSE) of the forecasts generated for Canada [1] (weeks of July 28th, 2022, through February 23rd, 2023) for each forecasting horizon (1-4 weeks), and time-period [2] of the epidemic, and averaged across all forecast periods (*Avg.*) for each model [3] of interest.

| Time Period | SW 1st Ranked | SW 2nd Ranked | SW Weighted Ensemble (2) | SW Unweighted Ensemble (2) | SE 1st Ranked | SE 2nd Ranked | SE Weighted Ensemble (2) | SE Unweighted Ensemble (2) | Simple Linear Regression | Prophet | General Additive Model | ARIMA[3] |
|---|---|---|---|---|---|---|---|---|---|---|---|---|
| **1-Week Forecasting Horizon** | | | | | | | | | | | | |
| 1[4] | - | - | - | - | - | - | - | - | - | - | - | - |
| 2 | 1707.90 | 1708.18 | 1862.79 | 1586.23 | **717.78** | 1755.91 | **717.78** | 1238.67 | 1737.24 | 1740.63 | 2111.05 | 1376.00 |
| 3 | 128.96 | **113.12** | 119.67 | 120.50 | 122.73 | 440.67 | 122.73 | 227.80 | 1911.07 | 1914.87 | 336.74 | 1260.46 |
| 4 | 5.89 | 5.80 | 5.92 | 5.91 | 6.99 | **2.08** | 6.99 | 4.37 | 9.99 | 10.00 | 7.91 | 18.79 |
| Avg. | 228.64 | 222.61 | 241.16 | 212.86 | **103.23** | 299.66 | **103.23** | 180.31 | 909.77 | 911.57 | 350.20 | 630.17 |
| **2-Week Forecasting Horizon** | | | | | | | | | | | | |
| 1[4] | - | - | - | - | - | - | - | - | - | - | - | - |
| 2 | 1502.72 | 1511.51 | 1829.96 | 1667.22 | **713.43** | 1218.38 | **713.43** | 1099.27 | 2564.55 | 2567.19 | 1975.01 | 1116.17 |
| 3 | 131.88 | 131.12 | 130.10 | **129.23** | 134.86 | 912.04 | 134.86 | 488.36 | 2595.78 | 2601.69 | 331.42 | 1541.00 |
| 4 | 5.16 | 5.04 | 5.12 | 5.06 | 5.82 | 8.68 | 5.82 | **4.64** | 7.77 | 7.77 | 6.73 | 16.92 |
| Avg. | 208.15 | 208.71 | 241.30 | 224.11 | **107.06** | 449.98 | **107.06** | 272.86 | 1253.92 | 1256.44 | 333.50 | 708.74 |
| **3-Week Forecasting Horizon** | | | | | | | | | | | | |
| 1[4] | - | - | - | - | - | - | - | - | - | - | - | - |
| 2 | 2985.27 | 2938.38 | 2679.21 | 2726.92 | **865.69** | 2137.18 | **865.69** | 1921.37 | 5047.45 | 5049.80 | 4044.35 | 1922.44 |
| 3 | 126.64 | 124.49 | 127.27 | **124.08** | 128.83 | 1197.74 | 128.83 | 605.15 | 3207.44 | 3215.32 | 371.19 | 1852.80 |
| 4 | 4.92 | 4.91 | 5.01 | 4.97 | 5.57 | 9.46 | 5.57 | **4.89** | 7.05 | 7.05 | 6.35 | 16.15 |
| Avg. | 359.40 | 353.73 | 328.02 | 331.73 | **115.43** | 628.27 | **115.43** | 377.60 | 1742.41 | 1745.64 | 562.46 | 910.01 |
| **4-Week Forecasting Horizon** | | | | | | | | | | | | |
| 1[4] | - | - | - | - | - | - | - | - | - | - | - | - |
| 2 | 3881.76 | 3811.02 | 4326.06 | 4460.62 | **1062.82** | 2535.50 | **1062.82** | 2563.14 | 7810.72 | 7811.83 | 5504.61 | 2573.67 |
| 3 | 123.44 | 124.97 | 123.35 | **121.22** | 127.83 | 1636.03 | 127.83 | 720.56 | 3875.43 | 3885.27 | 327.52 | 2157.75 |
| 4 | 4.49 | 4.48 | 4.55 | **4.47** | 5.08 | 10.89 | 5.08 | 4.71 | 6.27 | 6.27 | 5.75 | 16.63 |
| Avg. | 450.71 | 443.96 | 496.67 | 509.74 | **128.86** | 829.67 | **128.86** | 468.68 | 2281.24 | 2285.09 | 696.65 | 1093.30 |

[1] Data was obtained from the Our World in Data Team (OWID) [1].
[2] The time-period of the epidemic, where *1* refers to ascending phases (June 2022), *2* the peak (July 2022), *3* for the immediate declining phase (August-Early October 2022), and *4* for the tail-end of the epidemic (Mid-October 2022 - Current).
[3] The models of interest include models of the spatial-wave framework (SW), and *n*-sub-epidemic frameworks (SE), a simple linear regression model, Facebook's Prophet model (Prophet), a general additive model, and an auto-regressive integrated moving average model (ARIMA). All models were calibrated with 11-weeks' worth of data at a time.
[4] Metrics are not available for the ascending phase of the epidemic in Canada, as it occurred in June 2022. Therefore, not enough observed weeks were available to conduct forecasts for the ascending phase of the epidemic.
* The bolded values indicate the best performing model(s) at each time-period for each forecasting horizon. Here, *best performing* refers to the model with the lowest metric value.

**Table B2.** Average Mean Absolute Error (MAE) of the forecasts generated for Canada [1] (weeks of July 28th, 2022, through February 23rd, 2023) for each forecasting horizon (1-4 weeks), and time-period [2] of the epidemic, and averaged across all forecast periods (*Avg.*) for each model [3] of interest.

| Time Period | SW 1st Ranked | SW 2nd Ranked | SW Weighted Ensemble (2) | SW Unweighted Ensemble (2) | SE 1st Ranked | SE 2nd Ranked | SE Weighted Ensemble (2) | SE Unweighted Ensemble (2) | Simple Linear Regression | Prophet | General Additive Model | ARIMA[3] |
|---|---|---|---|---|---|---|---|---|---|---|---|---|
| **1-Week Forecasting Horizon** | | | | | | | | | | | | |
| 1[4] | - | - | - | - | - | - | - | - | - | - | - | - |
| 2 | 38.03 | 37.68 | 40.18 | 36.98 | **24.76** | 41.87 | **24.76** | 33.82 | 32.22 | 32.22 | 42.56 | 34.67 |
| 3 | 10.18 | **9.43** | 9.75 | 9.76 | 10.03 | 16.34 | 10.03 | 13.02 | 31.25 | 31.28 | 14.27 | 28.25 |
| 4 | 1.68 | 1.67 | 1.68 | 1.69 | 1.70 | **1.08** | 1.70 | 1.51 | 1.70 | 1.71 | 1.64 | 3.42 |
| Avg. | 8.66 | 8.34 | 8.72 | 8.40 | **6.62** | 9.99 | **6.62** | 8.34 | 16.07 | 16.08 | 10.67 | 16.07 |
| **2-Week Forecasting Horizon** | | | | | | | | | | | | |
| 1[4] | - | - | - | - | - | - | - | - | - | - | - | - |
| 2 | 36.52 | 35.75 | 39.08 | 37.91 | **24.92** | 30.78 | **24.92** | 31.59 | 41.41 | 41.40 | 36.11 | 30.17 |
| 3 | 9.51 | 9.44 | **9.40** | 9.43 | 9.59 | 19.01 | 9.59 | 13.79 | 36.47 | 36.50 | 14.41 | 31.55 |
| 4 | 1.46 | 1.44 | 1.46 | 1.43 | 1.47 | 1.93 | 1.47 | 1.44 | 1.37 | 1.37 | **1.35** | 3.25 |
| Avg. | 8.14 | 8.02 | 8.36 | 8.24 | **6.34** | 10.70 | **6.34** | 8.45 | 18.83 | 18.84 | 9.90 | 16.77 |
| **3-Week Forecasting Horizon** | | | | | | | | | | | | |
| 1[4] | - | - | - | - | - | - | - | - | - | - | - | - |
| 2 | 45.52 | 44.08 | 43.58 | 44.15 | **26.84** | 38.72 | **26.84** | 34.73 | 59.60 | 59.59 | 49.25 | 36.44 |
| 3 | 9.34 | **9.20** | 9.27 | 9.22 | 9.41 | 21.01 | 9.41 | 14.86 | 39.45 | 39.50 | 14.91 | 34.04 |
| 4 | 1.37 | 1.36 | 1.38 | 1.37 | 1.38 | 1.93 | 1.38 | 1.46 | 1.27 | 1.27 | **1.25** | 3.15 |
| Avg. | 8.96 | 8.76 | 8.74 | 8.78 | **6.35** | 12.06 | **6.35** | 9.10 | 21.79 | 21.80 | 11.40 | 18.31 |
| **4-Week Forecasting Horizon** | | | | | | | | | | | | |
| 1[4] | - | - | - | - | - | - | - | - | - | - | - | - |
| 2 | 51.77 | 49.86 | 53.66 | 54.94 | **29.76** | 41.38 | **29.76** | 42.12 | 75.62 | 75.60 | 58.18 | 43.00 |
| 3 | 9.17 | 9.19 | 9.14 | **9.10** | 9.33 | 22.80 | 9.33 | 15.53 | 42.48 | 42.53 | 13.84 | 36.56 |
| 4 | 1.24 | 1.24 | 1.25 | 1.23 | 1.25 | 2.02 | 1.25 | 1.38 | 1.13 | 1.13 | **1.12** | 3.18 |
| Avg. | 9.48 | 9.29 | 9.66 | 9.77 | **6.46** | 13.00 | **6.46** | 9.85 | 24.52 | 24.54 | 11.85 | 19.96 |

[1] Data was obtained from the Our World in Data Team (OWID) [1].
[2] The time-period of the epidemic, where *1* refers to ascending phases (June 2022), *2* the peak (July 2022), *3* for the immediate declining phase (August-Early October 2022), and *4* for the tail-end of the epidemic (Mid-October 2022 - Current).
[3] The models of interest include models of the spatial-wave framework (SW), and *n*-sub-epidemic frameworks (SE), a simple linear regression model, Facebook's Prophet model (Prophet), a general additive model, and an auto-regressive integrated moving average model (ARIMA). All models were calibrated with 11-weeks' worth of data at a time.
[4] Metrics are not available for the ascending phase of the epidemic in Canada, as it occurred in June 2022. Therefore, not enough observed weeks were available to conduct forecasts for the ascending phase of the epidemic.
* The bolded values indicate the best performing model(s) at each time-period for each forecasting horizon. Here, *best performing* refers to the model with the lowest metric value.

**Table B3.** Average 95% Prediction Interval Coverage (95% PI Coverage) of the forecasts generated for Canada [1] (weeks of July 28th, 2022, through February 23rd, 2023) for each forecasting horizon (1-4 weeks), and time-period [2] of the epidemic, and averaged across all forecast periods (*Avg.*) for each model [3] of interest.

| Time Period | SW 1st Ranked | SW 2nd Ranked | SW Weighted Ensemble (2) | SW Unweighted Ensemble (2) | SE 1st Ranked | SE 2nd Ranked | SE Weighted Ensemble (2) | SE Unweighted Ensemble (2) | Simple Linear Regression | Prophet | General Additive Model | ARIMA[3] |
|---|---|---|---|---|---|---|---|---|---|---|---|---|
| **1-Week Forecasting Horizon** | | | | | | | | | | | | |
| 1[4] | - | - | - | - | - | - | - | - | - | - | - | - |
| 2 | **100.00** | **100.00** | **100.00** | **100.00** | **100.00** | **100.00** | **100.00** | **100.00** | 66.67 | 66.67 | 66.67 | **100.00** |
| 3 | 100.00 | 100.00 | 100.00 | 100.00 | 100.00 | 100.00 | 100.00 | 100.00 | 45.45 | 81.82 | 81.82 | **90.91** |
| 4 | **100.00** | **100.00** | **100.00** | **100.00** | **100.00** | **100.00** | **100.00** | **100.00** | 86.67 | **100.00** | **100.00** | **100.00** |
| Avg. | 100.00 | 100.00 | 100.00 | 100.00 | 100.00 | 100.00 | 100.00 | 100.00 | 68.97 | 89.66 | 89.66 | **96.55** |
| **2-Week Forecasting Horizon** | | | | | | | | | | | | |
| 1[4] | - | - | - | - | - | - | - | - | - | - | - | - |
| 2 | **100.00** | **100.00** | **100.00** | **100.00** | **100.00** | **100.00** | **100.00** | **100.00** | 50.00 | 50.00 | 66.67 | **100.00** |
| 3 | 100.00 | 100.00 | 100.00 | 100.00 | 100.00 | **95.45** | 100.00 | 100.00 | 45.45 | 68.18 | 81.82 | 90.91 |
| 4 | **100.00** | **100.00** | **100.00** | **100.00** | **100.00** | **100.00** | **100.00** | **100.00** | 86.67 | **100.00** | **100.00** | **100.00** |
| Avg. | 100.00 | 100.00 | 100.00 | 100.00 | 100.00 | 98.21 | 100.00 | 100.00 | 67.24 | 82.76 | 89.66 | **96.55** |
| **3-Week Forecasting Horizon** | | | | | | | | | | | | |
| 1[4] | - | - | - | - | - | - | - | - | - | - | - | - |
| 2 | **100.00** | **100.00** | **100.00** | **100.00** | **100.00** | 83.33 | **100.00** | **100.00** | 33.33 | 33.33 | 55.56 | **100.00** |
| 3 | 100.00 | 100.00 | 100.00 | 100.00 | 100.00 | **93.94** | 100.00 | 100.00 | 45.45 | 60.61 | 84.85 | 84.85 |
| 4 | 100.00 | 100.00 | 100.00 | 100.00 | 100.00 | 100.00 | 100.00 | 100.00 | 86.67 | **95.56** | 100.00 | 100.00 |
| Avg. | 100.00 | 100.00 | 100.00 | 100.00 | 100.00 | 96.43 | 100.00 | 100.00 | 65.52 | 75.86 | 89.66 | **94.25** |
| **4-Week Forecasting Horizon** | | | | | | | | | | | | |
| 1[4] | - | - | - | - | - | - | - | - | - | - | - | - |
| 2 | **100.00** | **100.00** | **100.00** | **100.00** | **100.00** | 75.00 | **100.00** | **100.00** | 25.00 | 25.00 | 50.00 | **100.00** |
| 3 | 100.00 | 100.00 | 100.00 | 100.00 | 100.00 | **93.18** | 100.00 | 100.00 | 43.18 | 54.55 | 84.09 | 84.09 |
| 4 | 100.00 | 100.00 | 100.00 | 100.00 | 100.00 | 100.00 | 100.00 | 100.00 | 86.67 | **93.33** | 100.00 | 100.00 |
| Avg. | 100.00 | 100.00 | 100.00 | 100.00 | 100.00 | **95.54** | 100.00 | 100.00 | 63.79 | 71.55 | 88.79 | 93.97 |

[1] Data was obtained from the Our World in Data Team (OWID) [1].

[2] The time-period of the epidemic, where *1* refers to ascending phases (June 2022), *2* the peak (July 2022), *3* for the immediate declining phase (August-Early October 2022), and *4* for the tail-end of the epidemic (Mid-October 2022 - Current).

[3] The models of interest include models of the spatial-wave framework (SW), and *n*-sub-epidemic frameworks (SE), a simple linear regression model, Facebook's Prophet model (Prophet), a general additive model, and an auto-regressive integrated moving average model (ARIMA). All models were calibrated with 11-weeks' worth of data at a time.

[4] Metrics are not available for the ascending phase of the epidemic in Canada, as it occurred in June 2022. Therefore, not enough observed weeks were available to conduct forecasts for the ascending phase of the epidemic.

* The bolded values indicate the best performing model(s) at each time-period for each forecasting horizon. Here, *best performing* corresponds to the model which produces coverage closest to 95%.

** Each value is shown as a percentage (%).

**Table B4.** Average Weighted Interval Scores (WIS) of the forecasts generated for Canada[1] (weeks of July 28th, 2022, through February 23rd, 2023) for each forecasting horizon (1-4 weeks), and time-period [2] of the epidemic, and averaged across all forecast periods (*Avg.*) for each model [3] of interest.

| Time Period | SW 1st Ranked | SW 2nd Ranked | SW Weighted Ensemble (2) | SW Unweighted Ensemble (2) | SE 1st Ranked | SE 2nd Ranked | SE Weighted Ensemble (2) | SE Unweighted Ensemble (2) | Simple Linear Regression | Prophet | General Additive Model | ARIMA[3] |
|---|---|---|---|---|---|---|---|---|---|---|---|---|
| **1-Week Forecasting Horizon** | | | | | | | | | | | | |
| 1[4] | - | - | - | - | - | - | - | - | - | - | - | - |
| 2 | 21.85 | 21.48 | 22.02 | 21.70 | 16.20 | 41.86 | **16.01** | 26.91 | 24.41 | 22.83 | 29.06 | 19.29 |
| 3 | 6.57 | **6.23** | 6.44 | 6.39 | 6.49 | 14.25 | 6.47 | 9.19 | 23.20 | 21.40 | 9.39 | 16.84 |
| 4 | 1.15 | 1.15 | 1.14 | 1.15 | 1.13 | 2.51 | 1.12 | 1.64 | 1.31 | 1.24 | **0.95** | 1.93 |
| Avg. | 5.35 | 5.18 | 5.31 | 5.26 | 4.31 | 9.93 | **4.29** | 6.41 | 12.00 | 11.12 | 7.06 | 9.38 |
| **2-Week Forecasting Horizon** | | | | | | | | | | | | |
| 1[4] | - | - | - | - | - | - | - | - | - | - | - | - |
| 2 | 21.33 | 20.77 | 21.21 | 21.07 | **16.27** | 45.24 | 16.30 | 28.12 | 31.69 | 29.84 | 27.68 | 18.28 |
| 3 | 6.49 | **6.39** | 6.48 | 6.46 | 6.55 | 17.97 | 6.52 | 10.49 | 27.54 | 25.83 | 9.54 | 18.50 |
| 4 | 1.13 | 1.12 | 1.11 | 1.13 | 1.08 | 2.81 | 1.09 | 1.74 | 1.12 | 1.07 | **0.94** | 2.05 |
| Avg. | 5.25 | 5.15 | 5.22 | 5.21 | 4.32 | 11.79 | **4.31** | 7.06 | 14.30 | 13.44 | 6.97 | 9.97 |
| **3-Week Forecasting Horizon** | | | | | | | | | | | | |
| 1[4] | - | - | - | - | - | - | - | - | - | - | - | - |
| 2 | 28.07 | 26.96 | 27.49 | 27.45 | **18.76** | 59.35 | 18.89 | 36.80 | 47.40 | 45.65 | 40.54 | 22.32 |
| 3 | 6.36 | **6.23** | 6.27 | 6.30 | 6.38 | 20.71 | 6.36 | 11.57 | 30.19 | 28.90 | 9.77 | 20.31 |
| 4 | 1.10 | 1.09 | 1.10 | 1.10 | 1.06 | 3.06 | 1.06 | 1.83 | 1.11 | 1.06 | **0.94** | 2.10 |
| Avg. | 5.89 | 5.72 | 5.79 | 5.80 | **4.41** | 14.01 | 4.42 | 8.15 | 16.93 | 16.23 | 8.38 | 11.09 |
| **4-Week Forecasting Horizon** | | | | | | | | | | | | |
| 1[4] | - | - | - | - | - | - | - | - | - | - | - | - |
| 2 | 32.20 | 30.67 | 31.22 | 31.66 | 20.72 | 74.10 | **20.62** | 44.95 | 61.72 | 60.44 | 48.74 | 25.50 |
| 3 | 6.16 | 6.14 | **6.13** | 6.16 | 6.24 | 24.15 | 6.24 | 12.61 | 32.90 | 31.93 | 9.69 | 21.94 |
| 4 | 1.07 | 1.07 | 1.07 | 1.07 | 1.03 | 3.39 | 1.04 | 1.95 | 1.03 | 1.01 | **0.92** | 2.23 |
| Avg. | 6.22 | 6.05 | 6.11 | 6.17 | 4.49 | 16.59 | **4.48** | 9.21 | 19.40 | 18.88 | 9.19 | 12.11 |

[1] Data was obtained from the Our World in Data Team (OWID) [1].
[2] The time-period of the epidemic, where *1* refers to ascending phases (June 2022), *2* the peak (July 2022), *3* for the immediate declining phase (August-Early October 2022), and *4* for the tail-end of the epidemic (Mid-October 2022 - Current).
[3] The models of interest include models of the spatial-wave framework (SW), and *n*-sub-epidemic frameworks (SE), a simple linear regression model, Facebook's Prophet model (Prophet), a general additive model, and an auto-regressive integrated moving average model (ARIMA). All models were calibrated with 11-weeks' worth of data at a time.
[4] Metrics are not available for the ascending phase of the epidemic in Canada, as it occurred in June 2022. Therefore, not enough observed weeks were available to conduct forecasts for the ascending phase of the epidemic.
* The bolded values indicate the best performing model(s) at each time-period for each forecasting horizon. Here, *best performing* refers to the model with the lowest metric value.

**Table B5.** Skills scores[1] for overall average forecast performance metrics (July 28th, 2022, through February 23rd, 2023) comparing the ARIMA[2] model to the *n*-sub-epidemic (SE) and spatial-wave (SW) frameworks across forecasting horizons (1-4 weeks) for Canada.[3]

| **Baseline**[4] | ARIMA | | | | | | | |
|---|---|---|---|---|---|---|---|---|
| **Comparison**[5] | SW 1st Ranked | SW 2nd Ranked | SW Weighted Ensemble (2) | SW Unweighted Ensemble (2) | SE 1st Ranked | SE 2nd Ranked | SE Weighted Ensemble (2) | SE Unweighted Ensemble (2) |
| **Mean Squared Error (MSE)** | | | | | | | | |
| 1-Week | 63.72 | 64.67 | 61.73 | 66.22 | 83.62 | 52.45 | 83.62 | 71.39 |
| 2-Week | 70.63 | 70.55 | 65.95 | 68.38 | 84.89 | 36.51 | 84.89 | 61.50 |
| 3-Week | 60.51 | 61.13 | 63.95 | 63.55 | 87.32 | 30.96 | 87.32 | 58.51 |
| 4-Week | 58.78 | 59.39 | 54.57 | 53.38 | 88.21 | 24.11 | 88.21 | 57.13 |
| **Mean Absolute Error (MAE)** | | | | | | | | |
| 1-Week | 46.09 | 48.12 | 45.72 | 47.74 | 58.80 | 37.83 | 58.80 | 48.09 |
| 2-Week | 51.47 | 52.17 | 50.14 | 50.84 | 62.20 | 36.19 | 62.20 | 49.63 |
| 3-Week | 51.08 | 52.19 | 52.29 | 52.07 | 65.31 | 34.16 | 65.31 | 50.30 |
| 4-Week | 52.52 | 53.47 | 51.58 | 51.04 | 67.63 | 34.88 | 67.63 | 50.65 |
| **95% Prediction Interval Coverage (95% PI)** * | | | | | | | | |
| 1-Week | 27.55 | 27.50 | 27.84 | 27.55 | 34.56 | -470.63 | 34.61 | -219.38 |
| 2-Week | 31.41 | 31.02 | 31.26 | 30.95 | 39.33 | -561.30 | 39.55 | -229.91 |
| 3-Week | 37.33 | 37.00 | 37.46 | 36.93 | 46.79 | -641.29 | 46.90 | -277.38 |
| 4-Week | 40.09 | 39.68 | 39.94 | 39.80 | 51.17 | -763.62 | 51.38 | -310.15 |
| **Weighted Interval Score (WIS)** | | | | | | | | |
| 1-Week | 43.01 | 44.79 | 43.37 | 43.92 | 54.06 | -5.83 | 54.30 | 31.67 |
| 2-Week | 47.35 | 48.35 | 47.60 | 47.73 | 56.71 | -18.31 | 56.80 | 29.14 |
| 3-Week | 46.95 | 48.47 | 47.79 | 47.75 | 60.21 | -26.30 | 60.20 | 26.53 |
| 4-Week | 48.63 | 50.03 | 49.58 | 49.10 | 62.97 | -36.98 | 63.04 | 24.01 |

[1] Skill scores are calculated by subtracting the mean metric score for the comparison model from the benchmark model, dividing by the mean metric score for the benchmark model and multiplying by 100. They are shown here as a percentage.
[2] Auto-regressive integrated moving average
[3] Data was obtained from the Our World in Data Team (OWID) [1].
[4] The ARIMA model used an 11-week calibration period for all forecasts.
[5] Comparison models used only a 11-week calibration period and were comprised of the 1st ranked, 2nd ranked, weighted (W) ensemble, and unweighted (UW) ensemble models from both the *n*-sub-epidemic (SE) and spatial-wave (SW) frameworks.
* Skill scores for 95% PI coverage are based on average Winkler scores.

**Table C1.** Average Mean Squared Error (MSE) of the forecasts generated for France [1] (weeks of July 14th, 2022, through February 23rd, 2023) for each forecasting horizon (1-4 weeks), and time-period [2] of the epidemic, and averaged across all forecast periods (*Avg.*) for each model [3] of interest.

| Time Period | SW 1st Ranked | SW 2nd Ranked | SW Weighted Ensemble (2) | SW Unweighted Ensemble (2) | SE 1st Ranked | SE 2nd Ranked | SE Weighted Ensemble (2) | SE Unweighted Ensemble (2) | Simple Linear Regression | Prophet | General Additive Model | ARIMA[3] |
|---|---|---|---|---|---|---|---|---|---|---|---|---|
| **1-Week Forecasting Horizon** | | | | | | | | | | | | |
| 1 | 246033.94 | 250782.70 | 250123.34 | 248718.16 | 248613.15 | 250374.80 | 248613.15 | 252100.09 | 197927.92 | 198081.95 | **180233.56** | 197936.01 |
| 2 | 117624.23 | 116710.54 | 117841.20 | 117396.10 | 106837.12 | 861452.10 | 106837.12 | 392457.20 | 60865.79 | 60736.10 | 243944.46 | **43125.81** |
| 3 | 2402.32 | 1844.29 | 2281.62 | 2417.19 | **1561.20** | 5191.84 | **1561.20** | 2447.77 | 25813.65 | 25840.38 | 10119.64 | 32767.11 |
| 4 | 119.81 | 19.79 | 19.39 | 19.33 | 19.26 | 44.49 | 19.26 | 22.27 | **19.24** | 19.24 | 60.54 | 26.80 |
| Avg. | 33461.82 | 33250.10 | 33566.65 | 33458.79 | 31063.90 | 188141.30 | 31063.90 | 90493.51 | 25658.85 | 25643.78 | 59160.20 | **23671.13** |
| **2-Week Forecasting Horizon** | | | | | | | | | | | | |
| 1 | 182652.43 | 185723.04 | 183212.94 | 184343.18 | 184954.73 | 188324.37 | 184954.73 | 186179.85 | 131652.88 | 131780.75 | **112472.74** | 132198.42 |
| 2 | 277041.18 | 252257.71 | 261969.29 | 257548.43 | 237730.96 | 1546299.63 | 237730.96 | 982268.75 | 72615.48 | 72412.28 | 380808.52 | **47947.66** |
| 3 | 1919.06 | 1450.58 | 1837.46 | 1938.43 | **1348.91** | 4409.00 | **1348.91** | 2015.89 | 29223.27 | 29233.70 | 9875.30 | 35192.08 |
| 4 | 99.55 | 21.55 | 21.36 | 21.52 | 21.08 | 36.59 | 21.08 | 21.37 | **19.34** | 19.34 | 89.91 | 25.67 |
| Avg. | 64131.94 | 58956.78 | 60972.80 | 60121.57 | 55899.96 | 327501.17 | 55899.96 | 210145.66 | 26627.54 | 26592.43 | 85096.54 | **22986.69** |
| **3-Week Forecasting Horizon** | | | | | | | | | | | | |
| 1 | 141680.59 | 143743.91 | 142425.37 | 148732.04 | 144237.62 | 147929.21 | 144237.62 | 148523.37 | 92291.96 | 92394.66 | **75761.54** | 92988.45 |
| 2 | 724447.39 | 640035.63 | 680464.09 | 686317.39 | 572771.23 | 1232821.09 | 572771.23 | 885119.31 | 90339.19 | 90068.78 | 597284.54 | **54001.16** |
| 3 | 1656.82 | 1356.75 | 1618.85 | 1674.76 | **1235.86** | 3741.80 | **1235.86** | 1659.54 | 33684.44 | 33685.25 | 11700.49 | 37603.13 |
| 4 | 80.55 | 22.76 | 23.27 | 22.61 | 22.34 | 31.75 | 22.34 | 19.29 | 19.12 | **19.11** | 125.31 | 22.67 |
| Avg. | 155212.79 | 137717.11 | 146099.68 | 147541.34 | 123787.97 | 261087.06 | 123787.97 | 188660.18 | 30013.96 | 29961.75 | 129077.65 | **23467.50** |
| **4-Week Forecasting Horizon** | | | | | | | | | | | | |
| 1 | 118362.98 | 119876.82 | 120500.37 | 117009.68 | 121007.21 | 125183.30 | 121007.21 | 121991.58 | 69902.56 | 69986.14 | **57215.63** | 70599.82 |
| 2 | 2038123.32 | 1759961.72 | 1812804.84 | 1891560.17 | 1503936.00 | 932035.63 | 1503936.00 | 598411.55 | 115810.11 | 115468.98 | 851943.63 | **60520.07** |
| 3 | 1342.95 | 1153.49 | 1297.19 | 1364.79 | **1013.13** | 3231.80 | **1013.13** | 1057.26 | 36015.44 | 36009.40 | 12437.84 | 39133.34 |
| 4 | 70.72 | 24.08 | 24.26 | 23.75 | 23.42 | 29.92 | 23.42 | **18.45** | 18.75 | 18.75 | 165.86 | 21.83 |
| Avg. | 426122.91 | 368554.58 | 379543.92 | 395733.81 | 315588.50 | 197947.19 | 315588.50 | 128280.64 | 35074.23 | 35005.07 | 181325.18 | **24413.14** |

[1] Data was obtained from the Our World in Data Team (OWID) [1].
[2] The time-period of the epidemic, where *1* refers to ascending phases (May - Mid July 2022), *2* the peak (Late July – Late August 2022), *3* for the immediate declining phase (Early September–Mid-October 2022), and *4* for the tail-end of the epidemic (Late October 2022 - Current).
[3] The models of interest include models of the spatial-wave framework (SW), and *n*-sub-epidemic frameworks (SE), a simple linear regression model, Facebook's Prophet model (Prophet), a general additive model, and an auto-regressive integrated moving average model (ARIMA). All models were calibrated with 11-weeks' worth of data at a time.
* The bolded values indicate the best performing model(s) at each time-period for each forecasting horizon. Here, *best performing* refers to the model with the lowest metric value.

**Table C2.** Average Mean Absolute Error (MAE) of the forecasts generated for France [1] (weeks of July 14th, 2022, through February 23rd, 2023) for each forecasting horizon (1-4 weeks), and time-period [2] of the epidemic, and averaged across all forecast periods (*Avg.*) for each model [3] of interest.

| Time Period | SW 1st Ranked | SW 2nd Ranked | SW Weighted Ensemble (2) | SW Unweighted Ensemble (2) | SE 1st Ranked | SE 2nd Ranked | SE Weighted Ensemble (2) | SE Unweighted Ensemble (2) | Simple Linear Regression | Prophet | General Additive Model | ARIMA[3] |
|---|---|---|---|---|---|---|---|---|---|---|---|---|
| **1-Week Forecasting Horizon** | | | | | | | | | | | | |
| 1 | 496.02 | 500.78 | 500.12 | 498.72 | 498.61 | 500.37 | 498.61 | 502.10 | 444.89 | 445.06 | **424.54** | 444.90 |
| 2 | 272.68 | 284.03 | 284.00 | 282.70 | 262.57 | 669.15 | 262.57 | 448.40 | 224.05 | 223.86 | 422.89 | **175.74** |
| 3 | 42.80 | 35.25 | 40.76 | 42.85 | 33.32 | 51.44 | 33.32 | **33.09** | 119.69 | 120.07 | 69.00 | 141.83 |
| 4 | 4.89 | 2.54 | **2.46** | 2.48 | 2.54 | 4.59 | 2.54 | 2.63 | 2.61 | 2.61 | 4.39 | 3.56 |
| Avg. | 86.38 | 85.86 | 87.11 | 87.31 | **80.87** | 170.49 | **80.87** | 119.43 | 91.94 | 91.99 | 121.06 | 87.78 |
| **2-Week Forecasting Horizon** | | | | | | | | | | | | |
| 1 | 420.69 | 424.08 | 421.38 | 422.45 | 423.48 | 427.92 | 423.48 | 425.01 | 350.29 | 350.48 | **318.00** | 351.35 |
| 2 | 356.44 | 361.25 | 357.36 | 356.80 | 335.04 | 772.79 | 335.04 | 581.30 | 239.43 | 239.14 | 490.35 | **195.35** |
| 3 | 36.63 | 29.07 | 35.13 | 37.07 | **28.51** | 46.01 | **28.51** | 30.89 | 121.19 | 121.21 | 62.69 | 149.47 |
| 4 | 4.82 | 2.68 | 2.65 | 2.66 | 2.72 | 4.06 | 2.72 | **2.61** | 2.61 | 2.61 | 5.26 | 3.56 |
| Avg. | 99.59 | 97.77 | 98.32 | 98.71 | 92.21 | 187.85 | 92.21 | 143.73 | 92.22 | 92.17 | 130.27 | **90.45** |
| **3-Week Forecasting Horizon** | | | | | | | | | | | | |
| 1 | 361.96 | 364.25 | 362.87 | 371.42 | 365.86 | 371.58 | 365.86 | 372.62 | 272.36 | 272.56 | **228.12** | 274.47 |
| 2 | 467.22 | 468.91 | 471.96 | 466.13 | 430.29 | 684.30 | 430.29 | 562.52 | 265.25 | 264.90 | 593.01 | **211.49** |
| 3 | 31.62 | 26.96 | 30.81 | 31.98 | **25.48** | 42.78 | **25.48** | 27.97 | 124.49 | 124.53 | 62.80 | 155.97 |
| 4 | 4.46 | 2.69 | 2.70 | 2.72 | 2.77 | 3.57 | 2.77 | **2.45** | 2.54 | 2.54 | 5.89 | 3.23 |
| Avg. | 119.08 | 117.47 | 118.99 | 118.37 | 109.23 | 166.56 | 109.23 | 137.25 | 95.63 | 95.58 | 148.77 | **92.54** |
| **4-Week Forecasting Horizon** | | | | | | | | | | | | |
| 1 | 326.47 | 328.15 | 329.51 | 324.05 | 331.04 | 338.36 | 331.04 | 332.35 | 217.34 | 217.55 | **181.02** | 220.50 |
| 2 | 633.76 | 602.31 | 609.92 | 619.27 | 567.43 | 538.22 | 567.43 | 443.09 | 298.30 | 297.92 | 654.17 | **222.19** |
| 3 | 27.05 | 24.72 | 26.27 | 27.51 | 22.80 | 39.32 | 22.80 | **22.31** | 124.51 | 124.57 | 62.01 | 160.01 |
| 4 | 4.17 | 2.68 | 2.67 | 2.65 | 2.73 | 3.40 | 2.73 | **2.30** | 2.43 | 2.43 | 6.32 | 3.16 |
| Avg. | 151.07 | 143.28 | 145.28 | 147.31 | 135.73 | 134.27 | 135.73 | 109.71 | 100.52 | 100.47 | 159.82 | **93.83** |

[1] Data was obtained from the Our World in Data Team (OWID) [1].
[2] The time-period of the epidemic, where *1* refers to ascending phases (May - Mid July 2022), *2* the peak (Late July – Late August 2022), *3* for the immediate declining phase (Early September–Mid-October 2022), and *4* for the tail-end of the epidemic (Late October 2022 - Current).
[3] The models of interest include models of the spatial-wave framework (SW), and *n*-sub-epidemic frameworks (SE), a simple linear regression model, Facebook's Prophet model (Prophet), a general additive model, and an auto-regressive integrated moving average model (ARIMA). All models were calibrated with 11-weeks' worth of data at a time.
* The bolded values indicate the best performing model(s) at each time-period for each forecasting horizon. Here, *best performing* refers to the model with the lowest metric value.

**Table C3.** Average 95% Prediction Interval Coverage (95% PI Coverage) of the forecasts generated for France [1] (weeks of July 14th, 2022, through February 23rd, 2023) for each forecasting horizon (1-4 weeks), and time-period [2] of the epidemic, and averaged across all forecast periods (*Avg.*) for each model [3] of interest.

| Time Period | SW 1st Ranked | SW 2nd Ranked | SW Weighted Ensemble (2) | SW Unweighted Ensemble (2) | SE 1st Ranked | SE 2nd Ranked | SE Weighted Ensemble (2) | SE Unweighted Ensemble (2) | Simple Linear Regression | Prophet | General Additive Model | ARIMA[3] |
|---|---|---|---|---|---|---|---|---|---|---|---|---|
| **1-Week Forecasting Horizon** | | | | | | | | | | | | |
| 1 | 0.00 | 0.00 | 0.00 | 0.00 | 0.00 | **100.00** | 0.00 | 0.00 | 0.00 | 0.00 | 0.00 | 0.00 |
| 2 | 66.67 | 66.67 | 66.67 | 66.67 | 66.67 | 50.00 | 66.67 | 66.67 | 33.33 | 66.67 | 33.33 | **83.33** |
| 3 | **100.00** | **100.00** | **100.00** | **100.00** | **100.00** | **100.00** | **100.00** | **100.00** | 85.71 | **100.00** | **100.00** | **100.00** |
| 4 | 86.67 | **93.33** | **93.33** | **93.33** | **93.33** | 86.67 | **93.33** | **93.33** | 73.33 | **93.33** | 80.00 | **93.33** |
| Avg. | 82.76 | 86.21 | 86.21 | 86.21 | 86.21 | 82.76 | 86.21 | 86.21 | 65.52 | 86.21 | 72.41 | **89.66** |
| **2-Week Forecasting Horizon** | | | | | | | | | | | | |
| 1 | 0.00 | 0.00 | 0.00 | 0.00 | 0.00 | **100.00** | 0.00 | 50.00 | 0.00 | 0.00 | 0.00 | 0.00 |
| 2 | 75.00 | 75.00 | 75.00 | 75.00 | 66.67 | 50.00 | 66.67 | 66.67 | 33.33 | 58.33 | 41.67 | **91.67** |
| 3 | **100.00** | **100.00** | **100.00** | **100.00** | **100.00** | **100.00** | **100.00** | **100.00** | 85.71 | **100.00** | **100.00** | **100.00** |
| 4 | 86.67 | **93.33** | **93.33** | **93.33** | **93.33** | 90.00 | **93.33** | **93.33** | 73.33 | 86.67 | 80.00 | **93.33** |
| Avg. | 84.48 | 87.93 | 87.93 | 87.93 | 86.21 | 84.48 | 86.21 | 87.93 | 65.52 | 81.03 | 74.14 | **91.38** |
| **3-Week Forecasting Horizon** | | | | | | | | | | | | |
| 1 | 0.00 | 0.00 | 0.00 | 0.00 | 0.00 | **100.00** | 0.00 | 66.67 | 0.00 | 0.00 | 33.33 | 0.00 |
| 2 | 77.78 | 77.78 | 77.78 | 77.78 | 77.78 | 50.00 | 72.22 | 72.22 | 33.33 | 50.00 | 44.44 | **94.44** |
| 3 | 100.00 | 100.00 | 100.00 | 100.00 | 100.00 | 100.00 | 100.00 | 100.00 | 85.71 | **95.24** | 100.00 | 100.00 |
| 4 | 86.67 | **93.33** | **93.33** | **93.33** | **93.33** | 91.11 | **93.33** | **93.33** | 75.56 | 86.67 | 82.22 | **93.33** |
| Avg. | 85.06 | 88.51 | 88.51 | 88.51 | 88.51 | 85.06 | 87.36 | 89.66 | 66.67 | 78.16 | 77.01 | **91.95** |
| **4-Week Forecasting Horizon** | | | | | | | | | | | | |
| 1 | 0.00 | 25.00 | 0.00 | 0.00 | 0.00 | **100.00** | 0.00 | 75.00 | 0.00 | 0.00 | 50.00 | 25.00 |
| 2 | 79.17 | 79.17 | 79.17 | 79.17 | 75.00 | 58.33 | 75.00 | 75.00 | 29.17 | 41.67 | 50.00 | **95.83** |
| 3 | 100.00 | 100.00 | 100.00 | 100.00 | 100.00 | 100.00 | 100.00 | 100.00 | 85.71 | **92.86** | 100.00 | 100.00 |
| 4 | 86.67 | **93.33** | **93.33** | **93.33** | **93.33** | **93.33** | **93.33** | **93.33** | 78.33 | 83.33 | 83.33 | **93.33** |
| Avg. | 85.34 | 89.66 | 88.79 | 88.79 | 87.93 | 87.93 | 87.93 | 90.52 | 67.24 | 74.14 | 79.31 | **93.10** |

[1] Data was obtained from the Our World in Data Team (OWID) [1].
[2] The time-period of the epidemic, where *1* refers to ascending phases (May - Mid July 2022), *2* the peak (Late July – Late August 2022), *3* for the immediate declining phase (Early September–Mid-October 2022), and *4* for the tail-end of the epidemic (Late October 2022 - Current).
[3] The models of interest include models of the spatial-wave framework (SW), and *n*-sub-epidemic frameworks (SE), a simple linear regression model, Facebook's Prophet model (Prophet), a general additive model, and an auto-regressive integrated moving average model (ARIMA). All models were calibrated with 11-weeks' worth of data at a time.
* The bolded values indicate the best performing model(s) at each time-period for each forecasting horizon. Here, *best performing* corresponds to the model which produces coverage closest to 95%.
** Each value is shown as a percentage (%).

**Table C4.** Average Weighted Interval Scores (WIS) of the forecasts generated for France[1] (weeks of July 14th, 2022, through February 23rd, 2023) for each forecasting horizon (1-4 weeks), and time-period [2] of the epidemic, and averaged across all forecast periods (*Avg.*) for each model [3] of interest.

| Time Period | SW 1st Ranked | SW 2nd Ranked | SW Weighted Ensemble (2) | SW Unweighted Ensemble (2) | SE 1st Ranked | SE 2nd Ranked | SE Weighted Ensemble (2) | SE Unweighted Ensemble (2) | Simple Linear Regression | Prophet | General Additive Model | ARIMA[3] |
|---|---|---|---|---|---|---|---|---|---|---|---|---|
| **1-Week Forecasting Horizon** | | | | | | | | | | | | |
| 1 | 461.09 | 460.24 | 461.02 | 461.03 | 463.31 | **338.19** | 463.27 | 379.18 | 431.73 | 426.71 | 394.78 | 420.47 |
| 2 | 192.43 | 206.82 | 200.34 | 198.40 | 180.51 | 509.71 | 181.26 | 271.09 | 160.13 | 144.33 | 314.33 | **105.91** |
| 3 | **36.26** | 39.19 | 37.32 | 36.80 | 36.92 | 57.73 | 36.78 | 44.56 | 74.13 | 69.42 | 43.41 | 87.78 |
| 4 | 5.30 | 3.25 | 3.80 | 3.76 | 4.93 | 5.58 | 5.00 | 4.77 | **2.36** | 2.54 | 4.33 | 7.01 |
| Avg. | 67.21 | 69.80 | 68.32 | 67.77 | 64.78 | 133.94 | 64.94 | 82.39 | 67.13 | 62.65 | 91.36 | **61.23** |
| **2-Week Forecasting Horizon** | | | | | | | | | | | | |
| 1 | 378.52 | 376.28 | 377.68 | 376.35 | 380.62 | **275.75** | 380.77 | 309.60 | 336.26 | 332.68 | 277.96 | 321.86 |
| 2 | 243.61 | 251.91 | 246.19 | 248.24 | 224.45 | 642.78 | 224.58 | 342.13 | 173.34 | 160.38 | 363.83 | **112.75** |
| 3 | **34.75** | 37.98 | 36.13 | 35.78 | 35.71 | 56.00 | 36.29 | 43.69 | 76.50 | 72.65 | 41.19 | 93.38 |
| 4 | 5.02 | 3.29 | 3.72 | 3.75 | 4.87 | 5.34 | 4.98 | 4.58 | **2.34** | 2.44 | 5.33 | 8.08 |
| Avg. | 74.44 | 75.97 | 74.61 | 74.91 | 70.71 | 158.78 | 70.93 | 94.38 | 67.13 | 63.45 | 97.56 | **61.14** |
| **3-Week Forecasting Horizon** | | | | | | | | | | | | |
| 1 | 312.37 | 308.75 | 311.46 | 310.57 | 315.35 | 232.74 | 315.34 | 261.14 | 257.44 | 254.62 | **194.87** | 241.01 |
| 2 | 311.57 | 322.24 | 317.28 | 315.21 | 284.73 | 589.21 | 285.41 | 361.28 | 195.98 | 184.69 | 445.73 | **119.52** |
| 3 | **33.84** | 37.88 | 35.13 | 35.08 | 35.26 | 56.03 | 35.61 | 43.03 | 79.05 | 75.39 | 41.03 | 98.61 |
| 4 | 4.75 | 3.26 | 3.69 | 3.67 | 4.96 | 5.14 | 4.94 | 4.45 | **2.24** | 2.30 | 6.08 | 8.95 |
| Avg. | 85.86 | 88.15 | 86.78 | 86.29 | 80.86 | 146.11 | 81.08 | 96.44 | 69.66 | 66.38 | 111.99 | **61.47** |
| **4-Week Forecasting Horizon** | | | | | | | | | | | | |
| 1 | 269.37 | 265.16 | 268.08 | 267.03 | 273.06 | 206.74 | 272.93 | 229.39 | 201.64 | 199.50 | **153.88** | 188.54 |
| 2 | 412.49 | 412.06 | 410.52 | 409.63 | 367.38 | 479.00 | 367.20 | 336.13 | 225.95 | 218.92 | 496.87 | **125.93** |
| 3 | **32.44** | 36.72 | 34.17 | 33.75 | 34.40 | 56.15 | 34.42 | 42.39 | 79.31 | 76.99 | 40.01 | 102.31 |
| 4 | 4.60 | 3.26 | 3.61 | 3.61 | 5.03 | 5.10 | 5.01 | 4.47 | **2.13** | 2.19 | 6.85 | 9.72 |
| Avg. | 104.84 | 104.95 | 104.29 | 103.98 | 96.33 | 122.43 | 96.28 | 90.00 | 73.95 | 71.89 | 121.30 | **62.28** |

[1] Data was obtained from the Our World in Data Team (OWID) [1].
[2] The time-period of the epidemic, where *1* refers to ascending phases (May - Mid July 2022), *2* the peak (Late July – Late August 2022), *3* for the immediate declining phase (Early September–Mid-October 2022), and *4* for the tail-end of the epidemic (Late October 2022 - Current).
[3] The models of interest include models of the spatial-wave framework (SW), and *n*-sub-epidemic frameworks (SE), a simple linear regression model, Facebook's Prophet model (Prophet), a general additive model, and an auto-regressive integrated moving average model (ARIMA). All models were calibrated with 11-weeks' worth of data at a time.
* The bolded values indicate the best performing model(s) at each time-period for each forecasting horizon. Here, *best performing* refers to the model with the lowest metric value.

**Table C5.** Skills scores[1] for overall average forecast performance metrics (weeks of July 14th, 2022, through February 23rd, 2023) comparing the ARIMA[2] model to the *n*-sub-epidemic (SE) and spatial-wave (SW) frameworks across forecasting horizons (1-4 weeks) for France.[3]

| **Baseline[4]** | ARIMA | | | | | | | |
|---|---|---|---|---|---|---|---|---|
| **Comparison[5]** | SW 1st Ranked | SW 2nd Ranked | SW Weighted Ensemble (2) | SW Unweighted Ensemble (2) | SE 1st Ranked | SE 2nd Ranked | SE Weighted Ensemble (2) | SE Unweighted Ensemble (2) |
| **Mean Squared Error (MSE)** | | | | | | | | |
| 1-Week | -41.36 | -40.47 | -41.80 | -41.35 | -31.23 | -694.81 | -31.23 | -282.29 |
| 2-Week | -179.00 | -156.48 | -165.25 | -161.55 | -143.18 | -1324.74 | -143.18 | -814.21 |
| 3-Week | -561.39 | -486.84 | -522.56 | -528.71 | -427.49 | -1012.55 | -427.49 | -703.92 |
| 4-Week | -1645.47 | -1409.66 | -1454.67 | -1520.99 | -1192.70 | -710.82 | -1192.70 | -425.46 |
| **Mean Absolute Error (MAE)** | | | | | | | | |
| 1-Week | 1.59 | 2.19 | 0.76 | 0.53 | 7.86 | -94.23 | 7.86 | -36.07 |
| 2-Week | -10.10 | -8.09 | -8.69 | -9.13 | -1.94 | -107.68 | -1.94 | -58.90 |
| 3-Week | -28.68 | -26.94 | -28.58 | -27.91 | -18.03 | -79.98 | -18.03 | -48.31 |
| 4-Week | -61.00 | -52.71 | -54.83 | -57.00 | -44.66 | -43.10 | -44.66 | -16.92 |
| **95% Prediction Interval Coverage (95% PI) *** | | | | | | | | |
| 1-Week | -51.64 | -46.53 | -47.45 | -48.53 | -43.22 | -256.10 | -44.74 | -11.34 |
| 2-Week | -40.49 | -36.96 | -37.53 | -39.77 | -32.90 | -374.25 | -30.73 | -26.65 |
| 3-Week | -59.65 | -51.05 | -54.90 | -53.57 | -44.09 | -388.56 | -42.78 | -47.96 |
| 4-Week | -105.18 | -92.84 | -96.21 | -96.93 | -81.88 | -410.38 | -82.20 | -87.95 |
| **Weighted Interval Score (WIS)** | | | | | | | | |
| 1-Week | -9.76 | -14.00 | -11.58 | -10.69 | -5.81 | -118.75 | -6.06 | -34.56 |
| 2-Week | -21.75 | -24.24 | -22.02 | -22.52 | -15.64 | -159.68 | -16.01 | -54.35 |
| 3-Week | -39.67 | -43.39 | -41.16 | -40.37 | -31.54 | -137.69 | -31.89 | -56.88 |
| 4-Week | -68.34 | -68.52 | -67.47 | -66.96 | -54.68 | -96.58 | -54.60 | -44.51 |

[1] Skill scores are calculated by subtracting the mean metric score for the comparison model from the benchmark model, dividing by the mean metric score for the benchmark model and multiplying by 100. They are shown here as a percentage.
[2] Auto-regressive integrated moving average
[3] Data was obtained from the Our World in Data Team (OWID) [1].
[4] The ARIMA model used an 11-week calibration period for all forecasts.
[5] Comparison models used only a 11-week calibration period and were comprised of the 1st ranked, 2nd ranked, weighted (W) ensemble, and unweighted (UW) ensemble models from both the *n*-sub-epidemic (SE) and spatial-wave (SW) frameworks.
* Skill scores for 95% PI coverage are based on average Winkler scores.

**Table D1.** Average Mean Squared Error (MSE) of the forecasts generated for Germany [1] (weeks of July 14th, 2022, through February 23rd, 2023) for each forecasting horizon (1-4 weeks), and time-period [2] of the epidemic, and averaged across all forecast periods (*Avg.*) for each model [3] of interest.

| Time Period | SW 1st Ranked | SW 2nd Ranked | SW Weighted Ensemble (2) | SW Unweighted Ensemble (2) | SE 1st Ranked | SE 2nd Ranked | SE Weighted Ensemble (2) | SE Unweighted Ensemble (2) | Simple Linear Regression | Prophet | General Additive Model | ARIMA[3] |
|---|---|---|---|---|---|---|---|---|---|---|---|---|
| **1-Week Forecasting Horizon** | | | | | | | | | | | | |
| 1 | - | - | - | - | - | - | - | - | - | - | - | - |
| 2 | 499.96 | 476.08 | 492.08 | 472.54 | 434.72 | 241.01 | 434.72 | **130.42** | 24087.04 | 24144.24 | 3230.12 | 11793.96 |
| 3 | **1201.32** | 1261.47 | 1264.91 | 1267.97 | 1221.16 | 3259.66 | 1221.16 | 1383.41 | 18218.57 | 18192.54 | 3052.90 | 5317.09 |
| 4 | 8.11 | 9.48 | 9.33 | 9.16 | 9.57 | 8.63 | 9.57 | 8.57 | 9.27 | 9.26 | 162.98 | **7.83** |
| Avg. | **559.96** | 586.81 | 588.82 | 589.43 | 567.35 | 1474.00 | 567.35 | 629.08 | 9002.33 | 8992.63 | 1564.23 | 2794.26 |
| **2-Week Forecasting Horizon** | | | | | | | | | | | | |
| 1 | - | - | - | - | - | - | - | - | - | - | - | - |
| 2 | 339.38 | 302.22 | 331.50 | 333.52 | 293.56 | 312.40 | 293.56 | **87.78** | 54460.77 | 54546.90 | 10769.26 | 33321.80 |
| 3 | **1109.01** | 1168.51 | 1165.22 | 1144.74 | 1153.25 | 2305.14 | 1153.25 | 1144.90 | 24259.50 | 24223.92 | 2651.54 | 5787.03 |
| 4 | 10.38 | 11.66 | 11.16 | 10.98 | 11.53 | 10.05 | 11.53 | 9.97 | 9.54 | 9.53 | 428.60 | **7.59** |
| Avg. | **514.21** | 540.27 | 539.54 | 530.34 | 533.06 | 1049.31 | 533.06 | 521.41 | 12757.84 | 12744.86 | 1781.66 | 3747.14 |
| **3-Week Forecasting Horizon** | | | | | | | | | | | | |
| 1 | - | - | - | - | - | - | - | - | - | - | - | - |
| 2 | 4940.93 | 4945.14 | 4951.95 | 5313.47 | 4867.43 | **3047.08** | 4867.43 | 4455.09 | 67790.53 | 67891.15 | 10111.25 | 41382.55 |
| 3 | 926.27 | 972.10 | 962.62 | 963.30 | 961.54 | 2807.72 | 961.54 | **900.27** | 32801.28 | 32756.34 | 3473.86 | 9453.90 |
| 4 | 12.66 | 14.30 | 13.63 | 13.37 | 13.73 | 10.48 | 13.73 | 11.35 | 9.87 | 9.86 | 794.98 | **8.15** |
| Avg. | 592.15 | 613.69 | 609.33 | 621.97 | 605.98 | 1369.13 | 605.98 | **563.06** | 17046.73 | 17030.05 | 2317.11 | 5669.15 |
| **4-Week Forecasting Horizon** | | | | | | | | | | | | |
| 1 | - | - | - | - | - | - | - | - | - | - | - | - |
| 2 | 4003.09 | 4006.74 | 3924.69 | 4029.80 | 3953.65 | **2379.75** | 3953.65 | 3187.05 | 112941.27 | 113072.58 | 20494.91 | 75220.95 |
| 3 | **852.78** | 896.75 | 892.45 | 892.37 | 896.77 | 3000.85 | 896.77 | 866.86 | 41127.44 | 41072.35 | 4309.56 | 12971.18 |
| 4 | 15.28 | 17.66 | 16.22 | 17.10 | 16.59 | 10.64 | 16.59 | 11.89 | 10.19 | 10.17 | 1265.29 | **8.25** |
| Avg. | 528.22 | 549.29 | 543.79 | 547.83 | 546.92 | 1432.77 | 546.92 | **504.64** | 22336.23 | 22316.06 | 3293.05 | 8412.76 |

[1] Data was obtained from the Our World in Data Team (OWID) [1].
[2] The time-period of the epidemic, where *1* refers to ascending phases (June 2022), *2* the peak (Late June – Mid July 2022), *3* for the immediate declining phase (Late July – Mid October 2022), and *4* for the tail-end of the epidemic (Late-October 2022 - Current).
[3] The models of interest include models of the spatial-wave framework (SW), and *n*-sub-epidemic frameworks (SE), a simple linear regression model, Facebook's Prophet model (Prophet), a general additive model, and an auto-regressive integrated moving average model (ARIMA). All models were calibrated with 11-weeks' worth of data at a time.
[4] Metrics are not available for the ascending phase of the epidemic in Germany, as it occurred in June 2022. Therefore, not enough observed weeks were available to conduct forecasts for the ascending phase of the epidemic.
* The bolded values indicate the best performing model(s) at each time-period for each forecasting horizon. Here, *best performing* refers to the model with the lowest metric value.

**Table D2.** Average Mean Absolute Error (MAE) of the forecasts generated for Germany [1] (weeks of July 14th, 2022, through February 23rd, 2023) for each forecasting horizon (1-4 weeks), and time-period [2] of the epidemic, and averaged across all forecast periods (*Avg.*) for each model [3] of interest.

| Time Period | SW 1st Ranked | SW 2nd Ranked | SW Weighted Ensemble (2) | SW Unweighted Ensemble (2) | SE 1st Ranked | SE 2nd Ranked | SE Weighted Ensemble (2) | SE Unweighted Ensemble (2) | Simple Linear Regression | Prophet | General Additive Model | ARIMA[3] |
|---|---|---|---|---|---|---|---|---|---|---|---|---|
| **1-Week Forecasting Horizon** | | | | | | | | | | | | |
| 1 | - | - | - | - | - | - | - | - | - | - | - | - |
| 2 | 22.36 | 21.82 | 22.18 | 21.74 | 20.85 | 15.52 | 20.85 | **11.42** | 155.20 | 155.38 | 56.83 | 108.60 |
| 3 | **21.16** | 21.21 | 21.21 | 21.47 | 22.05 | 34.41 | 22.05 | 24.43 | 96.48 | 96.43 | 40.43 | 51.12 |
| 4 | 2.06 | 2.15 | 2.15 | 2.15 | 2.23 | 2.17 | 2.23 | 2.08 | 2.19 | 2.18 | 5.36 | **1.99** |
| Avg. | **11.32** | 11.37 | 11.39 | 11.49 | 11.76 | 17.08 | 11.76 | 12.42 | 49.73 | 49.72 | 22.86 | 27.69 |
| **2-Week Forecasting Horizon** | | | | | | | | | | | | |
| 1 | - | - | - | - | - | - | - | - | - | - | - | - |
| 2 | 18.05 | 16.94 | 17.84 | 17.89 | 16.64 | 17.62 | 16.64 | **8.61** | 223.23 | 223.42 | 96.07 | 171.40 |
| 3 | **20.65** | 21.31 | 21.02 | 20.89 | 21.36 | 29.69 | 21.36 | 21.85 | 107.92 | 107.86 | 36.19 | 52.36 |
| 4 | 2.22 | 2.33 | 2.31 | 2.29 | 2.39 | 2.35 | 2.39 | 2.24 | 2.20 | 2.20 | 7.71 | **2.02** |
| Avg. | **11.03** | 11.35 | 11.23 | 11.16 | 11.38 | 15.13 | 11.38 | 11.25 | 57.22 | 57.19 | 23.52 | 30.43 |
| **3-Week Forecasting Horizon** | | | | | | | | | | | | |
| 1 | - | - | - | - | - | - | - | - | - | - | - | - |
| 2 | 51.57 | 50.66 | 51.75 | 55.20 | 50.61 | **42.54** | 50.61 | 44.72 | 251.26 | 251.46 | 95.31 | 194.20 |
| 3 | **18.83** | 19.32 | 19.07 | 19.07 | 19.53 | 32.76 | 19.53 | 19.75 | 120.40 | 120.33 | 39.55 | 62.59 |
| 4 | 2.36 | 2.50 | 2.48 | 2.43 | 2.49 | 2.34 | 2.49 | 2.34 | 2.23 | 2.23 | 9.87 | **2.13** |
| Avg. | **11.44** | 11.70 | 11.62 | 11.71 | 11.78 | 17.36 | 11.78 | 11.61 | 63.79 | 63.76 | 26.12 | 35.86 |
| **4-Week Forecasting Horizon** | | | | | | | | | | | | |
| 1 | - | - | - | - | - | - | - | - | - | - | - | - |
| 2 | 47.55 | 47.08 | 46.28 | 47.61 | 46.61 | **36.26** | 46.61 | 37.79 | 313.05 | 313.25 | 128.30 | 250.75 |
| 3 | **18.50** | 19.02 | 18.87 | 18.82 | 19.17 | 33.66 | 19.17 | 19.56 | 130.41 | 130.34 | 40.51 | 69.90 |
| 4 | 2.49 | 2.63 | 2.56 | 2.59 | 2.58 | 2.24 | 2.58 | 2.34 | 2.24 | 2.24 | 11.76 | **2.11** |
| Avg. | **11.22** | 11.51 | 11.38 | 11.42 | 11.54 | 17.50 | 11.54 | 11.28 | 70.41 | 70.39 | 28.66 | 41.07 |

[1] Data was obtained from the Our World in Data Team (OWID) [1].
[2] The time-period of the epidemic, where *1* refers to ascending phases (June 2022), *2* the peak (Late June – Mid July 2022), *3* for the immediate declining phase (Late July – Mid October 2022), and *4* for the tail-end of the epidemic (Late-October 2022 - Current).
[3] The models of interest include models of the spatial-wave framework (SW), and *n*-sub-epidemic frameworks (SE), a simple linear regression model, Facebook's Prophet model (Prophet), a general additive model, and an auto-regressive integrated moving average model (ARIMA). All models were calibrated with 11-weeks' worth of data at a time.
[4] Metrics are not available for the ascending phase of the epidemic in Germany, as it occurred in June 2022. Therefore, not enough observed weeks were available to conduct forecasts for the ascending phase of the epidemic.
* The bolded values indicate the best performing model(s) at each time-period for each forecasting horizon. Here, *best performing* refers to the model with the lowest metric value.

**Table D3.** Average 95% Prediction Interval Coverage (95% PI Coverage) of the forecasts generated for Germany [1] (weeks of July 14th, 2022, through February 23rd, 2023) for each forecasting horizon (1-4 weeks), and time-period [2] of the epidemic, and averaged across all forecast periods (*Avg.*) for each model [3] of interest.

| Time Period | SW 1st Ranked | SW 2nd Ranked | SW Weighted Ensemble (2) | SW Unweighted Ensemble (2) | SE 1st Ranked | SE 2nd Ranked | SE Weighted Ensemble (2) | SE Unweighted Ensemble (2) | Simple Linear Regression | Prophet | General Additive Model | ARIMA[3] |
|---|---|---|---|---|---|---|---|---|---|---|---|---|
| **1-Week Forecasting Horizon** | | | | | | | | | | | | |
| 1 | - | - | - | - | - | - | - | - | - | - | - | - |
| 2 | **100.00** | **100.00** | **100.00** | **100.00** | **100.00** | **100.00** | **100.00** | **100.00** | 0.00 | 0.00 | **100.00** | 0.00 |
| 3 | **92.31** | **92.31** | **92.31** | **92.31** | **92.31** | **92.31** | **92.31** | **92.31** | 15.38 | 53.85 | 69.23 | 76.92 |
| 4 | **86.67** | **86.67** | **86.67** | **86.67** | **86.67** | **86.67** | **86.67** | **86.67** | 53.33 | 73.33 | 73.33 | **86.67** |
| Avg. | **89.66** | **89.66** | **89.66** | **89.66** | **89.66** | **89.66** | **89.66** | **89.66** | 34.48 | 62.07 | 72.41 | 79.31 |
| **2-Week Forecasting Horizon** | | | | | | | | | | | | |
| 1 | - | - | - | - | - | - | - | - | - | - | - | - |
| 2 | **100.00** | **100.00** | **100.00** | **100.00** | **100.00** | **100.00** | **100.00** | **100.00** | 0.00 | 0.00 | 50.00 | 0.00 |
| 3 | 92.31 | 92.31 | 92.31 | 92.31 | 92.31 | **96.15** | 92.31 | **96.15** | 15.38 | 34.62 | 76.92 | 88.46 |
| 4 | 83.33 | 83.33 | 83.33 | **86.67** | 83.33 | 80.00 | 83.33 | 83.33 | 46.67 | 56.67 | 73.33 | **86.67** |
| Avg. | 87.93 | 87.93 | 87.93 | **89.66** | 87.93 | 87.93 | 87.93 | **89.66** | 31.03 | 44.83 | 74.14 | 84.48 |
| **3-Week Forecasting Horizon** | | | | | | | | | | | | |
| 1 | - | - | - | - | - | - | - | - | - | - | - | - |
| 2 | 66.67 | 66.67 | 66.67 | 66.67 | **100.00** | **100.00** | **100.00** | **100.00** | 0.00 | 0.00 | 66.67 | 0.00 |
| 3 | 92.31 | 92.31 | 92.31 | 92.31 | 92.31 | **97.44** | 92.31 | **97.44** | 15.38 | 30.77 | 79.49 | 87.18 |
| 4 | 82.22 | 82.22 | 82.22 | 82.22 | 82.22 | 80.00 | 82.22 | 82.22 | 44.44 | 57.78 | 73.33 | **88.89** |
| Avg. | 86.21 | 86.21 | 86.21 | 86.21 | 87.36 | 88.51 | 87.36 | **89.66** | 29.89 | 43.68 | 75.86 | 85.06 |
| **4-Week Forecasting Horizon** | | | | | | | | | | | | |
| 1 | - | - | - | - | - | - | - | - | - | - | - | - |
| 2 | 75.00 | 75.00 | 75.00 | 75.00 | **100.00** | **100.00** | **100.00** | **100.00** | 0.00 | 0.00 | 50.00 | 0.00 |
| 3 | **92.31** | **92.31** | **92.31** | **92.31** | **92.31** | 98.08 | **92.31** | 98.08 | 15.38 | 26.92 | 80.77 | 86.54 |
| 4 | 81.67 | 83.33 | 81.67 | 81.67 | 81.67 | 80.00 | 81.67 | 81.67 | 45.00 | 51.67 | 73.33 | **91.67** |
| Avg. | 86.21 | 87.07 | 86.21 | 86.21 | 87.07 | 88.79 | 87.07 | **89.66** | 30.17 | 38.79 | 75.86 | 86.21 |

[1] Data was obtained from the Our World in Data Team (OWID) [1].
[2] The time-period of the epidemic, where *1* refers to ascending phases (June 2022), *2* the peak (Late June – Mid July 2022), *3* for the immediate declining phase (Late July – Mid October 2022), and *4* for the tail-end of the epidemic (Late-October 2022 - Current).
[3] The models of interest include models of the spatial-wave framework (SW), and *n*-sub-epidemic frameworks (SE), a simple linear regression model, Facebook's Prophet model (Prophet), a general additive model, and an auto-regressive integrated moving average model (ARIMA). All models were calibrated with 11-weeks' worth of data at a time.
[4] Metrics are not available for the ascending phase of the epidemic in Germany, as it occurred in June 2022. Therefore, not enough observed weeks were available to conduct forecasts for the ascending phase of the epidemic.
* The bolded values indicate the best performing model(s) at each time-period for each forecasting horizon. Here, *best performing* corresponds to the model which produces coverage closest to 95%.
** Each value is shown as a percentage (%).

**Table D4.** Average Weighted Interval Scores (WIS) of the forecasts generated for Germany[1] (weeks of July 14th, 2022, through February 23rd, 2023) for each forecasting horizon (1-4 weeks), and time-period [2] of the epidemic, and averaged across all forecast periods (*Avg.*) for each model [3] of interest.

| Time Period | SW 1st Ranked | SW 2nd Ranked | SW Weighted Ensemble (2) | SW Unweighted Ensemble (2) | SE 1st Ranked | SE 2nd Ranked | SE Weighted Ensemble (2) | SE Unweighted Ensemble (2) | Simple Linear Regression | Prophet | General Additive Model | ARIMA[3] |
|---|---|---|---|---|---|---|---|---|---|---|---|---|
| **1-Week Forecasting Horizon** | | | | | | | | | | | | |
| 1 | - | - | - | - | - | - | - | - | - | - | - | - |
| 2 | 12.08 | 12.42 | 12.36 | 12.14 | 11.78 | 18.78 | **11.67** | 12.63 | 128.78 | 118.48 | 33.81 | 73.39 |
| 3 | **15.90** | 16.42 | 16.08 | 16.11 | 16.09 | 26.70 | 16.09 | 19.33 | 74.20 | 67.74 | 27.15 | 33.49 |
| 4 | **1.39** | 1.42 | 1.39 | 1.40 | 1.44 | 1.50 | 1.43 | 1.40 | 1.84 | 1.64 | 3.88 | 1.65 |
| Avg. | **8.27** | 8.53 | 8.35 | 8.37 | 8.37 | 13.40 | 8.36 | 9.82 | 38.66 | 35.30 | 15.34 | 18.39 |
| **2-Week Forecasting Horizon** | | | | | | | | | | | | |
| 1 | - | - | - | - | - | - | - | - | - | - | - | - |
| 2 | 11.33 | 11.62 | 11.41 | 11.58 | **10.74** | 22.39 | 10.76 | 13.92 | 195.06 | 187.22 | 62.89 | 128.83 |
| 3 | **14.86** | 15.28 | 15.07 | 15.07 | 15.23 | 23.61 | 15.21 | 16.95 | 84.52 | 79.01 | 24.40 | 33.09 |
| 4 | 1.61 | 1.62 | 1.61 | **1.61** | 1.66 | 1.76 | 1.66 | 1.63 | 1.88 | 1.75 | 5.66 | 1.82 |
| Avg. | **7.88** | 8.09 | 7.98 | 7.99 | 8.06 | 12.27 | 8.05 | 8.92 | 45.59 | 42.78 | 16.04 | 20.21 |
| **3-Week Forecasting Horizon** | | | | | | | | | | | | |
| 1 | - | - | - | - | - | - | - | - | - | - | - | - |
| 2 | 34.35 | 34.50 | 34.03 | 34.87 | 33.08 | 30.71 | 32.96 | **29.09** | 221.31 | 214.26 | 59.04 | 145.43 |
| 3 | **13.68** | 14.12 | 13.90 | 13.88 | 13.96 | 24.92 | 13.98 | 16.63 | 95.90 | 91.31 | 25.28 | 38.68 |
| 4 | 1.75 | 1.75 | 1.75 | 1.75 | 1.80 | 1.86 | 1.80 | **1.73** | 1.91 | 1.80 | 7.29 | 2.03 |
| Avg. | **8.22** | 8.43 | 8.31 | 8.33 | 8.33 | 13.19 | 8.33 | 9.35 | 51.61 | 49.25 | 17.14 | 23.40 |
| **4-Week Forecasting Horizon** | | | | | | | | | | | | |
| 1 | - | - | - | - | - | - | - | - | - | - | - | - |
| 2 | 30.29 | 30.44 | 30.09 | 30.42 | 29.03 | 29.70 | 29.00 | **26.08** | 281.29 | 276.18 | 81.88 | 196.52 |
| 3 | **13.19** | 13.63 | 13.47 | 13.39 | 13.59 | 25.04 | 13.55 | 16.30 | 105.10 | 101.86 | 26.07 | 42.89 |
| 4 | 1.86 | 1.87 | 1.86 | 1.85 | 1.92 | 1.91 | 1.91 | **1.80** | 1.93 | 1.85 | 8.76 | 2.20 |
| Avg. | **7.92** | 8.13 | 8.04 | 8.01 | 8.08 | 13.24 | 8.06 | 9.14 | 57.81 | 56.14 | 19.04 | 27.14 |

[1] Data was obtained from the Our World in Data Team (OWID) [1].
[2] The time-period of the epidemic, where *1* refers to ascending phases (June 2022), *2* the peak (Late June – Mid July 2022), *3* for the immediate declining phase (Late July – Mid October 2022), and *4* for the tail-end of the epidemic (Late-October 2022 - Current).
[3] The models of interest include models of the spatial-wave framework (SW), and *n*-sub-epidemic frameworks (SE), a simple linear regression model, Facebook's Prophet model (Prophet), a general additive model, and an auto-regressive integrated moving average model (ARIMA). All models were calibrated with 11-weeks' worth of data at a time.
[4] Metrics are not available for the ascending phase of the epidemic in Germany, as it occurred in June 2022. Therefore, not enough observed weeks were available to conduct forecasts for the ascending phase of the epidemic.
* The bolded values indicate the best performing model(s) at each time-period for each forecasting horizon. Here, *best performing* refers to the model with the lowest metric value.

**Table D5.** Skills scores[1] for overall average forecast performance metrics (weeks of July 14th, 2022, through February 23rd, 2023) comparing the ARIMA[2] model to the *n*-sub-epidemic (SE) and spatial-wave (SW) frameworks across forecasting horizons (1-4 weeks) for Germany.[3]

| **Baseline[4]** | ARIMA | | | | | | | |
|---|---|---|---|---|---|---|---|---|
| **Comparison[5]** | SW 1st Ranked | SW 2nd Ranked | SW Weighted Ensemble (2) | SW Unweighted Ensemble (2) | SE 1st Ranked | SE 2nd Ranked | SE Weighted Ensemble (2) | SE Unweighted Ensemble (2) |
| **Mean Squared Error (MSE)** | | | | | | | | |
| 1-Week | 79.96 | 79.00 | 78.93 | 78.91 | 79.70 | 47.25 | 79.70 | 77.49 |
| 2-Week | 86.28 | 85.58 | 85.60 | 85.85 | 85.77 | 72.00 | 85.77 | 86.09 |
| 3-Week | 89.55 | 89.17 | 89.25 | 89.03 | 89.31 | 75.85 | 89.31 | 90.07 |
| 4-Week | 93.72 | 93.47 | 93.54 | 93.49 | 93.50 | 82.97 | 93.50 | 94.00 |
| **Mean Absolute Error (MAE)** | | | | | | | | |
| 1-Week | 59.13 | 58.94 | 58.88 | 58.52 | 57.54 | 38.32 | 57.54 | 55.15 |
| 2-Week | 63.76 | 62.71 | 63.08 | 63.31 | 62.59 | 50.26 | 62.59 | 63.02 |
| 3-Week | 68.09 | 67.37 | 67.60 | 67.34 | 67.13 | 51.58 | 67.13 | 67.62 |
| 4-Week | 72.69 | 71.97 | 72.29 | 72.20 | 71.91 | 57.39 | 71.91 | 72.54 |
| **95% Prediction Interval Coverage (95% PI) *** | | | | | | | | |
| 1-Week | 41.18 | 33.24 | 36.41 | 38.76 | 39.09 | 41.13 | 39.82 | 42.79 |
| 2-Week | 59.53 | 56.58 | 58.09 | 58.28 | 58.08 | 52.40 | 57.65 | 56.23 |
| 3-Week | 70.54 | 67.93 | 69.26 | 69.33 | 70.79 | 60.24 | 71.08 | 66.38 |
| 4-Week | 79.78 | 77.57 | 78.87 | 78.69 | 79.35 | 66.85 | 79.74 | 74.36 |
| **Weighted Interval Score (WIS)** | | | | | | | | |
| 1-Week | 55.06 | 53.65 | 54.59 | 54.50 | 54.52 | 27.17 | 54.57 | 46.60 |
| 2-Week | 61.00 | 59.99 | 60.50 | 60.50 | 60.15 | 39.31 | 60.18 | 55.87 |
| 3-Week | 64.88 | 64.00 | 64.50 | 64.41 | 64.40 | 43.65 | 64.39 | 60.05 |
| 4-Week | 70.82 | 70.05 | 70.39 | 70.48 | 70.21 | 51.22 | 70.30 | 66.34 |

[1] Skill scores are calculated by subtracting the mean metric score for the comparison model from the benchmark model, dividing by the mean metric score for the benchmark model and multiplying by 100. They are shown here as a percentage.
[2] Auto-regressive integrated moving average
[3] Data was obtained from the Our World in Data Team (OWID) [1].
[4] The ARIMA model used an 11-week calibration period for all forecasts.
[5] Comparison models used only a 11-week calibration period and were comprised of the 1st ranked, 2nd ranked, weighted (W) ensemble, and unweighted (UW) ensemble models from both the *n*-sub-epidemic (SE) and spatial-wave (SW) frameworks.
* Skill scores for 95% PI coverage are based on average Winkler scores.

**Table E1.** Average Mean Squared Error (MSE) of the forecasts generated for Spain [1] (weeks of July 14th, 2022, through February 23rd, 2023) for each forecasting horizon (1-4 weeks), and time-period [2] of the epidemic, and averaged across all forecast periods (*Avg.*) for each model [3] of interest.

| Time Period | SW 1st Ranked | SW 2nd Ranked | SW Weighted Ensemble (2) | SW Unweighted Ensemble (2) | SE 1st Ranked | SE 2nd Ranked | SE Weighted Ensemble (2) | SE Unweighted Ensemble (2) | Simple Linear Regression | Prophet | General Additive Model | ARIMA[3] |
|---|---|---|---|---|---|---|---|---|---|---|---|---|
| **1-Week Forecasting Horizon** | | | | | | | | | | | | |
| 1 | - | - | - | - | - | - | - | - | - | - | - | - |
| 2 | 49654.56 | 50504.67 | 49593.05 | 49297.28 | 46795.07 | 158507.45 | 46795.07 | 67217.28 | 91586.05 | 91289.07 | 134716.77 | **28177.20** |
| 3 | 4117.70 | 2354.15 | 2994.76 | 3354.45 | **2088.75** | 7768.47 | **2088.75** | 4215.93 | 58788.40 | 58619.31 | 8473.50 | 24449.59 |
| 4 | 696.21 | 687.89 | 690.28 | 691.68 | **681.38** | 1993.84 | **681.38** | 725.98 | 802.86 | 802.79 | 3533.47 | 982.00 |
| Avg. | 10081.16 | 9736.65 | 9757.51 | 9806.52 | **9020.26** | 30571.94 | **9020.26** | 13152.74 | 32451.14 | 32353.26 | 27514.05 | 12144.64 |
| **2-Week Forecasting Horizon** | | | | | | | | | | | | |
| 1 | - | - | - | - | - | - | - | - | - | - | - | - |
| 2 | 60784.42 | 60822.60 | 61089.17 | 60460.76 | 56722.71 | 285245.79 | 56722.71 | 89432.30 | 157447.57 | 157065.16 | 180306.82 | **35756.20** |
| 3 | 3972.29 | 2257.23 | 3030.02 | 2861.60 | **1961.70** | 6907.96 | **1961.70** | 3325.91 | 66931.42 | 66765.83 | 9518.72 | 38733.21 |
| 4 | 657.30 | **650.05** | 655.07 | 654.69 | 659.99 | 1889.97 | 659.99 | 713.35 | 744.20 | 744.12 | 7015.43 | 952.58 |
| Avg. | 11938.52 | 11467.99 | 11729.90 | 11574.88 | **10685.07** | 52128.70 | **10685.07** | 16730.43 | 46020.57 | 45908.91 | 37583.82 | 17375.45 |
| **3-Week Forecasting Horizon** | | | | | | | | | | | | |
| 1 | - | - | - | - | - | - | - | - | - | - | - | - |
| 2 | 66238.66 | 65853.27 | 65830.40 | 65411.27 | 61798.72 | 712036.46 | 61798.72 | 91258.05 | 249593.80 | 249070.06 | 209693.49 | **51530.73** |
| 3 | 3549.08 | 2072.15 | 2695.07 | 2711.81 | **1910.32** | 6021.70 | **1910.32** | 3090.46 | 75095.49 | 74919.98 | 8182.10 | 48521.64 |
| 4 | 636.57 | **630.42** | 633.29 | 631.97 | 639.26 | 2594.06 | 639.26 | 643.52 | 721.13 | 721.03 | 10842.05 | 865.58 |
| Avg. | 12750.72 | 12273.46 | 12442.94 | 12374.56 | **11534.63** | 125857.27 | **11534.63** | 16941.73 | 64147.28 | 64008.51 | 44393.00 | 22747.45 |
| **4-Week Forecasting Horizon** | | | | | | | | | | | | |
| 1 | - | - | - | - | - | - | - | - | - | - | - | - |
| 2 | 69981.69 | 69480.75 | 68531.46 | 68528.69 | **66213.09** | 2274638.34 | **66213.09** | 98574.54 | 354349.15 | 353674.23 | 253973.74 | 72828.55 |
| 3 | 3375.59 | 2011.58 | 2629.29 | 2436.91 | **1895.99** | 5881.04 | **1895.99** | 2891.31 | 85738.74 | 85544.70 | 6873.62 | 57354.17 |
| 4 | 631.33 | **625.79** | 631.67 | 631.35 | 628.89 | 3858.51 | 628.89 | 629.30 | 703.50 | 703.41 | 15040.60 | 845.57 |
| Avg. | 13345.32 | 12879.62 | 12889.60 | 12835.87 | **12286.05** | 395930.21 | **12286.05** | 18140.42 | 85134.88 | 84964.94 | 53983.01 | 28845.01 |

[1] Data was obtained from the Our World in Data Team (OWID) [1].
[2] The time-period of the epidemic, where *1* refers to ascending phases (June 2022), *2* the peak (Early July – Mid-August 2022), *3* for the immediate declining phase (Mid-August – Ealy October 2022), and *4* for the tail-end of the epidemic (Mid-October 2022 - Current).
[3] The models of interest include models of the spatial-wave framework (SW), and *n*-sub-epidemic frameworks (SE), a simple linear regression model, Facebook's Prophet model (Prophet), a general additive model, and an auto-regressive integrated moving average model (ARIMA). All models were calibrated with 11-weeks' worth of data at a time.
[4] Metrics are not available for the ascending phase of the epidemic in Spain, as it occurred in June 2022. Therefore, not enough observed weeks were available to conduct forecasts for the ascending phase of the epidemic.
* The bolded values indicate the best performing model(s) at each time-period for each forecasting horizon. Here, *best performing* refers to the model with the lowest metric value.

**Table E2.** Average Mean Absolute Error (MAE) of the forecasts generated for Spain [1] (weeks of July 14th, 2022, through February 23rd, 2023) for each forecasting horizon (1-4 weeks), and time-period [2] of the epidemic, and averaged across all forecast periods (*Avg.*) for each model [3] of interest.

| Time Period | SW 1st Ranked | SW 2nd Ranked | SW Weighted Ensemble (2) | SW Unweighted Ensemble (2) | SE 1st Ranked | SE 2nd Ranked | SE Weighted Ensemble (2) | SE Unweighted Ensemble (2) | Simple Linear Regression | Prophet | General Additive Model | ARIMA[3] |
|---|---|---|---|---|---|---|---|---|---|---|---|---|
| **1-Week Forecasting Horizon** | | | | | | | | | | | | |
| 1 | - | - | - | - | - | - | - | - | - | - | - | - |
| 2 | 153.38 | 155.14 | 152.74 | 148.59 | 152.86 | 375.57 | 152.86 | 221.83 | 267.19 | 266.72 | 308.33 | **145.60** |
| 3 | 47.62 | 39.59 | 41.47 | 43.81 | **39.33** | 62.73 | **39.33** | 48.63 | 159.14 | 158.50 | 65.11 | 111.84 |
| 4 | 16.90 | **16.44** | 16.60 | 16.64 | 16.57 | 27.46 | 16.57 | 16.62 | 18.02 | 18.03 | 39.14 | 24.35 |
| Avg. | 48.90 | 46.74 | 46.93 | 46.89 | **46.35** | 97.21 | **46.35** | 60.83 | 99.91 | 99.66 | 92.72 | 69.39 |
| **2-Week Forecasting Horizon** | | | | | | | | | | | | |
| 1 | - | - | - | - | - | - | - | - | - | - | - | - |
| 2 | 198.30 | 199.60 | 198.34 | 197.79 | 193.47 | 458.52 | 193.47 | 241.99 | 343.85 | 343.34 | 349.23 | **159.00** |
| 3 | 44.93 | 37.40 | 40.51 | 40.04 | **35.95** | 57.66 | **35.95** | 42.28 | 168.86 | 168.47 | 76.12 | 142.99 |
| 4 | 15.72 | **15.44** | 15.60 | 15.52 | 15.81 | 26.48 | 15.81 | 16.33 | 16.54 | 16.54 | 47.72 | 22.95 |
| Avg. | 55.25 | 53.25 | 53.98 | 53.71 | **52.00** | 109.57 | **52.00** | 62.39 | 114.99 | 114.80 | 107.54 | 79.52 |
| **3-Week Forecasting Horizon** | | | | | | | | | | | | |
| 1 | - | - | - | - | - | - | - | - | - | - | - | - |
| 2 | 205.98 | 207.11 | 205.16 | 204.75 | 200.56 | 587.14 | 200.56 | 250.74 | 439.11 | 438.57 | 390.60 | **186.47** |
| 3 | 43.34 | 35.05 | 38.98 | 38.86 | **34.28** | 55.27 | **34.28** | 41.83 | 175.25 | 174.93 | 71.04 | 158.63 |
| 4 | 15.20 | **14.90** | 14.99 | 15.00 | 15.13 | 27.16 | 15.13 | 15.06 | 15.88 | 15.88 | 53.14 | 21.08 |
| Avg. | 55.86 | 53.60 | 54.40 | 54.30 | **52.38** | 131.46 | **52.38** | 63.08 | 132.81 | 132.63 | 116.26 | 87.54 |
| **4-Week Forecasting Horizon** | | | | | | | | | | | | |
| 1 | - | - | - | - | - | - | - | - | - | - | - | - |
| 2 | 205.72 | 206.70 | 203.98 | 203.97 | **201.13** | 807.97 | **201.13** | 261.73 | 526.72 | 526.14 | 424.12 | 217.85 |
| 3 | 43.79 | 35.47 | 39.50 | 38.31 | **34.47** | 54.84 | **34.47** | 41.05 | 183.01 | 182.74 | 64.51 | 173.22 |
| 4 | 14.75 | 14.48 | 14.67 | 14.70 | 14.68 | 28.60 | 14.68 | **14.45** | 15.39 | 15.39 | 59.22 | 20.73 |
| Avg. | 55.69 | 53.41 | 54.16 | 53.85 | **52.29** | 170.22 | **52.29** | 64.42 | 149.79 | 149.62 | 123.59 | 96.78 |

[1] Data was obtained from the Our World in Data Team (OWID) [1].
[2] The time-period of the epidemic, where *1* refers to ascending phases (June 2022), *2* the peak (Early July – Mid-August 2022), *3* for the immediate declining phase (Mid-August – Ealy October 2022), and *4* for the tail-end of the epidemic (Mid-October 2022 - Current).
[3] The models of interest include models of the spatial-wave framework (SW), and *n*-sub-epidemic frameworks (SE), a simple linear regression model, Facebook's Prophet model (Prophet), a general additive model, and an auto-regressive integrated moving average model (ARIMA). All models were calibrated with 11-weeks' worth of data at a time.
[4] Metrics are not available for the ascending phase of the epidemic in Spain, as it occurred in June 2022. Therefore, not enough observed weeks were available to conduct forecasts for the ascending phase of the epidemic.
* The bolded values indicate the best performing model(s) at each time-period for each forecasting horizon. Here, *best performing* refers to the model with the lowest metric value.

**Table E3.** Average 95% Prediction Interval Coverage (95% PI Coverage) of the forecasts generated for Spain [1] (weeks of July 14th, 2022, through February 23rd, 2023) for each forecasting horizon (1-4 weeks), and time-period [2] of the epidemic, and averaged across all forecast periods (*Avg.*) for each model [3] of interest.

| Time Period | SW 1st Ranked | SW 2nd Ranked | SW Weighted Ensemble (2) | SW Unweighted Ensemble (2) | SE 1st Ranked | SE 2nd Ranked | SE Weighted Ensemble (2) | SE Unweighted Ensemble (2) | Simple Linear Regression | Prophet | General Additive Model | ARIMA[3] |
|---|---|---|---|---|---|---|---|---|---|---|---|---|
| **1-Week Forecasting Horizon** | | | | | | | | | | | | |
| 1 | - | - | - | - | - | - | - | - | - | - | - | - |
| 2 | **100.00** | 80.00 | 80.00 | 80.00 | 80.00 | 80.00 | 80.00 | **100.00** | 40.00 | 60.00 | 60.00 | **100.00** |
| 3 | **100.00** | **100.00** | **100.00** | **100.00** | **100.00** | **100.00** | **100.00** | **100.00** | 62.50 | 75.00 | **100.00** | **100.00** |
| 4 | 87.50 | 87.50 | 87.50 | 87.50 | 87.50 | **93.75** | 87.50 | 87.50 | 75.00 | 81.25 | 68.75 | **93.75** |
| Avg. | 93.10 | 89.66 | 89.66 | 89.66 | 89.66 | 93.10 | 89.66 | 93.10 | 65.52 | 75.86 | 75.86 | **96.55** |
| **2-Week Forecasting Horizon** | | | | | | | | | | | | |
| 1 | - | - | - | - | - | - | - | - | - | - | - | - |
| 2 | **90.00** | **90.00** | **90.00** | **90.00** | **90.00** | 80.00 | **90.00** | **100.00** | 30.00 | 40.00 | 70.00 | **100.00** |
| 3 | **100.00** | **100.00** | **100.00** | **100.00** | **100.00** | **100.00** | **100.00** | **100.00** | 56.25 | 68.75 | **100.00** | **100.00** |
| 4 | 87.50 | 87.50 | 87.50 | 87.50 | 87.50 | 96.88 | 87.50 | **93.75** | 75.00 | 78.13 | 71.88 | **93.75** |
| Avg. | 91.38 | 91.38 | 91.38 | 91.38 | 91.38 | **94.83** | 91.38 | 96.55 | 62.07 | 68.97 | 79.31 | 96.55 |
| **3-Week Forecasting Horizon** | | | | | | | | | | | | |
| 1 | - | - | - | - | - | - | - | - | - | - | - | - |
| 2 | **93.33** | 86.67 | **93.33** | 86.67 | 86.67 | 80.00 | 86.67 | 100.00 | 20.00 | 26.67 | 73.33 | 100.00 |
| 3 | **100.00** | **100.00** | **100.00** | **100.00** | **100.00** | **100.00** | **100.00** | **100.00** | 54.17 | 62.50 | **100.00** | **100.00** |
| 4 | 89.58 | 89.58 | 89.58 | 89.58 | 89.58 | **95.83** | 89.58 | 93.75 | 72.92 | 75.00 | 75.00 | **95.83** |
| Avg. | 93.10 | 91.95 | 93.10 | 91.95 | 91.95 | **94.25** | 91.95 | 96.55 | 58.62 | 63.22 | 81.61 | 97.70 |
| **4-Week Forecasting Horizon** | | | | | | | | | | | | |
| 1 | - | - | - | - | - | - | - | - | - | - | - | - |
| 2 | **100.00** | 85.00 | 85.00 | 85.00 | 85.00 | 80.00 | 85.00 | **100.00** | 15.00 | 20.00 | 75.00 | **100.00** |
| 3 | 100.00 | 100.00 | 100.00 | 100.00 | 100.00 | 100.00 | 100.00 | 100.00 | 50.00 | 59.38 | 100.00 | **96.88** |
| 4 | 90.63 | 90.63 | 90.63 | 90.63 | 90.63 | 98.44 | 90.63 | **95.31** | 71.88 | 73.44 | 76.56 | 96.88 |
| Avg. | **94.83** | 92.24 | 92.24 | 92.24 | 92.24 | 95.69 | 92.24 | 97.41 | 56.03 | 60.34 | 82.76 | 97.41 |

[1] Data was obtained from the Our World in Data Team (OWID) [1].
[2] The time-period of the epidemic, where *1* refers to ascending phases (June 2022), *2* the peak (Early July – Mid-August 2022), *3* for the immediate declining phase (Mid-August – Ealy October 2022), and *4* for the tail-end of the epidemic (Mid-October 2022 - Current).
[3] The models of interest include models of the spatial-wave framework (SW), and *n*-sub-epidemic frameworks (SE), a simple linear regression model, Facebook's Prophet model (Prophet), a general additive model, and an auto-regressive integrated moving average model (ARIMA). All models were calibrated with 11-weeks' worth of data at a time.
[4] Metrics are not available for the ascending phase of the epidemic in Spain, as it occurred in June 2022. Therefore, not enough observed weeks were available to conduct forecasts for the ascending phase of the epidemic.
* The bolded values indicate the best performing model(s) at each time-period for each forecasting horizon. Here, *best performing* corresponds to the model which produces coverage closest to 95%.
** Each value is shown as a percentage (%).

**Table E4.** Average Weighted Interval Scores (WIS) of the forecasts generated for Spain[1] (weeks of July 14th, 2022, through February 23rd, 2023) for each forecasting horizon (1-4 weeks), and time-period [2] of the epidemic, and averaged across all forecast periods (*Avg.*) for each model [3] of interest.

| Time Period | SW 1st Ranked | SW 2nd Ranked | SW Weighted Ensemble (2) | SW Unweighted Ensemble (2) | SE 1st Ranked | SE 2nd Ranked | SE Weighted Ensemble (2) | SE Unweighted Ensemble (2) | Simple Linear Regression | Prophet | General Additive Model | ARIMA[3] |
|---|---|---|---|---|---|---|---|---|---|---|---|---|
| **1-Week Forecasting Horizon** | | | | | | | | | | | | |
| 1 | - | - | - | - | - | - | - | - | - | - | - | - |
| 2 | 107.61 | 113.19 | 109.55 | 109.86 | 110.17 | 246.71 | 110.31 | 143.42 | 195.04 | 176.10 | 230.40 | **87.84** |
| 3 | 32.70 | **29.46** | 30.60 | 30.96 | 29.89 | 48.45 | 29.61 | 37.55 | 112.80 | 102.36 | 42.19 | 68.61 |
| 4 | 12.05 | 11.94 | 11.98 | 12.01 | **11.74** | 16.58 | 11.77 | 12.82 | 15.21 | 14.42 | 29.41 | 14.45 |
| Avg. | 34.22 | 34.23 | 33.94 | 34.11 | 33.72 | 65.05 | **33.68** | 42.16 | 73.14 | 66.56 | 67.59 | 42.05 |
| **2-Week Forecasting Horizon** | | | | | | | | | | | | |
| 1 | - | - | - | - | - | - | - | - | - | - | - | - |
| 2 | 119.57 | 124.63 | 122.77 | 121.99 | 118.49 | 312.70 | 118.36 | 169.19 | 263.63 | 243.55 | 264.91 | **97.94** |
| 3 | 32.07 | **28.95** | 30.70 | 30.14 | 29.01 | 47.75 | 29.03 | 36.70 | 120.50 | 112.08 | 43.53 | 87.47 |
| 4 | 11.56 | 11.48 | 11.54 | 11.50 | 11.40 | 16.19 | **11.38** | 12.26 | 14.11 | 13.57 | 36.84 | 14.38 |
| Avg. | 35.84 | 35.81 | 36.00 | 35.69 | 34.72 | 76.02 | **34.69** | 46.06 | 86.48 | 80.39 | 78.01 | 48.95 |
| **3-Week Forecasting Horizon** | | | | | | | | | | | | |
| 1 | - | - | - | - | - | - | - | - | - | - | - | - |
| 2 | 125.30 | 129.50 | 126.42 | 127.70 | 122.67 | 399.48 | 122.54 | 198.01 | 347.44 | 331.09 | 278.59 | **114.44** |
| 3 | 31.04 | **27.95** | 29.69 | 29.29 | 28.32 | 47.27 | 28.33 | 35.80 | 125.59 | 118.79 | 42.37 | 98.49 |
| 4 | 11.05 | 10.98 | 11.02 | 11.04 | **10.80** | 17.00 | 10.83 | 12.20 | 13.81 | 13.49 | 41.63 | 13.83 |
| Avg. | 36.27 | 36.09 | 36.07 | 36.19 | **34.92** | 91.30 | **34.92** | 50.75 | 102.16 | 97.30 | 82.69 | 54.53 |
| **4-Week Forecasting Horizon** | | | | | | | | | | | | |
| 1 | - | - | - | - | - | - | - | - | - | - | - | - |
| 2 | 127.65 | 131.84 | 128.64 | 129.33 | **126.58** | 571.98 | 126.79 | 247.58 | 426.20 | 413.95 | 294.75 | 132.82 |
| 3 | 30.68 | **27.57** | 29.16 | 28.91 | 27.90 | 47.03 | 27.87 | 35.26 | 131.69 | 126.85 | 41.37 | 108.74 |
| 4 | 10.83 | 10.72 | 10.78 | 10.77 | 10.53 | 18.58 | **10.51** | 12.27 | 13.47 | 13.20 | 45.92 | 13.92 |
| Avg. | 36.45 | 36.26 | 36.17 | 36.21 | **35.33** | 121.84 | 35.35 | 59.19 | 117.24 | 113.65 | 87.57 | 60.57 |

[1] Data was obtained from the Our World in Data Team (OWID) [1].
[2] The time-period of the epidemic, where *1* refers to ascending phases (June 2022), *2* the peak (Early July – Mid-August 2022), *3* for the immediate declining phase (Mid-August – Ealy October 2022), and *4* for the tail-end of the epidemic (Mid-October 2022 - Current).
[3] The models of interest include models of the spatial-wave framework (SW), and *n*-sub-epidemic frameworks (SE), a simple linear regression model, Facebook's Prophet model (Prophet), a general additive model, and an auto-regressive integrated moving average model (ARIMA). All models were calibrated with 11-weeks' worth of data at a time.
[4] Metrics are not available for the ascending phase of the epidemic in Spain, as it occurred in June 2022. Therefore, not enough observed weeks were available to conduct forecasts for the ascending phase of the epidemic.
* The bolded values indicate the best performing model(s) at each time-period for each forecasting horizon. Here, *best performing* refers to the model with the lowest metric value.

**Table E5.** Skills scores[1] for overall average forecast performance metrics (weeks of July 14th, 2022, through February 23rd, 2023) comparing the ARIMA[2] model to the *n*-sub-epidemic (SE) and spatial-wave (SW) frameworks across forecasting horizons (1-4 weeks) for Spain[3].

| **Baseline[4]** | ARIMA | | | | | | | |
|---|---|---|---|---|---|---|---|---|
| **Comparison[5]** | SW 1st Ranked | SW 2nd Ranked | SW Weighted Ensemble (2) | SW Unweighted Ensemble (2) | SE 1st Ranked | SE 2nd Ranked | SE Weighted Ensemble (2) | SE Unweighted Ensemble (2) |
| **Mean Squared Error (MSE)** | | | | | | | | |
| 1-Week | 16.99 | 19.83 | 19.66 | 19.25 | 25.73 | -151.73 | 25.73 | -8.30 |
| 2-Week | 31.29 | 34.00 | 32.49 | 33.38 | 38.50 | -200.01 | 38.50 | 3.71 |
| 3-Week | 43.95 | 46.04 | 45.30 | 45.60 | 49.29 | -453.28 | 49.29 | 25.52 |
| 4-Week | 53.73 | 55.35 | 55.31 | 55.50 | 57.41 | -1272.61 | 57.41 | 37.11 |
| **Mean Absolute Error (MAE)** | | | | | | | | |
| 1-Week | 29.52 | 32.64 | 32.37 | 32.43 | 33.21 | -40.08 | 33.21 | 12.34 |
| 2-Week | 30.51 | 33.03 | 32.12 | 32.46 | 34.61 | -37.79 | 34.61 | 21.54 |
| 3-Week | 36.19 | 38.77 | 37.86 | 37.97 | 40.16 | -50.17 | 40.16 | 27.94 |
| 4-Week | 42.46 | 44.81 | 44.04 | 44.36 | 45.97 | -75.87 | 45.97 | 33.43 |
| **95% Prediction Interval Coverage (95% PI) *** | | | | | | | | |
| 1-Week | 42.92 | 24.48 | 35.97 | 32.89 | 33.61 | -42.49 | 33.22 | 30.82 |
| 2-Week | 46.19 | 41.37 | 44.93 | 42.81 | 45.91 | -66.88 | 44.06 | 30.19 |
| 3-Week | 43.47 | 27.28 | 41.99 | 36.30 | 37.38 | -100.03 | 37.51 | 4.07 |
| 4-Week | 45.16 | 28.01 | 40.70 | 36.32 | 39.55 | -143.19 | 40.12 | -36.72 |
| **Weighted Interval Score (WIS)** | | | | | | | | |
| 1-Week | 18.61 | 18.58 | 19.28 | 18.87 | 19.80 | -54.71 | 19.90 | -0.26 |
| 2-Week | 26.78 | 26.85 | 26.46 | 27.09 | 29.07 | -55.30 | 29.13 | 5.91 |
| 3-Week | 33.50 | 33.81 | 33.86 | 33.63 | 35.96 | -67.42 | 35.97 | 6.94 |
| 4-Week | 39.83 | 40.15 | 40.29 | 40.21 | 41.67 | -101.15 | 41.64 | 2.29 |

[1] Skill scores are calculated by subtracting the mean metric score for the comparison model from the benchmark model, dividing by the mean metric score for the benchmark model and multiplying by 100. They are shown here as a percentage.
[2] Auto-regressive integrated moving average
[3] Data was obtained from the Our World in Data Team (OWID) [1].
[4] The ARIMA model used an 11-week calibration period for all forecasts.
[5] Comparison models used only a 11-week calibration period and were comprised of the 1st ranked, 2nd ranked, weighted (W) ensemble, and unweighted (UW) ensemble models from both the *n*-sub-epidemic (SE) and spatial-wave (SW) frameworks.
* Skill scores for 95% PI coverage are based on average Winkler scores.

**Table F1.** Average Mean Squared Error (MSE) of the forecasts generated for the United Kingdom [1] (weeks of July 14th, 2022, through February 23rd, 2023) for each forecasting horizon (1-4 weeks), and time-period [2] of the epidemic, and averaged across all forecast periods (*Avg.*) for each model [3] of interest.

| Time Period | SW 1st Ranked | SW 2nd Ranked | SW Weighted Ensemble (2) | SW Unweighted Ensemble (2) | SE 1st Ranked | SE 2nd Ranked | SE Weighted Ensemble (2) | SE Unweighted Ensemble (2) | Simple Linear Regression | Prophet | General Additive Model | ARIMA[3] |
|---|---|---|---|---|---|---|---|---|---|---|---|---|
| **1-Week Forecasting Horizon** | | | | | | | | | | | | |
| 1 | - | - | - | - | - | - | - | - | - | - | - | - |
| 2 | 19586.96 | 21377.96 | 20681.78 | 21085.42 | 18309.84 | **5208.93** | 18309.84 | 9705.05 | 22685.84 | 22677.56 | 22685.84 | 21638.41 |
| 3 | 5669.75 | 5685.86 | 5751.66 | 5604.70 | **5475.83** | 96724.31 | **5475.83** | 5956.12 | 15335.65 | 15266.15 | 11918.89 | 10464.61 |
| 4 | **13.35** | 14.23 | 14.15 | 14.36 | 15.95 | 16.23 | 15.95 | 15.71 | 17.67 | 17.67 | 400.15 | 259.75 |
| Avg. | 3223.93 | 3293.37 | 3298.81 | 3246.96 | 3094.30 | 43547.19 | 3094.30 | **3012.77** | 7666.01 | 7634.57 | 6332.19 | 5571.54 |
| **2-Week Forecasting Horizon** | | | | | | | | | | | | |
| 1 | - | - | - | - | - | - | - | - | - | - | - | - |
| 2 | 19746.67 | 21969.93 | 20915.08 | 20680.85 | 17911.77 | **2131.60** | 17911.77 | 5380.22 | 24224.50 | 24215.22 | 24224.50 | 22862.72 |
| 3 | 5389.89 | 5370.16 | 5376.92 | 5316.90 | **5139.11** | 62476.10 | **5139.11** | 5899.05 | 19489.42 | 19429.24 | 10217.01 | 13125.37 |
| 4 | **11.62** | 12.96 | 12.84 | 12.71 | 13.88 | 11.71 | 13.88 | 12.34 | 17.37 | 17.37 | 866.59 | 275.67 |
| Avg. | 3103.09 | 3171.60 | 3138.20 | 3103.15 | 2928.56 | 28086.08 | 2928.56 | **2836.31** | 9580.95 | 9553.65 | 5863.60 | 6814.75 |
| **3-Week Forecasting Horizon** | | | | | | | | | | | | |
| 1 | - | - | - | - | - | - | - | - | - | - | - | - |
| 2 | 32409.55 | 35880.52 | 33399.39 | 33889.19 | 29223.27 | **1636.13** | 29223.27 | 7085.92 | 40702.32 | 40689.50 | 40702.32 | 38526.65 |
| 3 | 5238.49 | 5286.78 | 5206.54 | 5355.93 | 5114.51 | 22393.90 | 5114.51 | **4917.04** | 24707.00 | 24636.40 | 7571.78 | 14839.20 |
| 4 | 12.11 | 13.05 | 12.91 | 13.08 | 13.83 | **11.23** | 13.83 | 12.32 | 17.27 | 17.27 | 1594.69 | 258.30 |
| Avg. | 3472.12 | 3613.95 | 3492.35 | 3576.29 | 3307.56 | 10100.87 | 3307.56 | **2454.91** | 12488.01 | 12455.92 | 5622.61 | 8114.16 |
| **4-Week Forecasting Horizon** | | | | | | | | | | | | |
| 1 | - | - | - | - | - | - | - | - | - | - | - | - |
| 2 | 83774.95 | 90724.08 | 87496.92 | 86861.94 | 77254.25 | **11785.34** | 77254.25 | 20170.12 | 101626.54 | 101605.93 | 101626.54 | 97643.82 |
| 3 | 4420.61 | 4405.54 | 4399.86 | 4427.78 | **4276.69** | 19154.73 | **4276.69** | 5046.89 | 27740.59 | 27655.75 | 8328.83 | 15921.79 |
| 4 | 11.55 | 12.27 | 12.26 | 12.22 | 12.85 | **10.97** | 12.85 | 11.33 | 15.87 | 15.87 | 2606.09 | 247.05 |
| Avg. | 4876.42 | 5109.66 | 4995.83 | 4986.43 | 4587.72 | 8998.67 | 4587.72 | **2963.78** | 15948.01 | 15909.26 | 8585.95 | 10632.17 |

[1] Data was obtained from the Our World in Data Team (OWID) [1].
[2] The time-period of the epidemic, where *1* refers to ascending phases (June – Early July 2022), *2* the peak (Mid-July 2022), *3* for the immediate declining phase (Late July – Mid-October 2022), and *4* for the tail-end of the epidemic (Late-October 2022 - Current).
[3] The models of interest include models of the spatial-wave framework (SW), and *n*-sub-epidemic frameworks (SE), a simple linear regression model, Facebook's Prophet model (Prophet), a general additive model, and an auto-regressive integrated moving average model (ARIMA). All models were calibrated with 11-weeks' worth of data at a time.
[4] Metrics are not available for the ascending phase of the epidemic in the United Kingdom, as it occurred from June to Early July 2022. Therefore, not enough observed weeks were available to conduct forecasts for the ascending phase of the epidemic.
* The bolded values indicate the best performing model(s) at each time-period for each forecasting horizon. Here, *best performing* refers to the model with the lowest metric value.

**Table F2.** Average Mean Absolute Error (MAE) of the forecasts generated for the United Kingdom [1] (weeks of July 14th, 2022, through February 23rd, 2023) for each forecasting horizon (1-4 weeks), and time-period [2] of the epidemic, and averaged across all forecast periods (*Avg.*) for each model [3] of interest.

| Time Period | SW 1st Ranked | SW 2nd Ranked | SW Weighted Ensemble (2) | SW Unweighted Ensemble (2) | SE 1st Ranked | SE 2nd Ranked | SE Weighted Ensemble (2) | SE Unweighted Ensemble (2) | Simple Linear Regression | Prophet | General Additive Model | ARIMA[3] |
|---|---|---|---|---|---|---|---|---|---|---|---|---|
| **1-Week Forecasting Horizon** | | | | | | | | | | | | |
| 1 | - | - | - | - | - | - | - | - | - | - | - | - |
| 2 | 139.95 | 146.21 | 143.81 | 145.21 | 135.31 | **72.17** | 135.31 | 98.51 | 150.62 | 150.59 | 150.62 | 147.10 |
| 3 | 44.39 | 43.40 | 44.14 | 43.30 | **41.43** | 125.96 | **41.43** | 48.59 | 78.92 | 78.81 | 70.53 | 67.93 |
| 4 | 2.72 | 2.78 | 2.75 | 2.80 | 2.89 | 3.10 | 2.89 | 2.97 | **2.48** | 2.49 | 9.46 | 7.39 |
| Avg. | 26.13 | 25.93 | 26.17 | 25.86 | **24.73** | 60.56 | **24.73** | 26.71 | 41.86 | 41.81 | 41.70 | 39.35 |
| **2-Week Forecasting Horizon** | | | | | | | | | | | | |
| 1 | - | - | - | - | - | - | - | - | - | - | - | - |
| 2 | 140.52 | 148.21 | 144.62 | 143.79 | 133.83 | **36.31** | 133.83 | 65.68 | 155.56 | 155.53 | 155.56 | 151.15 |
| 3 | 40.12 | 40.85 | 40.40 | 40.23 | **37.79** | 111.30 | **37.79** | 44.64 | 90.10 | 90.01 | 64.72 | 76.57 |
| 4 | 2.38 | 2.45 | 2.44 | 2.46 | 2.52 | 2.53 | 2.52 | 2.48 | **2.38** | 2.38 | 11.95 | 8.08 |
| Avg. | 24.06 | 24.69 | 24.36 | 24.27 | **22.86** | 52.46 | **22.86** | 23.56 | 46.98 | 46.94 | 40.56 | 43.72 |
| **3-Week Forecasting Horizon** | | | | | | | | | | | | |
| 1 | - | - | - | - | - | - | - | - | - | - | - | - |
| 2 | 173.74 | 182.92 | 176.34 | 177.68 | 165.15 | **31.58** | 165.15 | 81.51 | 194.18 | 194.14 | 194.18 | 188.87 |
| 3 | 39.15 | 39.19 | 38.93 | 39.23 | **37.91** | 66.84 | **37.91** | 38.96 | 99.72 | 99.62 | 55.12 | 83.06 |
| 4 | **2.33** | 2.36 | 2.35 | 2.40 | 2.41 | 2.50 | 2.41 | 2.44 | 2.34 | 2.34 | 14.98 | 8.14 |
| Avg. | 24.75 | 25.09 | 24.75 | 24.95 | 23.94 | 32.34 | 23.94 | **21.54** | 52.61 | 52.56 | 39.15 | 47.96 |
| **4-Week Forecasting Horizon** | | | | | | | | | | | | |
| 1 | - | - | - | - | - | - | - | - | - | - | - | - |
| 2 | 252.22 | 263.52 | 258.46 | 256.83 | 241.40 | **75.34** | 241.40 | 116.83 | 278.95 | 278.92 | 278.95 | 272.75 |
| 3 | 36.47 | 36.06 | 35.90 | 36.11 | **34.95** | 62.38 | **34.95** | 39.44 | 105.22 | 105.10 | 57.00 | 87.06 |
| 4 | 2.17 | 2.17 | 2.19 | 2.22 | 2.21 | 2.41 | 2.21 | **2.17** | 2.17 | 2.17 | 17.95 | 7.98 |
| Avg. | 26.17 | 26.37 | 26.14 | 26.19 | 25.13 | 31.81 | 25.13 | **22.83** | 57.91 | 57.85 | 44.46 | 52.56 |

[1] Data was obtained from the Our World in Data Team (OWID) [1].

[2] The time-period of the epidemic, where *1* refers to ascending phases (June – Early July 2022), *2* the peak (Mid-July 2022), *3* for the immediate declining phase (Late July – Mid-October 2022), and *4* for the tail-end of the epidemic (Late-October 2022 - Current).

[3] The models of interest include models of the spatial-wave framework (SW), and *n*-sub-epidemic frameworks (SE), a simple linear regression model, Facebook's Prophet model (Prophet), a general additive model, and an auto-regressive integrated moving average model (ARIMA). All models were calibrated with 11-weeks' worth of data at a time.

[4] Metrics are not available for the ascending phase of the epidemic in the United Kingdom, as it occurred from June to Early July 2022. Therefore, not enough observed weeks were available to conduct forecasts for the ascending phase of the epidemic.

* The bolded values indicate the best performing model(s) at each time-period for each forecasting horizon. Here, *best performing* refers to the model with the lowest metric value.

**Table F3.** Average 95% Prediction Interval Coverage (95% PI Coverage) of the forecasts generated for the United Kingdom [1] (weeks of July 14th, 2022, through February 23rd, 2023) for each forecasting horizon (1-4 weeks), and time-period [2] of the epidemic, and averaged across all forecast periods (*Avg.*) for each model [3] of interest.

| Time Period | SW 1st Ranked | SW 2nd Ranked | SW Weighted Ensemble (2) | SW Unweighted Ensemble (2) | SE 1st Ranked | SE 2nd Ranked | SE Weighted Ensemble (2) | SE Unweighted Ensemble (2) | Simple Linear Regression | Prophet | General Additive Model | ARIMA[3] |
|---|---|---|---|---|---|---|---|---|---|---|---|---|
| **1-Week Forecasting Horizon** | | | | | | | | | | | | |
| 1 | - | - | - | - | - | - | - | - | - | - | - | - |
| 2 | **0.00** | **0.00** | **0.00** | **0.00** | **0.00** | **0.00** | **0.00** | **0.00** | **0.00** | **0.00** | **0.00** | **0.00** |
| 3 | 84.62 | 84.62 | 84.62 | 84.62 | 84.62 | **92.31** | 84.62 | **92.31** | 69.23 | 76.92 | 76.92 | 84.62 |
| 4 | **93.33** | **93.33** | **93.33** | **93.33** | **93.33** | 100.00 | **93.33** | **93.33** | 80.00 | 100.00 | 86.67 | 100.00 |
| Avg. | 86.21 | 86.21 | 86.21 | 86.21 | 86.21 | **93.10** | 86.21 | 89.66 | 72.41 | 86.21 | 79.31 | 89.66 |
| **2-Week Forecasting Horizon** | | | | | | | | | | | | |
| 1 | - | - | - | - | - | - | - | - | - | - | - | - |
| 2 | 0.00 | 0.00 | 0.00 | 0.00 | 0.00 | **50.00** | 0.00 | **50.00** | 0.00 | 0.00 | 0.00 | 0.00 |
| 3 | 88.46 | 88.46 | 88.46 | 88.46 | 88.46 | 88.46 | 88.46 | **92.31** | 65.38 | 76.92 | 80.77 | 84.62 |
| 4 | **96.67** | **96.67** | **96.67** | **96.67** | **96.67** | 100.00 | **96.67** | 100.00 | 80.00 | **93.33** | 86.67 | 100.00 |
| Avg. | 89.66 | 89.66 | 89.66 | 89.66 | 89.66 | 93.10 | 89.66 | **94.83** | 70.69 | 82.76 | 81.03 | 89.66 |
| **3-Week Forecasting Horizon** | | | | | | | | | | | | |
| 1 | - | - | - | - | - | - | - | - | - | - | - | - |
| 2 | 0.00 | 0.00 | 0.00 | 0.00 | 0.00 | **66.67** | 0.00 | **66.67** | 0.00 | 0.00 | 0.00 | 0.00 |
| 3 | 89.74 | 89.74 | 89.74 | 89.74 | 89.74 | 87.18 | 89.74 | **92.31** | 61.54 | 71.79 | 82.05 | 82.05 |
| 4 | **97.78** | **97.78** | **97.78** | **97.78** | **97.78** | 100.00 | **97.78** | 100.00 | 80.00 | 88.89 | 84.44 | 100.00 |
| Avg. | 90.80 | 90.80 | 90.80 | 90.80 | 90.80 | 93.10 | 90.80 | **95.40** | 68.97 | 78.16 | 80.46 | 88.51 |
| **4-Week Forecasting Horizon** | | | | | | | | | | | | |
| 1 | - | - | - | - | - | - | - | - | - | - | - | - |
| 2 | 0.00 | 0.00 | 0.00 | 0.00 | 0.00 | **50.00** | 0.00 | **50.00** | 0.00 | 0.00 | 0.00 | 0.00 |
| 3 | 90.38 | **92.31** | 90.38 | **92.31** | 90.38 | 86.54 | 90.38 | **92.31** | 57.69 | 65.38 | 78.85 | 82.69 |
| 4 | **98.33** | **98.33** | **98.33** | **98.33** | **98.33** | 100.00 | **98.33** | **98.33** | 81.67 | 85.00 | 85.00 | 100.00 |
| Avg. | 91.38 | 92.24 | 91.38 | 92.24 | 91.38 | 92.24 | 91.38 | **93.97** | 68.10 | 73.28 | 79.31 | 88.79 |

[1] Data was obtained from the Our World in Data Team (OWID) [1].
[2] The time-period of the epidemic, where *1* refers to ascending phases (June – Early July 2022), *2* the peak (Mid-July 2022), *3* for the immediate declining phase (Late July – Mid-October 2022), and *4* for the tail-end of the epidemic (Late-October 2022 - Current).
[3] The models of interest include models of the spatial-wave framework (SW), and *n*-sub-epidemic frameworks (SE), a simple linear regression model, Facebook's Prophet model (Prophet), a general additive model, and an auto-regressive integrated moving average model (ARIMA). All models were calibrated with 11-weeks' worth of data at a time.
[4] Metrics are not available for the ascending phase of the epidemic in the United Kingdom, as it occurred from June to Early July 2022. Therefore, not enough observed weeks were available to conduct forecasts for the ascending phase of the epidemic.
* The bolded values indicate the best performing model(s) at each time-period for each forecasting horizon. Here, *best performing* corresponds to the model which produces coverage closest to 95%.
** Each value is shown as a percentage (%).

**Table F4.** Average Weighted Interval Scores (WIS) of the forecasts generated for the United Kingdom [1] (weeks of July 14th, 2022, through February 23rd, 2023) for each forecasting horizon (1-4 weeks), and time-period [2] of the epidemic, and averaged across all forecast periods (*Avg.*) for each model [3] of interest.

| Time Period | SW 1st Ranked | SW 2nd Ranked | SW Weighted Ensemble (2) | SW Unweighted Ensemble (2) | SE 1st Ranked | SE 2nd Ranked | SE Weighted Ensemble (2) | SE Unweighted Ensemble (2) | Simple Linear Regression | Prophet | General Additive Model | ARIMA[3] |
|---|---|---|---|---|---|---|---|---|---|---|---|---|
| **1-Week Forecasting Horizon** | | | | | | | | | | | | |
| 1 | - | - | - | - | - | - | - | - | - | - | - | - |
| 2 | 107.86 | 114.32 | 109.41 | 111.72 | 104.96 | **48.69** | 104.55 | 69.08 | 138.87 | 134.39 | 138.87 | 117.85 |
| 3 | 35.20 | 34.89 | 34.96 | 35.16 | 33.88 | 5521517.59 | **33.86** | 1786905.29 | 63.42 | 59.98 | 57.34 | 49.47 |
| 4 | 1.82 | **1.79** | 1.80 | 1.80 | 1.88 | 2.59 | 1.86 | 2.11 | 2.08 | 1.98 | 5.51 | 4.81 |
| Avg. | 20.44 | 20.51 | 20.37 | 20.55 | 19.78 | 2475166.08 | **19.74** | 801029.98 | 34.30 | 32.55 | 33.35 | 28.73 |
| **2-Week Forecasting Horizon** | | | | | | | | | | | | |
| 1 | - | - | - | - | - | - | - | - | - | - | - | - |
| 2 | 103.07 | 111.51 | 106.99 | 107.44 | 99.46 | **26.25** | 99.13 | 48.50 | 143.03 | 139.36 | 143.03 | 115.84 |
| 3 | 33.99 | 33.73 | 33.71 | 33.77 | **32.45** | 3732000.48 | 32.73 | 1235391.22 | 71.07 | 67.72 | 49.58 | 56.33 |
| 4 | 1.71 | **1.69** | 1.69 | 1.69 | 1.74 | 2.48 | 1.74 | 2.02 | 2.10 | 2.02 | 7.01 | 5.19 |
| Avg. | 19.67 | 19.84 | 19.68 | 19.72 | **18.88** | 1672967.92 | 18.99 | 553798.78 | 37.88 | 36.21 | 30.79 | 31.93 |
| **3-Week Forecasting Horizon** | | | | | | | | | | | | |
| 1 | - | - | - | - | - | - | - | - | - | - | - | - |
| 2 | 130.46 | 141.16 | 135.10 | 135.53 | 125.87 | **22.02** | 125.66 | 52.29 | 180.86 | 177.64 | 180.86 | 148.44 |
| 3 | 32.91 | 32.69 | 32.79 | 32.85 | **31.91** | 1410110.68 | 31.91 | 221825.91 | 79.66 | 77.01 | 41.13 | 61.25 |
| 4 | 1.65 | **1.63** | 1.63 | 1.64 | 1.66 | 2.50 | 1.67 | 1.96 | 2.05 | 2.00 | 8.80 | 5.33 |
| Avg. | 20.10 | 20.36 | 20.20 | 20.25 | 19.50 | 632120.64 | **19.50** | 99442.02 | 43.01 | 41.68 | 29.23 | 35.33 |
| **4-Week Forecasting Horizon** | | | | | | | | | | | | |
| 1 | - | - | - | - | - | - | - | - | - | - | - | - |
| 2 | 202.09 | 215.02 | 206.25 | 209.21 | 196.33 | **62.20** | 196.63 | 100.62 | 264.83 | 262.95 | 264.83 | 227.80 |
| 3 | 30.24 | 29.68 | 29.97 | 29.92 | **29.09** | 1882264.00 | 29.12 | 553405.17 | 84.73 | 82.86 | 41.38 | 64.45 |
| 4 | 1.58 | **1.56** | 1.56 | 1.56 | 1.60 | 2.47 | 1.60 | 1.91 | 1.87 | 1.83 | 10.47 | 5.44 |
| Avg. | 21.34 | 21.52 | 21.35 | 21.44 | **20.64** | 843776.94 | 20.66 | 248082.64 | 48.08 | 47.16 | 33.10 | 39.56 |

[1] Data was obtained from the Our World in Data Team (OWID) [1].
[2] The time-period of the epidemic, where *1* refers to ascending phases (June – Early July 2022), *2* the peak (Mid-July 2022), *3* for the immediate declining phase (Late July – Mid-October 2022), and *4* for the tail-end of the epidemic (Late-October 2022 - Current).
[3] The models of interest include models of the spatial-wave framework (SW), and *n*-sub-epidemic frameworks (SE), a simple linear regression model, Facebook's Prophet model (Prophet), a general additive model, and an auto-regressive integrated moving average model (ARIMA). All models were calibrated with 11-weeks' worth of data at a time.
[4] Metrics are not available for the ascending phase of the epidemic in the United Kingdom, as it occurred from June to Early July 2022. Therefore, not enough observed weeks were available to conduct forecasts for the ascending phase of the epidemic.
* The bolded values indicate the best performing model(s) at each time-period for each forecasting horizon. Here, *best performing* refers to the model with the lowest metric value.

**Table F5.** Skills scores[1] for overall average forecast performance metrics (weeks of July 14th, 2022, through February 23rd, 2023) comparing the ARIMA[2] model to the *n*-sub-epidemic (SE) and spatial-wave (SW) frameworks across forecasting horizons (1-4 weeks) for the United Kingdom.[3]

| **Baseline[4]** | ARIMA | | | | | | | |
|---|---|---|---|---|---|---|---|---|
| **Comparison[5]** | SW 1st Ranked | SW 2nd Ranked | SW Weighted Ensemble (2) | SW Unweighted Ensemble (2) | SE 1st Ranked | SE 2nd Ranked | SE Weighted Ensemble (2) | SE Unweighted Ensemble (2) |
| **Mean Squared Error (MSE)** | | | | | | | | |
| 1-Week | 42.14 | 40.89 | 40.79 | 41.72 | 44.46 | -681.60 | 44.46 | 45.93 |
| 2-Week | 54.47 | 53.46 | 53.95 | 54.46 | 57.03 | -312.14 | 57.03 | 58.38 |
| 3-Week | 57.21 | 55.46 | 56.96 | 55.93 | 59.24 | -24.48 | 59.24 | 69.75 |
| 4-Week | 54.14 | 51.94 | 53.01 | 53.10 | 56.85 | 15.36 | 56.85 | 72.12 |
| **Mean Absolute Error (MAE)** | | | | | | | | |
| 1-Week | 33.59 | 34.09 | 33.49 | 34.26 | 37.13 | -53.91 | 37.13 | 32.10 |
| 2-Week | 44.97 | 43.52 | 44.28 | 44.49 | 47.71 | -19.99 | 47.71 | 46.11 |
| 3-Week | 48.40 | 47.67 | 48.39 | 47.97 | 50.08 | 32.56 | 50.08 | 55.09 |
| 4-Week | 50.21 | 49.82 | 50.27 | 50.17 | 52.19 | 39.48 | 52.19 | 56.57 |
| **95% Prediction Interval Coverage (95% PI) *** | | | | | | | | |
| 1-Week | 19.22 | 16.05 | 18.82 | 15.99 | 19.19 | -176082669.26 | 21.11 | -669.19 |
| 2-Week | 28.27 | 27.34 | 27.12 | 27.42 | 30.85 | -90911449.70 | 30.17 | -244222.85 |
| 3-Week | 32.77 | 30.02 | 31.06 | 30.48 | 34.14 | -15413385.90 | 33.44 | -1276.57 |
| 4-Week | 39.38 | 37.23 | 39.31 | 38.04 | 40.45 | -39917184.88 | 40.25 | -121723.29 |
| **Weighted Interval Score (WIS)** | | | | | | | | |
| 1-Week | 28.84 | 28.60 | 29.08 | 28.48 | 31.15 | -8616262.73 | 31.27 | -2788385.57 |
| 2-Week | 38.38 | 37.86 | 38.37 | 38.24 | 40.87 | -5239830.03 | 40.53 | -1734462.16 |
| 3-Week | 43.10 | 42.36 | 42.82 | 42.69 | 44.80 | -1789100.87 | 44.80 | -281368.03 |
| 4-Week | 46.06 | 45.60 | 46.02 | 45.82 | 47.83 | -2132685.59 | 47.78 | -626969.84 |

[1] Skill scores are calculated by subtracting the mean metric score for the comparison model from the benchmark model, dividing by the mean metric score for the benchmark model and multiplying by 100. They are shown here as a percentage.
[2] Auto-regressive integrated moving average
[3] Data was obtained from the Our World in Data Team (OWID) [1].
[4] The ARIMA model used an 11-week calibration period for all forecasts.
[5] Comparison models used only a 11-week calibration period and were comprised of the 1st ranked, 2nd ranked, weighted (W) ensemble, and unweighted (UW) ensemble models from both the *n*-sub-epidemic (SE) and spatial-wave (SW) frameworks.
* Skill scores for 95% PI coverage are based on average Winkler scores.

**Table G1.** Average Mean Squared Error (MSE) of the forecasts generated for US(CDC) [1] (weeks of July 14th, 2022, through February 23rd, 2023) for each forecasting horizon (1-4 weeks), and time-period [2] of the epidemic, and averaged across all forecast periods (*Avg.*) for each model [3] of interest.

| Time Period | SW 1st Ranked | SW 2nd Ranked | SW Weighted Ensemble (2) | SW Unweighted Ensemble (2) | SE 1st Ranked | SE 2nd Ranked | SE Weighted Ensemble (2) | SE Unweighted Ensemble (2) | Simple Linear Regression | Prophet | General Additive Model | ARIMA[3] |
|---|---|---|---|---|---|---|---|---|---|---|---|---|
| **1-Week Forecasting Horizon** | | | | | | | | | | | | |
| 1 | 1144934.62 | 1081542.50 | 1111350.35 | 1115865.97 | 657273.29 | 112288035.65 | 657273.29 | 1155126.24 | 730248.33 | 732730.78 | **4446.23** | 7425.68 |
| 2 | 63198.60 | 64103.92 | 64103.92 | 63198.60 | 18798.38 | 1793155.30 | 18798.38 | 625756.66 | 1019.35 | **906.37** | 327160.13 | 323761.00 |
| 3 | 33882.39 | 30186.17 | 30899.04 | 29264.32 | **16791.33** | 228128.64 | **16791.33** | 18793.96 | 774448.21 | 771849.67 | 36848.39 | 65313.29 |
| 4 | **419.86** | 522.98 | 445.44 | 467.71 | 526.95 | 1241.57 | 526.95 | 592.91 | 3427.16 | 3426.97 | 441.66 | 550.16 |
| Avg. | 99978.78 | 93634.40 | 96056.67 | 95442.65 | 55423.30 | 7932127.61 | 55423.30 | 111815.23 | 478860.65 | 477594.22 | **32070.46** | 47900.92 |
| **2-Week Forecasting Horizon** | | | | | | | | | | | | |
| 1 | 6776418.38 | 6703460.53 | 6814981.54 | 6700138.38 | 6780661.15 | 298533877.93 | 6780661.15 | 9374967.28 | 764395.05 | 767134.28 | 57094.24 | **52422.05** |
| 2 | 73977.65 | 73808.80 | 73808.80 | 73379.28 | **14512.26** | 2072407.77 | **14512.26** | 782390.81 | 330654.28 | 328876.80 | 1267369.15 | 1252645.00 |
| 3 | 24963.61 | **17735.58** | 21668.64 | 19403.57 | 24390.39 | 751824.75 | 24390.39 | 26140.82 | 1104279.94 | 1101058.11 | 98694.36 | 148461.19 |
| 4 | **431.56** | 547.15 | 502.07 | 471.23 | 530.14 | 1540.70 | 530.14 | 621.52 | 2955.48 | 2955.38 | 793.94 | 837.83 |
| Avg. | 483811.99 | 474826.57 | 484672.10 | 475476.74 | 481771.80 | 21075336.78 | 481771.80 | 688165.30 | 674395.80 | 672745.82 | **102365.75** | 129008.50 |
| **3-Week Forecasting Horizon** | | | | | | | | | | | | |
| 1 | 28499933.45 | 28860787.36 | 28430970.57 | 28073177.64 | 30038551.79 | 798283018.25 | 30038551.79 | 41011716.33 | 663277.51 | 665699.84 | 411954.11 | **397795.47** |
| 2 | 295057.16 | 292669.43 | 292669.43 | 292383.08 | **146444.42** | 2364349.86 | **146444.42** | 1152953.68 | 808506.80 | 805180.75 | 2363759.03 | 2333230.00 |
| 3 | 25861.15 | **19855.23** | 21228.06 | 20942.88 | 32678.40 | 1003870.41 | 32678.40 | 30218.32 | 1507201.08 | 1503286.03 | 168394.86 | 277286.80 |
| 4 | **382.52** | 488.77 | 445.79 | 440.11 | 465.67 | 1955.47 | 465.67 | 591.05 | 2551.59 | 2551.52 | 1007.04 | 1085.52 |
| Avg. | 1990087.16 | 2011614.33 | 1982714.39 | 1957869.85 | 2094864.10 | 55690064.40 | 2094864.10 | 2885027.24 | 906061.90 | 903954.21 | **203174.32** | 261250.52 |
| **4-Week Forecasting Horizon** | | | | | | | | | | | | |
| 1 | 94742142.52 | 98000333.87 | 96624755.91 | 99506777.40 | 99199381.81 | 2084740949.24 | 99199381.81 | 140962755.83 | **583269.39** | 585010.51 | 1114393.94 | 1100045.83 |
| 2 | 463679.55 | 460270.98 | 460270.98 | 460056.22 | **269995.82** | 2287407.23 | **269995.82** | 1254254.67 | 1800909.87 | 1795444.81 | 4214878.73 | 4160178.75 |
| 3 | 37263.09 | **31009.53** | 32214.51 | 32540.57 | 40123.81 | 878321.52 | 40123.81 | 36583.80 | 1955102.20 | 1950449.68 | 235036.13 | 421262.51 |
| 4 | **356.78** | 455.64 | 412.15 | 419.04 | 415.91 | 3724.50 | 415.91 | 544.94 | 2192.84 | 2192.79 | 1089.70 | 1194.52 |
| Avg. | 6570611.79 | 6791780.95 | 6697563.33 | 6896498.29 | 6872927.57 | 144339989.48 | 6872927.57 | 9785191.60 | 1181759.04 | 1179123.74 | **352246.27** | 452152.27 |

[1] Data was obtained from the Centers for Disease Control (CDC) website [2].

[2] The time-period of the epidemic, where *1* refers to ascending phases (May- Mid July 2022), *2* the peak (Late July 2022), *3* for the immediate declining phase (Early August – Early November 2022), and *4* for the tail-end of the epidemic (Mid-November 2022 - Current).

[3] The models of interest include models of the spatial-wave framework (SW), and *n*-sub-epidemic frameworks (SE), a simple linear regression model, Facebook's Prophet model (Prophet), a general additive model, and an auto-regressive integrated moving average model (ARIMA). All models were calibrated with 11-weeks' worth of data at a time.

* The bolded values indicate the best performing model(s) at each time-period for each forecasting horizon. Here, *best performing* refers to the model with the lowest metric value.

**Table G2.** Average Mean Absolute Error (MAE) of the forecasts generated for the US(CDC) [1] (weeks of July 14th, 2022, through February 23rd, 2023) for each forecasting horizon (1-4 weeks), and time-period [2] of the epidemic, and averaged across all forecast periods (*Avg.*) for each model [3] of interest.

| Time Period | SW 1st Ranked | SW 2nd Ranked | SW Weighted Ensemble (2) | SW Unweighted Ensemble (2) | SE 1st Ranked | SE 2nd Ranked | SE Weighted Ensemble (2) | SE Unweighted Ensemble (2) | Simple Linear Regression | Prophet | General Additive Model | ARIMA[3] |
|---|---|---|---|---|---|---|---|---|---|---|---|---|
| **1-Week Forecasting Horizon** | | | | | | | | | | | | |
| 1 | 1069.72 | 1038.45 | 1053.82 | 1056.01 | 696.56 | 7687.71 | 696.56 | 923.02 | 849.09 | 850.49 | **66.18** | 85.91 |
| 2 | 251.39 | 253.19 | 253.19 | 251.39 | 137.11 | 1339.09 | 137.11 | 791.05 | 31.93 | **30.11** | 571.98 | 569.00 |
| 3 | 122.58 | 120.56 | 120.51 | 119.87 | 96.64 | 265.46 | 96.64 | **94.91** | 676.52 | 675.43 | 117.62 | 163.62 |
| 4 | 17.47 | 19.53 | 18.38 | 18.76 | 20.54 | 27.53 | 20.54 | 18.94 | 51.92 | 51.91 | **16.94** | 19.93 |
| Avg. | 156.10 | 153.60 | 154.23 | 154.10 | 113.17 | 732.32 | 113.17 | 149.83 | 450.81 | 450.24 | **95.02** | 122.69 |
| **2-Week Forecasting Horizon** | | | | | | | | | | | | |
| 1 | 2012.07 | 1960.53 | 2004.06 | 2001.44 | 1879.01 | 11931.71 | 1879.01 | 2217.60 | 820.23 | 821.71 | **177.08** | 186.13 |
| 2 | 271.48 | 271.28 | 271.28 | 270.44 | **119.80** | 1436.30 | **119.80** | 880.37 | 422.25 | 420.28 | 1028.89 | 1023.00 |
| 3 | 101.22 | **88.16** | 96.13 | 94.18 | 109.48 | 370.77 | 109.48 | 111.58 | 769.48 | 768.34 | 184.58 | 236.94 |
| 4 | **17.17** | 19.25 | 18.76 | 18.16 | 19.83 | 30.92 | 19.83 | 19.57 | 48.06 | 48.06 | 22.20 | 24.21 |
| Avg. | 209.89 | 199.84 | 207.07 | 205.58 | 200.96 | 1087.63 | 200.96 | 251.61 | 512.24 | 511.64 | **157.18** | 187.19 |
| **3-Week Forecasting Horizon** | | | | | | | | | | | | |
| 1 | 3473.94 | 3367.26 | 3385.94 | 3349.81 | 3586.51 | 18223.86 | 3586.51 | 4154.79 | 753.07 | 754.27 | **458.65** | 464.10 |
| 2 | 466.16 | 464.76 | 464.76 | 464.20 | **292.97** | 1529.89 | **292.97** | 1041.79 | 724.25 | 722.13 | 1397.46 | 1388.67 |
| 3 | 101.57 | **93.23** | 95.43 | 96.80 | 117.79 | 416.22 | 117.79 | 114.44 | 860.25 | 859.06 | 247.45 | 314.42 |
| 4 | **16.78** | 18.70 | 17.77 | 17.93 | 18.94 | 32.97 | 18.94 | 20.20 | 44.31 | 44.30 | 23.87 | 26.84 |
| Avg. | 317.48 | 306.14 | 308.32 | 306.62 | 328.97 | 1550.58 | 328.97 | 392.56 | 566.81 | 566.16 | **224.57** | 262.62 |
| **4-Week Forecasting Horizon** | | | | | | | | | | | | |
| 1 | 5713.95 | 5617.01 | 5721.76 | 5666.22 | 6013.69 | 27511.27 | 6013.69 | 7048.10 | **694.90** | 695.98 | 792.13 | 795.75 |
| 2 | 595.51 | 593.64 | 593.64 | 593.22 | **419.09** | 1505.93 | **419.09** | 1080.54 | 1089.66 | 1087.39 | 1829.45 | 1817.75 |
| 3 | 112.51 | **107.24** | 109.40 | 109.53 | 124.82 | 410.63 | 124.82 | 120.77 | 946.03 | 944.98 | 299.21 | 374.25 |
| 4 | **16.23** | 18.00 | 17.62 | 17.58 | 17.95 | 36.84 | 17.95 | 19.42 | 40.83 | 40.83 | 24.52 | 28.31 |
| Avg. | 482.27 | 473.23 | 481.51 | 477.72 | 504.24 | 2188.51 | 504.24 | 596.67 | 621.52 | 620.94 | **291.25** | 333.81 |

[1] Data was obtained from the Centers for Disease Control (CDC) website [2].
[2] The time-period of the epidemic, where *1* refers to ascending phases (May- Mid July 2022), *2* the peak (Late July 2022), *3* for the immediate declining phase (Early August – Early November 2022), and *4* for the tail-end of the epidemic (Mid-November 2022 - Current).
[3] The models of interest include models of the spatial-wave framework (SW), and *n*-sub-epidemic frameworks (SE), a simple linear regression model, Facebook's Prophet model (Prophet), a general additive model, and an auto-regressive integrated moving average model (ARIMA). All models were calibrated with 11-weeks' worth of data at a time.
* The bolded values indicate the best performing model(s) at each time-period for each forecasting horizon. Here, *best performing* refers to the model with the lowest metric value.

**Table G3.** Average 95% Prediction Interval Coverage (95% PI Coverage) of the forecasts generated for the US(CDC) [1] (weeks of July 14th, 2022, through February 23rd, 2023) for each forecasting horizon (1-4 weeks), and time-period [2] of the epidemic, and averaged across all forecast periods (*Avg.*) for each model [3] of interest.

| Time Period | SW 1st Ranked | SW 2nd Ranked | SW Weighted Ensemble (2) | SW Unweighted Ensemble (2) | SE 1st Ranked | SE 2nd Ranked | SE Weighted Ensemble (2) | SE Unweighted Ensemble (2) | Simple Linear Regression | Prophet | General Additive Model | ARIMA[3] |
|---|---|---|---|---|---|---|---|---|---|---|---|---|
| **1-Week Forecasting Horizon** | | | | | | | | | | | | |
| 1 | 0.00 | 0.00 | 0.00 | 0.00 | 0.00 | 50.00 | 0.00 | 0.00 | 0.00 | 50.00 | 50.00 | **100.00** |
| 2 | **100.00** | **100.00** | **100.00** | **100.00** | **100.00** | 0.00 | **100.00** | **100.00** | **100.00** | **100.00** | 0.00 | 0.00 |
| 3 | 100.00 | 100.00 | 100.00 | 100.00 | 87.50 | **93.75** | 87.50 | 100.00 | 18.75 | 37.50 | 81.25 | **93.75** |
| 4 | **100.00** | **100.00** | **100.00** | **100.00** | **100.00** | **90.00** | **100.00** | **100.00** | 10.00 | 50.00 | 80.00 | **90.00** |
| Avg. | **93.10** | **93.10** | **93.10** | **93.10** | 86.21 | 86.21 | 86.21 | **93.10** | 17.24 | 44.83 | 75.86 | 89.66 |
| **2-Week Forecasting Horizon** | | | | | | | | | | | | |
| 1 | 25.00 | 25.00 | 25.00 | 25.00 | 0.00 | 25.00 | 0.00 | 0.00 | 25.00 | 50.00 | 50.00 | **100.00** |
| 2 | **100.00** | **100.00** | **100.00** | **100.00** | **100.00** | 0.00 | **100.00** | **100.00** | 50.00 | 50.00 | 0.00 | 0.00 |
| 3 | 100.00 | 100.00 | 100.00 | 100.00 | 84.38 | **90.63** | 84.38 | 100.00 | 18.75 | 28.13 | 81.25 | **90.63** |
| 4 | 100.00 | 100.00 | 100.00 | 100.00 | 100.00 | 90.00 | 100.00 | 100.00 | 10.00 | 30.00 | 85.00 | **95.00** |
| Avg. | **94.83** | **94.83** | **94.83** | **94.83** | 84.48 | 82.76 | 84.48 | 93.10 | 17.24 | 31.03 | 77.59 | 89.66 |
| **3-Week Forecasting Horizon** | | | | | | | | | | | | |
| 1 | 33.33 | 33.33 | 33.33 | 33.33 | 0.00 | 16.67 | 0.00 | 0.00 | 33.33 | 50.00 | 33.33 | **83.33** |
| 2 | **100.00** | **100.00** | **100.00** | **100.00** | **100.00** | 0.00 | **100.00** | **100.00** | 33.33 | 33.33 | 0.00 | 0.00 |
| 3 | **100.00** | **100.00** | **100.00** | **100.00** | 83.33 | 89.58 | 83.33 | **100.00** | 16.67 | 22.92 | 81.25 | 87.50 |
| 4 | 100.00 | 100.00 | 100.00 | 100.00 | 100.00 | 90.00 | 100.00 | 100.00 | 6.67 | 20.00 | 83.33 | **96.67** |
| Avg. | **95.40** | **95.40** | **95.40** | **95.40** | 83.91 | 81.61 | 83.91 | 93.10 | 14.94 | 24.14 | 75.86 | 87.36 |
| **4-Week Forecasting Horizon** | | | | | | | | | | | | |
| 1 | 37.50 | 37.50 | 37.50 | 37.50 | 0.00 | 12.50 | 0.00 | 0.00 | 50.00 | **62.50** | 25.00 | **62.50** |
| 2 | **100.00** | **100.00** | **100.00** | **100.00** | **100.00** | 0.00 | **100.00** | **100.00** | 25.00 | 25.00 | 0.00 | 0.00 |
| 3 | 100.00 | 100.00 | 100.00 | 100.00 | 82.81 | 87.50 | 82.81 | **98.44** | 15.63 | 18.75 | 81.25 | 85.94 |
| 4 | 100.00 | 100.00 | 100.00 | 100.00 | 100.00 | 90.00 | 100.00 | 100.00 | 5.00 | 17.50 | 85.00 | **97.50** |
| Avg. | **95.69** | **95.69** | **95.69** | **95.69** | 83.62 | 80.17 | 83.62 | 92.24 | 14.66 | 21.55 | 75.86 | 85.34 |

[1] Data was obtained from the Centers for Disease Control (CDC) website [2].
[2] The time-period of the epidemic, where *1* refers to ascending phases (May- Mid July 2022), *2* the peak (Late July 2022), *3* for the immediate declining phase (Early August – Early November 2022), and *4* for the tail-end of the epidemic (Mid-November 2022 - Current).
[3] The models of interest include models of the spatial-wave framework (SW), and *n*-sub-epidemic frameworks (SE), a simple linear regression model, Facebook's Prophet model (Prophet), a general additive model, and an auto-regressive integrated moving average model (ARIMA). All models were calibrated with 11-weeks' worth of data at a time.
* The bolded values indicate the best performing model(s) at each time-period for each forecasting horizon. Here, *best performing* corresponds to the model which produces coverage closest to 95%.
** Each value is shown as a percentage (%).

**Table G4.** Average Weighted Interval Scores (WIS) of the forecasts generated for the US(CDC) [1] (weeks of July 14th, 2022, through February 23rd, 2023) for each forecasting horizon (1-4 weeks), and time-period [2] of the epidemic, and averaged across all forecast periods (*Avg.*) for each model [3] of interest.

| Time Period | SW 1st Ranked | SW 2nd Ranked | SW Weighted Ensemble (2) | SW Unweighted Ensemble (2) | SE 1st Ranked | SE 2nd Ranked | SE Weighted Ensemble (2) | SE Unweighted Ensemble (2) | Simple Linear Regression | Prophet | General Additive Model | ARIMA[3] |
|---|---|---|---|---|---|---|---|---|---|---|---|---|
| **1-Week Forecasting Horizon** | | | | | | | | | | | | |
| 1 | 840.64 | 843.28 | 843.76 | 839.71 | 535.41 | 7216.23 | 536.16 | 2397.51 | 639.17 | 573.07 | **39.60** | 45.84 |
| 2 | 136.53 | 135.35 | 134.05 | 136.50 | 102.58 | 1160.43 | 102.64 | 388.24 | **58.34** | 79.19 | 521.82 | 469.53 |
| 3 | 80.95 | 79.94 | 81.53 | 79.41 | **65.38** | 217.13 | 65.48 | 99.81 | 533.87 | 485.55 | 91.65 | 108.13 |
| 4 | 19.95 | 19.73 | 19.97 | 19.84 | 12.08 | 20.60 | 12.08 | 14.42 | 43.58 | 37.47 | **10.70** | 12.55 |
| Avg. | 114.22 | 113.73 | 114.68 | 113.27 | 80.70 | 664.59 | 80.81 | 238.77 | 355.67 | 323.07 | **74.98** | 83.34 |
| **2-Week Forecasting Horizon** | | | | | | | | | | | | |
| 1 | 1585.16 | 1600.16 | 1597.45 | 1587.76 | 1568.64 | 12576.66 | 1567.14 | 4505.62 | 614.55 | 562.45 | 131.97 | **104.15** |
| 2 | 173.23 | 168.62 | 172.87 | 169.67 | **134.34** | 1231.64 | 136.29 | 420.48 | 322.22 | 305.92 | 957.41 | 862.05 |
| 3 | 70.10 | **67.03** | 68.79 | 67.84 | 76.01 | 271.70 | 76.07 | 119.70 | 624.44 | 585.46 | 143.13 | 159.58 |
| 4 | 19.60 | 19.47 | 19.50 | 19.52 | **12.02** | 21.53 | 12.04 | 14.72 | 43.01 | 38.92 | 13.41 | 14.84 |
| Avg. | 160.73 | 159.87 | 160.81 | 159.51 | 158.89 | 1067.15 | 158.90 | 396.35 | 412.84 | 385.77 | **125.71** | 130.07 |
| **3-Week Forecasting Horizon** | | | | | | | | | | | | |
| 1 | 2836.72 | 2891.74 | 2869.49 | 2867.57 | 3022.99 | 22706.51 | 3024.18 | 7929.31 | 546.07 | 509.58 | 396.36 | **283.70** |
| 2 | 289.47 | 282.25 | 280.22 | 286.09 | 203.38 | 1318.70 | **201.53** | 504.17 | 573.01 | 550.11 | 1304.51 | 1157.30 |
| 3 | 70.02 | 68.35 | 68.32 | **68.01** | 85.00 | 281.97 | 85.12 | 126.46 | 712.09 | 682.07 | 184.03 | 211.51 |
| 4 | 19.27 | 19.16 | 19.19 | 19.26 | 11.86 | 21.75 | **11.85** | 14.55 | 40.83 | 37.93 | 15.05 | 16.79 |
| Avg. | 250.89 | 253.48 | 251.87 | 251.80 | 266.48 | 1774.51 | 266.56 | 639.02 | 464.38 | 443.51 | **179.04** | 181.96 |
| **4-Week Forecasting Horizon** | | | | | | | | | | | | |
| 1 | 4735.45 | 4864.20 | 4802.50 | 4801.34 | 5047.85 | 42858.55 | 5048.06 | 15819.28 | 489.34 | **464.46** | 713.52 | 504.70 |
| 2 | 387.28 | 373.08 | 383.42 | 383.90 | **256.12** | 1296.96 | 256.30 | 551.11 | 905.91 | 890.48 | 1714.97 | 1508.01 |
| 3 | 76.64 | 75.16 | **74.06** | 74.63 | 92.64 | 287.09 | 92.43 | 137.03 | 792.48 | 770.07 | 219.88 | 256.82 |
| 4 | 18.84 | 18.58 | 18.60 | 18.77 | 11.52 | 23.12 | **11.50** | 14.57 | 38.22 | 36.15 | 16.08 | 18.52 |
| Avg. | 388.72 | 396.20 | 391.70 | 392.01 | 412.04 | 3166.85 | 411.94 | 1190.62 | 515.39 | 500.07 | 235.20 | **234.89** |

[1] Data was obtained from the Centers for Disease Control (CDC) website [2].
[2] The time-period of the epidemic, where *1* refers to ascending phases (May- Mid July 2022), *2* the peak (Late July 2022), *3* for the immediate declining phase (Early August – Early November 2022), and *4* for the tail-end of the epidemic (Mid-November 2022 - Current).
[3] The models of interest include models of the spatial-wave framework (SW), and *n*-sub-epidemic frameworks (SE), a simple linear regression model, Facebook's Prophet model (Prophet), a general additive model, and an auto-regressive integrated moving average model (ARIMA). All models were calibrated with 11-weeks' worth of data at a time.
* The bolded values indicate the best performing model(s) at each time-period for each forecasting horizon. Here, *best performing* refers to the model with the lowest metric value.

**Table G5.** Skills scores[1] for overall average forecast performance metrics (weeks of July 14th, 2022, through February 23rd, 2023) comparing the ARIMA[2] model to the *n*-sub-epidemic (SE) and spatial-wave (SW) frameworks across forecasting horizons (1-4 weeks) for the US(CDC).[3]

| **Baseline**[4] | ARIMA | | | | | | | |
|---|---|---|---|---|---|---|---|---|
| **Comparison**[5] | SW 1st Ranked | SW 2nd Ranked | SW Weighted Ensemble (2) | SW Unweighted Ensemble (2) | SE 1st Ranked | SE 2nd Ranked | SE Weighted Ensemble (2) | SE Unweighted Ensemble (2) |
| **Mean Squared Error (MSE)** | | | | | | | | |
| 1-Week | -108.72 | -95.48 | -100.53 | -99.25 | -15.70 | -16459.45 | -15.70 | -133.43 |
| 2-Week | -275.02 | -268.06 | -275.69 | -268.56 | -273.44 | -16236.39 | -273.44 | -433.43 |
| 3-Week | -661.75 | -669.99 | -658.93 | -649.42 | -701.86 | -21216.73 | -701.86 | -1004.31 |
| 4-Week | -1353.19 | -1402.10 | -1381.26 | -1425.26 | -1420.05 | -31822.87 | -1420.05 | -2064.14 |
| **Mean Absolute Error (MAE)** | | | | | | | | |
| 1-Week | -27.23 | -25.20 | -25.71 | -25.60 | 7.76 | -496.89 | 7.76 | -22.12 |
| 2-Week | -12.13 | -6.76 | -10.62 | -9.82 | -7.36 | -481.03 | -7.36 | -34.41 |
| 3-Week | -20.89 | -16.57 | -17.40 | -16.75 | -25.26 | -490.43 | -25.26 | -49.48 |
| 4-Week | -44.48 | -41.77 | -44.25 | -43.11 | -51.06 | -555.62 | -51.06 | -78.75 |
| **95% Prediction Interval Coverage (95% PI) *** | | | | | | | | |
| 1-Week | -51.42 | -59.44 | -55.60 | -53.33 | -9.17 | -578.05 | -10.62 | -235.90 |
| 2-Week | -55.77 | -70.44 | -60.61 | -61.24 | -70.11 | -461.72 | -70.03 | -308.34 |
| 3-Week | -79.13 | -98.17 | -87.46 | -86.13 | -109.57 | -510.11 | -109.74 | -402.75 |
| 4-Week | -106.91 | -131.29 | -117.98 | -112.12 | -144.98 | -701.61 | -145.39 | -602.36 |
| **Weighted Interval Score (WIS)** | | | | | | | | |
| 1-Week | -37.06 | -36.47 | -37.61 | -35.91 | 3.17 | -697.44 | 3.04 | -186.50 |
| 2-Week | -23.57 | -22.91 | -23.63 | -22.64 | -22.16 | -720.45 | -22.17 | -204.72 |
| 3-Week | -37.89 | -39.31 | -38.42 | -38.38 | -46.45 | -875.24 | -46.50 | -251.19 |
| 4-Week | -65.49 | -68.68 | -66.76 | -66.89 | -75.42 | -1248.24 | -75.38 | -406.89 |

[1] Skill scores are calculated by subtracting the mean metric score for the comparison model from the benchmark model, dividing by the mean metric score for the benchmark model and multiplying by 100. They are shown here as a percentage.
[2] Auto-regressive integrated moving average
[3] Data was obtained from the Centers for Disease Control (CDC) website [2].
[4] The ARIMA model used an 11-week calibration period for all forecasts.
[5] Comparison models used only a 11-week calibration period and were comprised of the 1st ranked, 2nd ranked, weighted (W) ensemble, and unweighted (UW) ensemble models from both the *n*-sub-epidemic (SE) and spatial-wave (SW) frameworks.
* Skill scores for 95% PI coverage are based on average Winkler scores.

**Table H1.** Average Mean Squared Error (MSE) of the forecasts generated for the US(OWID) [1] (weeks of July 14th, 2022, through February 23rd, 2023) for each forecasting horizon (1-4 weeks), and time-period [2] of the epidemic, and averaged across all forecast periods (*Avg.*) for each model [3] of interest.

| Time Period | SW 1st Ranked | SW 2nd Ranked | SW Weighted Ensemble (2) | SW Unweighted Ensemble (2) | SE 1st Ranked | SE 2nd Ranked | SE Weighted Ensemble (2) | SE Unweighted Ensemble (2) | Simple Linear Regression | Prophet | General Additive Model | ARIMA[3] |
|---|---|---|---|---|---|---|---|---|---|---|---|---|
| **1-Week Forecasting Horizon** | | | | | | | | | | | | |
| 1 | 113839.48 | 112579.08 | 110719.36 | 112193.13 | 58592.59 | 35379.45 | 58592.59 | **31441.13** | 1229427.23 | 1229873.74 | 330779.84 | 429149.01 |
| 2 | 710298.60 | 712773.23 | 713556.10 | 693382.22 | **5153.29** | 103113.69 | **5153.29** | 90398.35 | 403617.64 | 401955.34 | 544740.74 | 544644.00 |
| 3 | 103230.32 | 103124.32 | 103484.41 | 103646.29 | 52915.82 | 56272.79 | 52915.82 | **43588.74** | 909592.71 | 910173.24 | 136487.59 | 196766.72 |
| 4 | **1191.77** | 1216.04 | 1196.15 | 1213.28 | 1722.43 | 1908.54 | 1722.43 | 1599.70 | 3898.81 | 3895.83 | 3758.00 | 3130.78 |
| Avg. | 97478.48 | 97338.17 | 97301.84 | 96903.50 | 37929.53 | 40009.12 | 37929.53 | **31944.15** | 686413.81 | 686737.54 | 140749.16 | 187398.41 |
| **2-Week Forecasting Horizon** | | | | | | | | | | | | |
| 1 | 458505.28 | 477250.69 | 464739.27 | 470098.97 | 425482.26 | 389806.92 | 425482.26 | **373429.23** | 1418918.89 | 1418995.84 | 722822.86 | 768536.87 |
| 2 | 727230.77 | 728928.85 | 723275.30 | 731168.50 | **31437.86** | 157865.86 | **31437.86** | 155973.68 | 1394871.91 | 1391672.61 | 1422257.00 | 1549124.00 |
| 3 | 102974.12 | 103461.58 | 103638.04 | 103713.21 | 71679.36 | 54552.49 | 71679.36 | **52573.18** | 1230126.27 | 1231126.39 | 131562.29 | 258470.52 |
| 4 | **1168.45** | 1200.34 | 1189.94 | 1185.29 | 1881.24 | 2280.54 | 1881.24 | 1883.30 | 3838.97 | 3837.48 | 3806.50 | 3526.43 |
| Avg. | 145454.67 | 148376.54 | 146550.37 | 147602.00 | 99837.47 | 89937.16 | 99837.47 | **86411.30** | 923561.37 | 924013.03 | 222379.21 | 303000.39 |
| **3-Week Forecasting Horizon** | | | | | | | | | | | | |
| 1 | 1095258.18 | 1127947.01 | 1111273.27 | 1101606.59 | 1099386.98 | 997818.14 | 1099386.98 | **979218.95** | 1680100.46 | 1679694.60 | 1734781.05 | 1603621.00 |
| 2 | 883926.69 | 885916.69 | 881686.67 | 889338.78 | **178196.27** | 270020.83 | **178196.27** | 259132.17 | 2480954.48 | 2476294.44 | 2278752.92 | 2589389.67 |
| 3 | 97033.29 | 97665.47 | 97641.54 | 96720.81 | 79356.27 | 80944.54 | 79356.27 | **60975.43** | 1615256.02 | 1616554.62 | 175118.13 | 385448.85 |
| 4 | **1212.32** | 1237.15 | 1232.27 | 1228.81 | 1865.93 | 1865.01 | 1865.93 | 1757.10 | 3745.02 | 3744.03 | 3707.72 | 4164.55 |
| Avg. | 235420.37 | 240353.43 | 237893.19 | 236314.78 | 202081.79 | 192114.69 | 202081.79 | **178126.61** | 1209496.99 | 1209996.51 | 415497.55 | 524288.68 |
| **4-Week Forecasting Horizon** | | | | | | | | | | | | |
| 1 | 1663940.43 | 1701838.97 | 1684230.83 | 1681923.27 | 1714340.44 | 1483370.80 | 1714340.44 | **1467788.88** | 1842487.44 | 1841587.23 | 3376492.41 | 3016906.84 |
| 2 | 798325.10 | 800043.63 | 793033.89 | 805814.10 | **199793.54** | 226111.29 | **199793.54** | 218294.48 | 4479463.64 | 4472761.76 | 3879338.62 | 4487662.50 |
| 3 | 96080.97 | 96597.22 | 96490.15 | 96227.59 | 89954.80 | 81021.07 | 89954.80 | **65338.02** | 2005989.14 | 2007575.46 | 182052.58 | 455980.12 |
| 4 | **1033.52** | 1057.09 | 1043.43 | 1050.37 | 1638.61 | 1576.44 | 1638.61 | 1587.14 | 3248.52 | 3247.77 | 3231.64 | 4297.36 |
| Avg. | 310332.77 | 315910.75 | 313177.48 | 313156.95 | 293432.45 | 257535.97 | 293432.45 | **246467.43** | 1516249.23 | 1516768.97 | 700827.68 | 823632.78 |

[1] Data was obtained from the Our World in Data Team (OWID) [1].

[2] The time-period of the epidemic, where *1* refers to ascending phases (May – Early August 2022), *2* the peak (Mid-August 2022), *3* for the immediate declining phase (Mid-August – Mid-November 2022), and *4* for the tail-end of the epidemic (Mid-November 2022 - Current).

[3] The models of interest include models of the spatial-wave framework (SW), and *n*-sub-epidemic frameworks (SE), a simple linear regression model, Facebook's Prophet model (Prophet), a general additive model, and an auto-regressive integrated moving average model (ARIMA). All models were calibrated with 11-weeks' worth of data at a time.

* The bolded values indicate the best performing model(s) at each time-period for each forecasting horizon. Here, *best performing* refers to the model with the lowest metric value.

**Table H2.** Average Mean Absolute Error (MAE) of the forecasts generated for the US(OWID) [1] (weeks of July 14th, 2022, through February 23rd, 2023) for each forecasting horizon (1-4 weeks), and time-period [2] of the epidemic, and averaged across all forecast periods (*Avg.*) for each model [3] of interest.

| Time Period | SW 1st Ranked | SW 2nd Ranked | SW Weighted Ensemble (2) | SW Unweighted Ensemble (2) | SE 1st Ranked | SE 2nd Ranked | SE Weighted Ensemble (2) | SE Unweighted Ensemble (2) | Simple Linear Regression | Prophet | General Additive Model | ARIMA[3] |
|---|---|---|---|---|---|---|---|---|---|---|---|---|
| **1-Week Forecasting Horizon** | | | | | | | | | | | | |
| 1 | 259.68 | 266.46 | 259.51 | 261.39 | 203.27 | **143.41** | 203.27 | 151.89 | 1031.45 | 1031.83 | 522.18 | 588.70 |
| 2 | 842.79 | 844.26 | 844.72 | 832.70 | **71.79** | 321.11 | **71.79** | 300.66 | 635.31 | 634.00 | 738.07 | 738.00 |
| 3 | 239.99 | 237.56 | 238.44 | 239.54 | 170.00 | 194.50 | 170.00 | **159.33** | 686.79 | 687.10 | 304.96 | 379.06 |
| 4 | **27.48** | 27.74 | 27.49 | 27.67 | 33.82 | 37.67 | 33.82 | 34.09 | 56.70 | 56.65 | 52.51 | 48.15 |
| Avg. | 204.87 | 204.59 | 204.06 | 204.56 | 133.64 | 148.55 | 133.64 | **128.63** | 558.73 | 558.90 | 280.21 | 329.07 |
| **2-Week Forecasting Horizon** | | | | | | | | | | | | |
| 1 | 558.98 | 570.10 | 562.18 | 566.70 | 523.49 | 449.10 | 523.49 | **441.12** | 1084.86 | 1084.96 | 758.91 | 750.77 |
| 2 | 852.62 | 853.61 | 850.33 | 854.95 | **153.03** | 391.10 | **153.03** | 388.13 | 1090.01 | 1088.59 | 1127.28 | 1168.00 |
| 3 | 233.87 | 233.48 | 233.35 | 234.18 | 197.78 | 188.72 | 197.78 | **175.59** | 764.72 | 765.06 | 297.81 | 399.72 |
| 4 | **26.63** | 27.05 | 26.80 | 26.77 | 33.32 | 37.50 | 33.32 | 34.03 | 55.41 | 55.39 | 54.25 | 52.65 |
| Avg. | 242.88 | 244.35 | 243.00 | 244.23 | 195.80 | 189.90 | 195.80 | **180.49** | 624.42 | 624.57 | 322.82 | 378.89 |
| **3-Week Forecasting Horizon** | | | | | | | | | | | | |
| 1 | 848.51 | 860.07 | 853.74 | 851.13 | 838.16 | 744.47 | 838.16 | **713.40** | 1156.67 | 1156.69 | 1115.31 | 1038.34 |
| 2 | 933.06 | 934.14 | 931.87 | 935.96 | **330.57** | 495.09 | **330.57** | 476.52 | 1445.71 | 1444.18 | 1417.50 | 1499.00 |
| 3 | 221.29 | 222.09 | 221.91 | 221.12 | 201.09 | 214.46 | 201.09 | **183.26** | 843.31 | 843.70 | 319.74 | 445.34 |
| 4 | **27.24** | 27.51 | 27.42 | 27.42 | 33.97 | 34.14 | 33.97 | 33.75 | 53.98 | 53.97 | 52.82 | 56.23 |
| Avg. | 278.82 | 280.96 | 279.89 | 279.23 | 247.32 | 247.50 | 247.32 | **225.25** | 689.56 | 689.72 | 393.70 | 456.13 |
| **4-Week Forecasting Horizon** | | | | | | | | | | | | |
| 1 | 1059.67 | 1071.05 | 1065.00 | 1064.92 | 1070.20 | 929.02 | 1070.20 | **897.75** | 1208.25 | 1207.90 | 1492.19 | 1365.32 |
| 2 | 883.62 | 884.65 | 880.69 | 887.85 | **376.31** | 448.00 | **376.31** | 435.50 | 1893.41 | 1891.76 | 1799.72 | 1922.00 |
| 3 | 214.06 | 214.53 | 214.53 | 214.15 | 204.97 | 216.65 | 204.97 | **180.52** | 909.57 | 909.98 | 328.63 | 456.96 |
| 4 | **25.22** | 25.53 | 25.30 | 25.42 | 31.81 | 30.26 | 31.81 | 31.30 | 49.99 | 49.98 | 49.39 | 56.80 |
| Avg. | 301.69 | 303.64 | 302.61 | 302.66 | 282.45 | 271.47 | 282.45 | **247.08** | 747.57 | 747.69 | 462.82 | 522.38 |

[1] Data was obtained from the Our World in Data Team (OWID) [1].
[2] The time-period of the epidemic, where *1* refers to ascending phases (May – Early August 2022), *2* the peak (Mid-August 2022), *3* for the immediate declining phase (Mid-August – Mid-November 2022), and *4* for the tail-end of the epidemic (Mid-November 2022 - Current).
[3] The models of interest include models of the spatial-wave framework (SW), and *n*-sub-epidemic frameworks (SE), a simple linear regression model, Facebook's Prophet model (Prophet), a general additive model, and an auto-regressive integrated moving average model (ARIMA). All models were calibrated with 11-weeks' worth of data at a time.
* The bolded values indicate the best performing model(s) at each time-period for each forecasting horizon. Here, *best performing* refers to the model with the lowest metric value.

**Table H3.** Average 95% Prediction Interval Coverage (95% PI Coverage) of the forecasts generated for the US(OWID) [1] (weeks of July 14th, 2022, through February 23rd, 2023) for each forecasting horizon (1-4 weeks), and time-period [2] of the epidemic, and averaged across all forecast periods (*Avg.*) for each model [3] of interest.

| Time Period | SW 1st Ranked | SW 2nd Ranked | SW Weighted Ensemble (2) | SW Unweighted Ensemble (2) | SE 1st Ranked | SE 2nd Ranked | SE Weighted Ensemble (2) | SE Unweighted Ensemble (2) | Simple Linear Regression | Prophet | General Additive Model | ARIMA[3] |
|---|---|---|---|---|---|---|---|---|---|---|---|---|
| **1-Week Forecasting Horizon** | | | | | | | | | | | | |
| 1 | 75.00 | 75.00 | 75.00 | 75.00 | **100.00** | **100.00** | **100.00** | **100.00** | 25.00 | 25.00 | 25.00 | 50.00 |
| 2 | 0.00 | 0.00 | 0.00 | 0.00 | **100.00** | **100.00** | **100.00** | **100.00** | 0.00 | **100.00** | 0.00 | 0.00 |
| 3 | 43.75 | 56.25 | 50.00 | 56.25 | 81.25 | 87.50 | 75.00 | **100.00** | 37.50 | 62.50 | 87.50 | 87.50 |
| 4 | 62.50 | 50.00 | 50.00 | 50.00 | 75.00 | **100.00** | 75.00 | **100.00** | 87.50 | **100.00** | 87.50 | **100.00** |
| Avg. | 51.72 | 55.17 | 51.72 | 55.17 | 82.76 | **93.10** | 79.31 | 100.00 | 48.28 | 68.97 | 75.86 | 82.76 |
| **2-Week Forecasting Horizon** | | | | | | | | | | | | |
| 1 | 37.50 | 37.50 | 37.50 | 37.50 | **62.50** | **62.50** | **62.50** | **62.50** | 25.00 | 25.00 | 12.50 | 37.50 |
| 2 | 0.00 | 0.00 | 0.00 | 0.00 | **100.00** | **100.00** | **100.00** | **100.00** | 0.00 | 50.00 | 0.00 | 0.00 |
| 3 | 53.13 | 56.25 | 56.25 | 56.25 | 71.88 | 84.38 | 71.88 | 87.50 | 37.50 | 50.00 | **90.63** | 87.50 |
| 4 | 56.25 | 50.00 | 56.25 | 56.25 | 87.50 | 100.00 | **93.75** | 100.00 | 87.50 | 100.00 | 87.50 | 100.00 |
| Avg. | 50.00 | 50.00 | 51.72 | 51.72 | 75.86 | 86.21 | 77.59 | **87.93** | 48.28 | 60.34 | 75.86 | 81.03 |
| **3-Week Forecasting Horizon** | | | | | | | | | | | | |
| 1 | 25.00 | 25.00 | 25.00 | 25.00 | **50.00** | **50.00** | **50.00** | **50.00** | 25.00 | 25.00 | 8.33 | 33.33 |
| 2 | 0.00 | 0.00 | 0.00 | 0.00 | **100.00** | **100.00** | **100.00** | **100.00** | 0.00 | 33.33 | 0.00 | 0.00 |
| 3 | 58.33 | 64.58 | 62.50 | 60.42 | 72.92 | 89.58 | 72.92 | **93.75** | 37.50 | 45.83 | 89.58 | 83.33 |
| 4 | 54.17 | 54.17 | 54.17 | 54.17 | 91.67 | 100.00 | 91.67 | **95.83** | 83.33 | 100.00 | 87.50 | 100.00 |
| Avg. | 50.57 | 54.02 | 52.87 | 51.72 | 75.86 | 87.36 | 75.86 | **88.51** | 47.13 | 57.47 | 74.71 | 78.16 |
| **4-Week Forecasting Horizon** | | | | | | | | | | | | |
| 1 | 18.75 | 18.75 | 18.75 | 18.75 | **43.75** | **43.75** | **43.75** | **43.75** | 25.00 | 25.00 | 6.25 | 37.50 |
| 2 | 0.00 | 0.00 | 0.00 | 0.00 | **100.00** | **100.00** | **100.00** | **100.00** | 0.00 | 25.00 | 0.00 | 0.00 |
| 3 | 59.38 | 64.06 | 62.50 | 62.50 | 73.44 | 85.94 | 73.44 | **92.19** | 34.38 | 39.06 | 90.63 | 81.25 |
| 4 | 59.38 | 53.13 | 56.25 | 56.25 | **93.75** | 100.00 | 90.63 | 96.88 | 81.25 | 90.63 | 84.38 | 100.00 |
| Avg. | 51.72 | 52.59 | 52.59 | 52.59 | 75.86 | 84.48 | 75.00 | **87.07** | 44.83 | 50.86 | 74.14 | 77.59 |

[1] Data was obtained from the Our World in Data Team (OWID) [1].
[2] The time-period of the epidemic, where *1* refers to ascending phases (May – Early August 2022), *2* the peak (Mid-August 2022), *3* for the immediate declining phase (Mid-August – Mid-November 2022), and *4* for the tail-end of the epidemic (Mid-November 2022 - Current).
[3] The models of interest include models of the spatial-wave framework (SW), and *n*-sub-epidemic frameworks (SE), a simple linear regression model, Facebook's Prophet model (Prophet), a general additive model, and an auto-regressive integrated moving average model (ARIMA). All models were calibrated with 11-weeks' worth of data at a time.
* The bolded values indicate the best performing model(s) at each time-period for each forecasting horizon. Here, *best performing* corresponds to the model which produces coverage closest to 95%.
** Each value is shown as a percentage (%).

**Table H4.** Average Weighted Interval Scores (WIS) of the forecasts generated for the US(OWID) [1] (weeks of July 14th, 2022, through February 23rd, 2023) for each forecasting horizon (1-4 weeks), and time-period [2] of the epidemic, and averaged across all forecast periods (*Avg.*) for each model [3] of interest.

| Time Period | SW 1st Ranked | SW 2nd Ranked | SW Weighted Ensemble (2) | SW Unweighted Ensemble (2) | SE 1st Ranked | SE 2nd Ranked | SE Weighted Ensemble (2) | SE Unweighted Ensemble (2) | Simple Linear Regression | Prophet | General Additive Model | ARIMA[3] |
|---|---|---|---|---|---|---|---|---|---|---|---|---|
| **1-Week Forecasting Horizon** | | | | | | | | | | | | |
| 1 | 191.98 | 195.78 | 193.98 | 193.28 | 115.69 | **90.06** | 115.29 | 94.33 | 841.43 | 786.98 | 405.45 | 435.70 |
| 2 | 747.30 | 747.40 | 747.35 | 750.97 | **68.43** | 204.60 | 69.54 | 119.88 | 420.66 | 372.96 | 521.74 | 476.64 |
| 3 | 185.53 | 182.93 | 183.54 | 184.01 | 115.59 | 108.60 | 115.28 | **101.29** | 532.23 | 485.04 | 186.62 | 220.85 |
| 4 | **19.76** | 20.11 | 20.00 | 19.88 | 20.67 | 48.05 | 20.77 | 31.72 | 30.60 | 28.34 | 33.32 | 36.15 |
| Avg. | 160.06 | 159.25 | 159.30 | 159.56 | 87.79 | 92.65 | 87.63 | **81.78** | 432.65 | 396.84 | 186.07 | 208.35 |
| **2-Week Forecasting Horizon** | | | | | | | | | | | | |
| 1 | 447.34 | 453.90 | 448.45 | 448.83 | 365.52 | 359.40 | 365.64 | **339.36** | 888.44 | 843.60 | 587.99 | 541.87 |
| 2 | 752.22 | 756.15 | 751.56 | 753.79 | 97.75 | 252.06 | **96.80** | 158.13 | 858.96 | 807.27 | 846.77 | 846.22 |
| 3 | 180.15 | 179.21 | 179.18 | 179.16 | 132.92 | 116.31 | 132.42 | **110.00** | 606.28 | 568.97 | 182.35 | 244.55 |
| 4 | 19.76 | 20.14 | 19.90 | 20.00 | **19.54** | 52.40 | 19.64 | 25.53 | 32.63 | 30.01 | 36.73 | 39.96 |
| Avg. | 192.49 | 193.11 | 192.12 | 192.27 | 132.52 | 136.89 | 132.25 | **119.99** | 495.66 | 466.39 | 221.04 | 249.87 |
| **3-Week Forecasting Horizon** | | | | | | | | | | | | |
| 1 | 714.22 | 723.25 | 717.33 | 719.09 | 640.72 | 628.54 | 640.37 | **599.70** | 954.16 | 918.26 | 897.75 | 724.59 |
| 2 | 831.60 | 834.73 | 834.05 | 833.28 | 196.16 | 331.30 | **195.30** | 251.88 | 1199.31 | 1154.97 | 1071.55 | 1128.45 |
| 3 | 169.84 | 169.72 | 169.45 | 169.61 | 137.31 | 136.42 | 137.05 | **117.17** | 677.86 | 647.93 | 200.22 | 284.72 |
| 4 | 20.12 | 20.34 | 20.27 | 20.24 | 18.82 | 58.32 | **18.75** | 36.10 | 33.91 | 31.77 | 37.76 | 43.25 |
| Avg. | 226.44 | 227.79 | 226.78 | 227.08 | 176.09 | 189.47 | 175.85 | **166.01** | 556.31 | 532.73 | 281.66 | 307.87 |
| **4-Week Forecasting Horizon** | | | | | | | | | | | | |
| 1 | 919.47 | 928.52 | 922.26 | 925.30 | 851.91 | 803.05 | 852.13 | **784.52** | 990.47 | 962.92 | 1230.19 | 927.05 |
| 2 | 784.84 | 788.92 | 786.13 | 787.54 | 215.55 | 294.03 | **215.21** | 241.65 | 1631.78 | 1600.93 | 1387.71 | 1508.95 |
| 3 | 167.76 | 167.45 | 167.52 | 167.47 | 145.17 | 159.78 | 145.42 | **125.87** | 739.83 | 718.42 | 200.19 | 305.11 |
| 4 | 18.16 | 18.54 | 18.33 | 18.35 | **17.55** | 66.23 | 17.55 | 40.27 | 32.59 | 30.97 | 37.86 | 46.03 |
| Avg. | 251.45 | 252.78 | 251.80 | 252.24 | 209.87 | 227.33 | 210.03 | **197.10** | 610.05 | 592.93 | 338.43 | 360.94 |

[1] Data was obtained from the Our World in Data Team (OWID) [1].
[2] The time-period of the epidemic, where *1* refers to ascending phases (May – Early August 2022), *2* the peak (Mid-August 2022), *3* for the immediate declining phase (Mid-August – Mid-November 2022), and *4* for the tail-end of the epidemic (Mid-November 2022 - Current).
[3] The models of interest include models of the spatial-wave framework (SW), and *n*-sub-epidemic frameworks (SE), a simple linear regression model, Facebook's Prophet model (Prophet), a general additive model, and an auto-regressive integrated moving average model (ARIMA). All models were calibrated with 11-weeks' worth of data at a time.
* The bolded values indicate the best performing model(s) at each time-period for each forecasting horizon. Here, *best performing* refers to the model with the lowest metric value.

**Table H5.** Skills scores[1] for overall average forecast performance metrics (weeks of July 14th, 2022, through February 23rd, 2023) comparing the ARIMA[2] model to the *n*-sub-epidemic (SE) and spatial-wave (SW) frameworks across forecasting horizons (1-4 weeks) for the US(OWID).[3]

| **Baseline[4]** | ARIMA | | | | | | | |
|---|---|---|---|---|---|---|---|---|
| **Comparison[5]** | SW 1st Ranked | SW 2nd Ranked | SW Weighted Ensemble (2) | SW Unweighted Ensemble (2) | SE 1st Ranked | SE 2nd Ranked | SE Weighted Ensemble (2) | SE Unweighted Ensemble (2) |
| **Mean Squared Error (MSE)** | | | | | | | | |
| 1-Week | 47.98 | 48.06 | 48.08 | 48.29 | 79.76 | 78.65 | 79.76 | 82.95 |
| 2-Week | 52.00 | 51.03 | 51.63 | 51.29 | 67.05 | 70.32 | 67.05 | 71.48 |
| 3-Week | 55.10 | 54.16 | 54.63 | 54.93 | 61.46 | 63.36 | 61.46 | 66.03 |
| 4-Week | 62.32 | 61.64 | 61.98 | 61.98 | 64.37 | 68.73 | 64.37 | 70.08 |
| **Mean Absolute Error (MAE)** | | | | | | | | |
| 1-Week | 37.74 | 37.83 | 37.99 | 37.84 | 59.39 | 54.86 | 59.39 | 60.91 |
| 2-Week | 35.90 | 35.51 | 35.86 | 35.54 | 48.32 | 49.88 | 48.32 | 52.36 |
| 3-Week | 38.87 | 38.40 | 38.64 | 38.78 | 45.78 | 45.74 | 45.78 | 50.62 |
| 4-Week | 42.25 | 41.87 | 42.07 | 42.06 | 45.93 | 48.03 | 45.93 | 52.70 |
| **95% Prediction Interval Coverage (95% PI) *** | | | | | | | | |
| 1-Week | -55.77 | -54.44 | -53.16 | -53.95 | 69.33 | 43.88 | 67.84 | 70.51 |
| 2-Week | -67.50 | -67.45 | -65.82 | -65.90 | 38.05 | 6.60 | 37.90 | 41.80 |
| 3-Week | -48.48 | -49.38 | -48.50 | -48.38 | 24.22 | -16.90 | 24.70 | 30.61 |
| 4-Week | -32.22 | -32.78 | -31.65 | -32.14 | 21.25 | -51.58 | 20.75 | 27.95 |
| **Weighted Interval Score (WIS)** | | | | | | | | |
| 1-Week | 23.18 | 23.57 | 23.54 | 23.42 | 57.86 | 55.53 | 57.94 | 60.75 |
| 2-Week | 22.97 | 22.71 | 23.11 | 23.05 | 46.97 | 45.21 | 47.07 | 51.98 |
| 3-Week | 26.45 | 26.01 | 26.34 | 26.24 | 42.80 | 38.46 | 42.88 | 46.08 |
| 4-Week | 30.33 | 29.97 | 30.24 | 30.11 | 41.85 | 37.02 | 41.81 | 45.39 |

[1] Skill scores are calculated by subtracting the mean metric score for the comparison model from the benchmark model, dividing by the mean metric score for the benchmark model and multiplying by 100. They are shown here as a percentage.
[2] Auto-regressive integrated moving average
[3] Data was obtained from the Our World in Data Team (OWID) [1].
[4] The ARIMA model used an 11-week calibration period for all forecasts.
[5] Comparison models used only a 11-week calibration period and were comprised of the 1st ranked, 2nd ranked, weighted (W) ensemble, and unweighted (UW) ensemble models from both the *n*-sub-epidemic (SE) and spatial-wave (SW) frameworks.
* Skill scores for 95% PI coverage are based on average Winkler scores.

**Table I1.** Average Mean Squared Error (MSE) of the forecasts generated for the World [1] (weeks of July 14th, 2022, through February 23rd, 2023) for each forecasting horizon (1-4 weeks), and time-period [2] of the epidemic, and averaged across all forecast periods (*Avg.*) for each model [3] of interest.

| Time Period | SW 1st Ranked | SW 2nd Ranked | SW Weighted Ensemble (2) | SW Unweighted Ensemble (2) | SE 1st Ranked | SE 2nd Ranked | SE Weighted Ensemble (2) | SE Unweighted Ensemble (2) | Simple Linear Regression | Prophet | General Additive Model | ARIMA[3] |
|---|---|---|---|---|---|---|---|---|---|---|---|---|
| **1-Week Forecasting Horizon** | | | | | | | | | | | | |
| 1 | 313492.75 | 305516.68 | 322949.88 | 299977.13 | 317162.25 | 961204.23 | 317162.25 | 444060.96 | 1615339.06 | 1617970.91 | **260060.48** | 971506.39 |
| 2 | 369019.50 | 378059.28 | 398810.51 | 357539.60 | 332382.39 | 1395300.62 | 332382.39 | **71759.40** | 1304867.29 | 1303870.42 | 790933.58 | 772502.44 |
| 3 | 517064.39 | 91305.69 | 151419.97 | 119528.28 | **90087.66** | 275012.96 | **90087.66** | 108211.95 | 2387960.00 | 2382490.73 | 255292.43 | 523802.32 |
| 4 | **13709.68** | 15366.98 | 14190.02 | 14056.78 | 18453.47 | 19581.89 | 18453.47 | 17639.06 | 56448.65 | 56285.52 | 46129.75 | 43387.82 |
| Avg. | 364295.85 | 114911.53 | 153101.39 | 129151.34 | **112997.10** | 361603.71 | **112997.10** | 118578.56 | 1670559.04 | 1667517.05 | 242238.90 | 471306.15 |
| **2-Week Forecasting Horizon** | | | | | | | | | | | | |
| 1 | 982126.35 | 974673.64 | 931783.31 | 970658.16 | 953195.62 | 2062787.62 | 953195.62 | 730768.04 | 1766024.50 | 1768565.69 | **635530.18** | 2041999.40 |
| 2 | 905200.23 | 920828.68 | 872493.34 | 916896.02 | 825649.50 | 1778146.83 | 825649.50 | **210173.74** | 3467991.46 | 3465614.04 | 2275221.29 | 2540188.20 |
| 3 | 442582.04 | 97871.26 | 145721.95 | 112977.81 | 93137.81 | 261611.00 | 93137.81 | **91349.16** | 3329575.99 | 3322840.61 | 255240.04 | 683853.38 |
| 4 | **11086.73** | 11997.35 | 11369.55 | 11122.67 | 15553.85 | 16536.35 | 15553.85 | 14107.99 | 63152.20 | 63113.74 | 45175.97 | 33163.89 |
| Avg. | 426147.63 | 224602.46 | 244730.92 | 232560.28 | 213900.19 | 493372.34 | 213900.19 | **147046.32** | 2388928.05 | 2385069.37 | 383184.33 | 795311.36 |
| **3-Week Forecasting Horizon** | | | | | | | | | | | | |
| 1 | 2702017.62 | 2625630.95 | 2582440.85 | 2715357.02 | 2569737.52 | 4298670.26 | 2569737.52 | 1764149.51 | 1870046.72 | 1872073.36 | **1618414.38** | 4463960.82 |
| 2 | 1618850.59 | 1645226.13 | 1768795.81 | 1660010.56 | 1501048.62 | 1933204.00 | 1501048.62 | **342079.16** | 7019823.39 | 7015532.39 | 4929264.17 | 5602136.75 |
| 3 | 450629.46 | 110714.63 | 175617.63 | 130317.90 | 104989.60 | 431969.05 | 104989.60 | **103725.46** | 4380214.00 | 4371851.84 | 435085.34 | 1311756.35 |
| 4 | 11199.31 | 11964.58 | **10817.33** | 11423.71 | 15432.71 | 17156.24 | 15432.71 | 14274.62 | 64479.28 | 64446.21 | 48085.46 | 31555.16 |
| Avg. | 658029.31 | 452870.55 | 494694.32 | 474533.19 | 434626.27 | 835378.84 | 434626.27 | **270340.07** | 3260854.48 | 3255858.26 | 774028.29 | 1624720.69 |
| **4-Week Forecasting Horizon** | | | | | | | | | | | | |
| 1 | 5960275.92 | 5695426.04 | 5754435.64 | 5846959.78 | 5611168.27 | 7435797.59 | 5611168.27 | 3265884.22 | **2096470.57** | 2097392.61 | 3618676.56 | 8239008.34 |
| 2 | 2331558.55 | 2343828.34 | 2331743.27 | 2431754.99 | 2144490.84 | 1949977.15 | 2144490.84 | **495250.01** | 11874601.43 | 11868051.35 | 8637036.65 | 9855377.37 |
| 3 | 442890.13 | 114236.53 | 150308.85 | 112866.44 | 107058.52 | 589972.33 | 107058.52 | **102706.61** | 5521451.34 | 5511302.93 | 491988.43 | 1698134.20 |
| 4 | 11546.60 | 12126.03 | 11447.08 | **11439.80** | 15898.02 | 17269.27 | 15898.02 | 14478.18 | 63128.56 | 63103.76 | 49810.42 | 32432.80 |
| Avg. | 1039789.77 | 820718.55 | 846971.50 | 841489.56 | 794957.50 | 1253715.92 | 794957.50 | **435707.32** | 4287765.08 | 4281453.68 | 1270433.76 | 2535278.99 |

[1] Data was obtained from the Our World in Data Team (OWID) [1].

[2] The time-period of the epidemic, where *1* refers to ascending phases (May – July 2022), *2* the peak (Early August - Mid-August 2022), *3* for the immediate declining phase (Mid-August – Early December 2022), and *4* for the tail-end of the epidemic (Mid-December 2022 - Current).

[3] The models of interest include models of the spatial-wave framework (SW), and *n*-sub-epidemic frameworks (SE), a simple linear regression model, Facebook's Prophet model (Prophet), a general additive model, and an auto-regressive integrated moving average model (ARIMA). All models were calibrated with 11-weeks' worth of data at a time.

* The bolded values indicate the best performing model(s) at each time-period for each forecasting horizon. Here, *best performing* refers to the model with the lowest metric value.

**Table I2.** Average Mean Absolute Error (MAE) of the forecasts generated for the World [1] (weeks of July 14th, 2022, through February 23rd, 2023) for each forecasting horizon (1-4 weeks), and time-period [2] of the epidemic, and averaged across all forecast periods (*Avg.*) for each model [3] of interest.

| Time Period | SW 1st Ranked | SW 2nd Ranked | SW Weighted Ensemble (2) | SW Unweighted Ensemble (2) | SE 1st Ranked | SE 2nd Ranked | SE Weighted Ensemble (2) | SE Unweighted Ensemble (2) | Simple Linear Regression | Prophet | General Additive Model | ARIMA[3] |
|---|---|---|---|---|---|---|---|---|---|---|---|---|
| **1-Week Forecasting Horizon** | | | | | | | | | | | | |
| 1 | 519.51 | 509.13 | 523.75 | 503.91 | 509.39 | 904.04 | 509.39 | 547.04 | 1206.95 | 1208.14 | **407.85** | 965.45 |
| 2 | 597.14 | 601.82 | 622.73 | 584.99 | 568.05 | 858.83 | 568.05 | **267.79** | 1002.42 | 1001.36 | 858.54 | 868.80 |
| 3 | 387.46 | **240.88** | 294.14 | 275.79 | 242.72 | 451.27 | 242.72 | 299.64 | 1196.49 | 1195.56 | 405.20 | 579.92 |
| 4 | 98.59 | 102.01 | 99.54 | **98.58** | 117.53 | 116.82 | 117.53 | 114.05 | 212.56 | 212.57 | 183.77 | 136.03 |
| Avg. | 345.85 | **260.00** | 293.58 | 277.94 | 262.53 | 445.49 | 262.53 | 278.24 | 946.69 | 946.20 | 383.29 | 532.58 |
| **2-Week Forecasting Horizon** | | | | | | | | | | | | |
| 1 | 854.65 | 854.09 | 834.40 | 849.64 | 846.71 | 1306.66 | 846.71 | 744.71 | 1158.76 | 1159.40 | **649.48** | 1364.04 |
| 2 | 885.44 | 891.74 | 872.22 | 883.90 | 848.16 | 970.60 | 848.16 | **436.02** | 1629.30 | 1628.16 | 1374.16 | 1461.20 |
| 3 | 377.60 | 254.74 | 293.08 | 272.44 | **248.43** | 412.66 | **248.43** | 265.84 | 1362.89 | 1362.02 | 425.77 | 627.89 |
| 4 | 88.79 | 91.21 | 89.78 | **88.15** | 105.98 | 111.73 | 105.98 | 96.69 | 226.72 | 226.93 | 176.44 | 128.44 |
| Avg. | 392.26 | 321.20 | 339.95 | 329.84 | 317.30 | 470.98 | 317.30 | **286.29** | 1085.90 | 1085.43 | 454.14 | 640.95 |
| **3-Week Forecasting Horizon** | | | | | | | | | | | | |
| 1 | 1313.54 | 1305.72 | 1299.61 | 1329.39 | 1297.74 | 1821.53 | 1297.74 | 1113.88 | 1203.49 | 1203.95 | **988.08** | 1895.62 |
| 2 | 1140.04 | 1151.00 | 1187.06 | 1151.94 | 1100.38 | 1013.06 | 1100.38 | **546.10** | 2314.01 | 2312.80 | 1947.61 | 2111.43 |
| 3 | 391.06 | 267.97 | 315.00 | 289.72 | **256.74** | 470.48 | **256.74** | 267.22 | 1514.05 | 1513.18 | 538.88 | 811.97 |
| 4 | 92.09 | 94.72 | **90.06** | 92.70 | 109.43 | 115.29 | 109.43 | 102.63 | 232.86 | 232.98 | 188.27 | 129.43 |
| Avg. | 465.98 | 394.40 | 422.70 | 409.18 | 387.05 | 561.93 | 387.05 | **334.31** | 1227.84 | 1227.32 | 597.88 | 848.94 |
| **4-Week Forecasting Horizon** | | | | | | | | | | | | |
| 1 | 1814.44 | 1796.49 | 1785.97 | 1806.56 | 1784.97 | 2322.64 | 1784.97 | 1354.93 | **1253.70** | 1253.85 | 1364.50 | 2430.45 |
| 2 | 1347.18 | 1353.95 | 1348.39 | 1369.39 | 1293.66 | 1039.85 | 1293.66 | **640.50** | 3007.68 | 3006.39 | 2530.01 | 2770.62 |
| 3 | 374.66 | 262.36 | 286.30 | 263.63 | **250.71** | 508.05 | **250.71** | 261.26 | 1643.69 | 1642.84 | 574.66 | 896.28 |
| 4 | 91.98 | 93.50 | **91.23** | 91.25 | 108.39 | 116.00 | 108.39 | 102.95 | 231.97 | 232.05 | 189.58 | 137.58 |
| Avg. | 522.44 | 455.58 | 467.60 | 457.89 | 447.00 | 637.81 | 447.00 | **362.34** | 1356.65 | 1356.10 | 698.27 | 1001.12 |

[1] Data was obtained from the Our World in Data Team (OWID) [1].
[2] The time-period of the epidemic, where *1* refers to ascending phases (May – July 2022), *2* the peak (Early August - Mid-August 2022), *3* for the immediate declining phase (Mid-August – Early December 2022), and *4* for the tail-end of the epidemic (Mid-December 2022 - Current).
[3] The models of interest include models of the spatial-wave framework (SW), and *n*-sub-epidemic frameworks (SE), a simple linear regression model, Facebook's Prophet model (Prophet), a general additive model, and an auto-regressive integrated moving average model (ARIMA). All models were calibrated with 11-weeks' worth of data at a time.
* The bolded values indicate the best performing model(s) at each time-period for each forecasting horizon. Here, *best performing* refers to the model with the lowest metric value.

**Table I3.** Average 95% Prediction Interval Coverage (95% PI Coverage) of the forecasts generated for the World [1] (weeks of July 14th, 2022, through February 23rd, 2023) for each forecasting horizon (1-4 weeks), and time-period [2] of the epidemic, and averaged across all forecast periods (*Avg.*) for each model [3] of interest.

| Time Period | SW 1st Ranked | SW 2nd Ranked | SW Weighted Ensemble (2) | SW Unweighted Ensemble (2) | SE 1st Ranked | SE 2nd Ranked | SE Weighted Ensemble (2) | SE Unweighted Ensemble (2) | Simple Linear Regression | Prophet | General Additive Model | ARIMA[3] |
|---|---|---|---|---|---|---|---|---|---|---|---|---|
| **1-Week Forecasting Horizon** | | | | | | | | | | | | |
| 1 | **66.67** | **66.67** | **66.67** | **66.67** | **66.67** | 33.33 | **66.67** | **66.67** | 33.33 | 33.33 | **66.67** | 33.33 |
| 2 | **100.00** | **100.00** | **100.00** | **100.00** | **100.00** | **100.00** | **100.00** | **100.00** | 50.00 | 50.00 | 50.00 | 50.00 |
| 3 | 88.24 | **94.12** | **94.12** | **94.12** | **94.12** | 88.24 | **94.12** | **94.12** | 35.29 | 58.82 | 82.35 | 70.59 |
| 4 | **100.00** | **100.00** | **100.00** | **100.00** | **100.00** | **100.00** | **100.00** | **100.00** | 57.14 | **100.00** | 85.71 | **100.00** |
| Avg. | 89.66 | **93.10** | **93.10** | **93.10** | **93.10** | 86.21 | **93.10** | **93.10** | 41.38 | 65.52 | 79.31 | 72.41 |
| **2-Week Forecasting Horizon** | | | | | | | | | | | | |
| 1 | 66.67 | 66.67 | 66.67 | 66.67 | **83.33** | 50.00 | 66.67 | **83.33** | 33.33 | 33.33 | 50.00 | 33.33 |
| 2 | **100.00** | **100.00** | **100.00** | **100.00** | **100.00** | **100.00** | **100.00** | **100.00** | 25.00 | 25.00 | 25.00 | 25.00 |
| 3 | 88.24 | **94.12** | **94.12** | **94.12** | **94.12** | **94.12** | **94.12** | 97.06 | 26.47 | 44.12 | 85.29 | 70.59 |
| 4 | **100.00** | **100.00** | **100.00** | **100.00** | **100.00** | **100.00** | **100.00** | **100.00** | 50.00 | 85.71 | 85.71 | **100.00** |
| Avg. | 89.66 | 93.10 | 93.10 | 93.10 | **94.83** | 91.38 | 93.10 | 96.55 | 32.76 | 51.72 | 77.59 | 70.69 |
| **3-Week Forecasting Horizon** | | | | | | | | | | | | |
| 1 | 55.56 | 55.56 | 55.56 | 55.56 | 55.56 | 44.44 | 55.56 | **77.78** | 33.33 | 44.44 | 44.44 | 44.44 |
| 2 | 83.33 | 66.67 | 83.33 | 83.33 | **100.00** | **100.00** | **100.00** | **100.00** | 16.67 | 16.67 | 16.67 | 16.67 |
| 3 | 86.27 | **92.16** | **92.16** | **92.16** | **92.16** | **92.16** | **92.16** | 98.04 | 23.53 | 37.25 | 84.31 | 68.63 |
| 4 | **100.00** | **100.00** | **100.00** | **100.00** | **100.00** | **100.00** | **100.00** | **100.00** | 42.86 | 57.14 | 85.71 | **100.00** |
| Avg. | 86.21 | 88.51 | 89.66 | 89.66 | 90.80 | 89.66 | 90.80 | **96.55** | 28.74 | 41.38 | 75.86 | 70.11 |
| **4-Week Forecasting Horizon** | | | | | | | | | | | | |
| 1 | 50.00 | 50.00 | 50.00 | 50.00 | 50.00 | 41.67 | 50.00 | **75.00** | 33.33 | 33.33 | 41.67 | 50.00 |
| 2 | 87.50 | 75.00 | 87.50 | 87.50 | **100.00** | **100.00** | **100.00** | **100.00** | 12.50 | 12.50 | 12.50 | 12.50 |
| 3 | 86.76 | 91.18 | **92.65** | **92.65** | 91.18 | 91.18 | 91.18 | 98.53 | 22.06 | 30.88 | 83.82 | 64.71 |
| 4 | **100.00** | **100.00** | **100.00** | **100.00** | **100.00** | **100.00** | **100.00** | **100.00** | 42.86 | 57.14 | 85.71 | **100.00** |
| Avg. | 86.21 | 87.93 | 89.66 | 89.66 | 89.66 | 88.79 | 89.66 | **96.55** | 27.59 | 36.21 | 75.00 | 68.10 |

[1] Data was obtained from the Our World in Data Team (OWID) [1].
[2] The time-period of the epidemic, where *1* refers to ascending phases (May – July 2022), *2* the peak (Early August - Mid-August 2022), *3* for the immediate declining phase (Mid-August – Early December 2022), and *4* for the tail-end of the epidemic (Mid-December 2022 - Current).
[3] The models of interest include models of the spatial-wave framework (SW), and *n*-sub-epidemic frameworks (SE), a simple linear regression model, Facebook's Prophet model (Prophet), a general additive model, and an auto-regressive integrated moving average model (ARIMA). All models were calibrated with 11-weeks' worth of data at a time.
* The bolded values indicate the best performing model(s) at each time-period for each forecasting horizon. Here, *best performing* corresponds to the model which produces coverage closest to 95%.
** Each value is shown as a percentage (%).

**Table I4.** Average Weighted Interval Scores (WIS) of the forecasts generated for the World [1] (weeks of July 14th, 2022, through February 23rd, 2023) for each forecasting horizon (1-4 weeks), and time-period [2] of the epidemic, and averaged across all forecast periods (*Avg.*) for each model [3] of interest.

| Time Period | SW 1st Ranked | SW 2nd Ranked | SW Weighted Ensemble (2) | SW Unweighted Ensemble (2) | SE 1st Ranked | SE 2nd Ranked | SE Weighted Ensemble (2) | SE Unweighted Ensemble (2) | Simple Linear Regression | Prophet | General Additive Model | ARIMA[3] |
|---|---|---|---|---|---|---|---|---|---|---|---|---|
| **1-Week Forecasting Horizon** | | | | | | | | | | | | |
| 1 | 312.90 | 312.43 | **310.61** | 311.91 | 323.65 | 642.15 | 326.35 | 385.10 | 957.73 | 885.27 | 311.25 | 670.67 |
| 2 | 340.17 | 356.52 | 344.16 | 347.94 | 325.19 | 611.84 | 325.85 | **286.32** | 809.55 | 755.60 | 614.41 | 556.04 |
| 3 | 299.47 | **161.87** | 192.42 | 192.86 | 163.85 | 264.24 | 164.77 | 187.84 | 909.18 | 827.07 | 263.98 | 387.55 |
| 4 | **59.45** | 60.98 | 59.53 | 59.58 | 65.53 | 79.28 | 65.15 | 68.21 | 136.82 | 121.73 | 112.16 | 95.17 |
| Avg. | 245.73 | **166.52** | 183.03 | 183.70 | 167.77 | 282.66 | 168.55 | 186.16 | 720.90 | 657.91 | 256.39 | 357.88 |
| **2-Week Forecasting Horizon** | | | | | | | | | | | | |
| 1 | 520.01 | 524.41 | 523.54 | 523.09 | 536.53 | 910.68 | 539.12 | 545.52 | 954.94 | 896.50 | **491.51** | 919.48 |
| 2 | 511.78 | 532.78 | 520.31 | 526.11 | 501.70 | 673.00 | 500.84 | **368.43** | 1411.27 | 1366.37 | 1045.66 | 1070.33 |
| 3 | 283.87 | 165.18 | 187.08 | 188.03 | **164.42** | 281.38 | 165.31 | 181.55 | 1077.44 | 1002.59 | 264.78 | 432.53 |
| 4 | 55.61 | 55.98 | 55.70 | **55.45** | 61.40 | 74.04 | 61.62 | 62.67 | 148.09 | 133.17 | 111.98 | 92.65 |
| Avg. | 268.92 | 201.34 | 213.16 | 214.01 | **201.31** | 323.44 | 202.09 | 203.40 | 863.46 | 806.84 | 305.21 | 444.85 |
| **3-Week Forecasting Horizon** | | | | | | | | | | | | |
| 1 | 844.74 | 841.62 | 831.43 | 841.85 | 867.66 | 1299.01 | 868.71 | 824.98 | 966.85 | 920.63 | **770.80** | 1274.77 |
| 2 | 675.46 | 700.05 | 689.51 | 685.82 | 662.18 | 693.90 | 660.29 | **429.22** | 2077.88 | 2028.81 | 1538.85 | 1659.40 |
| 3 | 291.35 | 172.05 | 195.64 | 195.67 | **171.35** | 946.17 | 171.47 | 233.26 | 1228.81 | 1169.69 | 330.90 | 575.33 |
| 4 | **55.93** | 56.68 | 56.35 | 56.48 | 61.57 | 76.79 | 61.85 | 63.50 | 160.22 | 148.16 | 120.22 | 99.55 |
| Avg. | 318.26 | **249.89** | 261.85 | 262.72 | 250.73 | 755.42 | 250.85 | 267.01 | 1002.33 | 956.60 | 408.86 | 607.61 |
| **4-Week Forecasting Horizon** | | | | | | | | | | | | |
| 1 | 1234.65 | 1223.36 | 1224.74 | 1224.29 | 1259.12 | 1704.07 | 1262.44 | 1125.82 | 1013.21 | **981.22** | 1124.65 | 1649.33 |
| 2 | 795.41 | 828.83 | 808.02 | 815.10 | 779.97 | 695.66 | 781.87 | **471.39** | 2755.08 | 2720.66 | 2042.80 | 2265.79 |
| 3 | 276.92 | 171.26 | 187.85 | 187.83 | **169.66** | 61344.73 | 169.76 | 747.77 | 1356.60 | 1313.00 | 361.99 | 659.55 |
| 4 | 55.84 | 56.00 | **55.72** | 55.98 | 61.80 | 77.47 | 61.86 | 63.31 | 167.99 | 159.01 | 125.07 | 106.81 |
| Avg. | 358.39 | **297.63** | 305.99 | 306.49 | 298.42 | 36203.66 | 298.97 | 602.60 | 1130.61 | 1097.21 | 499.61 | 739.30 |

[1] Data was obtained from the Our World in Data Team (OWID) [1].
[2] The time-period of the epidemic, where *1* refers to ascending phases (May – July 2022), *2* the peak (Early August - Mid-August 2022), *3* for the immediate declining phase (Mid-August – Early December 2022), and *4* for the tail-end of the epidemic (Mid-December 2022 - Current).
[3] The models of interest include models of the spatial-wave framework (SW), and *n*-sub-epidemic frameworks (SE), a simple linear regression model, Facebook's Prophet model (Prophet), a general additive model, and an auto-regressive integrated moving average model (ARIMA). All models were calibrated with 11-weeks' worth of data at a time.
* The bolded values indicate the best performing model(s) at each time-period for each forecasting horizon. Here, *best performing* refers to the model with the lowest metric value.

**Table I5.** Skills scores[1] for overall average forecast performance metrics (weeks of July 14th, 2022, through February 23rd, 2023) comparing the ARIMA[2] model to the *n*-sub-epidemic (SE) and spatial-wave (SW) frameworks across forecasting horizons (1-4 weeks) for the World.[3]

| **Baseline[4]** | ARIMA | | | | | | | |
|---|---|---|---|---|---|---|---|---|
| **Comparison[5]** | SW 1st Ranked | SW 2nd Ranked | SW Weighted Ensemble (2) | SW Unweighted Ensemble (2) | SE 1st Ranked | SE 2nd Ranked | SE Weighted Ensemble (2) | SE Unweighted Ensemble (2) |
| **Mean Squared Error (MSE)** | | | | | | | | |
| 1-Week | 22.71 | 75.62 | 67.52 | 72.60 | 76.02 | 23.28 | 76.02 | 74.84 |
| 2-Week | 46.42 | 71.76 | 69.23 | 70.76 | 73.10 | 37.96 | 73.10 | 81.51 |
| 3-Week | 59.50 | 72.13 | 69.55 | 70.79 | 73.25 | 48.58 | 73.25 | 83.36 |
| 4-Week | 58.99 | 67.63 | 66.59 | 66.81 | 68.64 | 50.55 | 68.64 | 82.81 |
| **Mean Absolute Error (MAE)** | | | | | | | | |
| 1-Week | 35.06 | 51.18 | 44.88 | 47.81 | 50.71 | 16.35 | 50.71 | 47.76 |
| 2-Week | 38.80 | 49.89 | 46.96 | 48.54 | 50.50 | 26.52 | 50.50 | 55.33 |
| 3-Week | 45.11 | 53.54 | 50.21 | 51.80 | 54.41 | 33.81 | 54.41 | 60.62 |
| 4-Week | 47.81 | 54.49 | 53.29 | 54.26 | 55.35 | 36.29 | 55.35 | 63.81 |
| **95% Prediction Interval Coverage (95% PI) *** | | | | | | | | |
| 1-Week | -25.72 | 54.10 | 51.74 | 51.36 | 52.29 | -0.87 | 51.45 | 37.80 |
| 2-Week | 23.38 | 68.71 | 67.15 | 66.97 | 68.48 | 7.74 | 68.02 | 53.67 |
| 3-Week | 43.59 | 72.22 | 70.46 | 70.23 | 70.89 | -113.92 | 70.72 | 49.28 |
| 4-Week | 55.98 | 73.50 | 72.02 | 71.51 | 70.39 | -1349.30 | 70.21 | 48.96 |
| **Weighted Interval Score (WIS)** | | | | | | | | |
| 1-Week | 31.34 | 53.47 | 48.86 | 48.67 | 53.12 | 21.02 | 52.90 | 47.98 |
| 2-Week | 39.55 | 54.74 | 52.08 | 51.89 | 54.75 | 27.29 | 54.57 | 54.28 |
| 3-Week | 47.62 | 58.87 | 56.91 | 56.76 | 58.73 | -24.33 | 58.72 | 56.06 |
| 4-Week | 51.52 | 59.74 | 58.61 | 58.54 | 59.63 | -4797.05 | 59.56 | 18.49 |

[1] Skill scores are calculated by subtracting the mean metric score for the comparison model from the benchmark model, dividing by the mean metric score for the benchmark model and multiplying by 100. They are shown here as a percentage.
[2] Auto-regressive integrated moving average
[3] Data was obtained from the Our World in Data Team (OWID) [1].
[4] The ARIMA model used an 11-week calibration period for all forecasts.
[5] Comparison models used only a 11-week calibration period and were comprised of the 1st ranked, 2nd ranked, weighted (W) ensemble, and unweighted (UW) ensemble models from both the *n*-sub-epidemic (SE) and spatial-wave (SW) frameworks.
* Skill scores for 95% PI coverage are based on average Winkler scores.
